## Supplementary material for "Optimising supervised machine learning algorithms predicting cigarette cravings and lapses for a smoking cessation just-in-time adaptive intervention (JITAI)": S1 Appendix Surveys

### Online screening survey

| **What is your age (in years)?** | 0-99 |
| --- | --- |
| **Do you smoke cigarettes at all nowadays?** | 1) No |
|  | 2) Yes |
| **How many cigarettes per day do you usually smoke?** | 0-99 |
| **Do you live in London?** | 1) No |
|  | 2) Yes |
| **Are you willing to meet with a researcher at University College London twice during the 10-day study?** | 1) No |
|  | 2) Yes |
| **Do you own a smartphone capable of running the Fitbit and m-Path apps (i.e., Android 8.0 and up; iOS 14.0 and up)?** | 1) No |
|  | 2) Yes |
| **Do you have internet/Wi-Fi access for the duration of the study?** | 1) No |
|  | 2) Yes |
| **Do you smoke cigarettes at all nowadays?** | 1) No |
|  | 2) Yes |
| **Are you willing to set a quit date within 7 days from the initial study visit (and preferably the next day)?** | 1) No |
|  | 2) Yes |
| **Are you willing to wear a Fitbit device and respond to multiple daily surveys (taking a total of 48 minutes per day) on your smartphone for a period of 10 days?** | 1) No |
|  | 2) Yes |
| **Are you able and willing to provide an exhaled carbon monoxide (eCO) measure? Please be aware that individuals with asthma or COPD may find it difficult to provide an eCO measure.** | 1) No |
|  | 2) Yes |
| **Do you have a known history of arrythmias (e.g., atrial fibrillation)?** | 1) No |
|  | 2) Yes |
| **Do you regularly take beta blockers (e.g., atenolol, bisoprolol)?** | 1) No |
|  | 2) Yes |
| **Do you have an implanted cardiac rhythm device?** | 1) No |
|  | 2) Yes |

### Additional baseline survey questions for eligible participants

| **What is your name?** | Free text |
| --- | --- |
| **What is your e-mail address?** | Free text |
| **What is your mobile phone number?** | Free text |
| **Which of the following describes how you think of yourself?** | 1) Male |
|  | 2) Female |
|  | 3) In another way  4) Prefer not to say |
| **What type of job do you have?** | 1) Manual |
|  | 2) Non-manual |
|  | 3) Other (e.g., student, unemployed, retired) |
| **What is your ethnic group?** | 1) Asian or Asian British (any Asian background)  2) Black, Black British, Caribbean or African (any Black, Black British or Caribbean background) |
|  | 3) Mixed or multiple ethnic groups (e.g., White and Black African, White and Asian) |
|  | 4) White (any White background) |
|  | 5) Other ethnic group (e.g., Arab) |
| **Do you have any post-16 educational qualifications?** | 1) No |
|  | 2) Yes |
| **How soon after waking do you have your first cigarette?** | 1) Within 5 minutes |
|  | 2) 6-30 minutes |
|  | 3) 31-60 minutes |
|  | 4) After 60 minutes |
| **Which of the following best describes you?** | 1) I don't want to stop smoking |
|  | 2) I think I should stop smoking but don't really want to |
|  | 3) I want to stop smoking but haven't thought about when |
|  | 4) I really want to stop smoking but don't know when I will |
|  | 5) I want to stop smoking and hope to soon |
|  | 6) I really want to stop smoking and intend to in the next 3 months |
|  | 7) I really want to stop smoking and intend to in the next month |
| **Have you made a serious attempt to quit smoking in the past 12 months? By serious we mean you decided that you would try to make sure you never smoked again.** | 1) No, never |
|  | 2) Yes, but not in the past year |
|  | 3) Yes, in the past year |
| **Have you ever used any of the following to help you stop smoking?** | 1) Nicotine replacement product (e.g. patches/gum/inhaler) without a prescription |
|  | 2) Nicotine replacement product on prescription or given to you by a health professional |
|  | 3) Zyban (bupropion) |
|  | 4) Champix (varenicline) |
|  | 5) E-cigarette or other vaping device |
|  | 6) Attended a Stop Smoking group |
|  | 7) Attended one or more Stop Smoking one-to-one counselling/advice/support sessions |
|  | 8) Phoned a smoking helpline |
|  | 9) A book or booklet |
|  | 10) Visited a website |
|  | 11) Used an application ('app') on a handled computer (smartphone, tablet, PDA) |
|  | 12) None of these |
|  | 13) Other |

### EMA items

| **I feel sad [negative affect]** | 0-10 |
| --- | --- |
| **I feel irritable [negative affect]** | 0-10 |
| **I feel stressed [negative affect]** | 0-10 |
| **I feel anxious [negative affect]** | 0-10 |
| **I feel bored [negative affect]** | 0-10 |
| **What is your bodily pain intensity right now? [pain]** | 0-10 [0 = no pain; 10 = worst pain] |
| **I feel calm [positive affect]** | 0-10 |
| **I feel contented [positive affect]** | 0-10 |
| **I feel happy [positive affect]** | 0-10 |
| **I feel excited [positive affect]** | 0-10 |
| **I feel enthusiastic [positive affect]** | 0-10 |
| **I am craving a cigarette [craving]** | 0-10 |
| **I feel motivated NOT to smoke [motivation]** | 0-10 |
| **I feel confident in my ability NOT to smoke [self-efficacy]** | 0-10 |
| **Who are you with?** | 1) Alone |
|  | 2) With partner/spouse |
|  | 3) With friend(s) |
|  | 4) With child(ren)  5) With relative(s) |
|  | 6) With colleague(s) |
|  | 7) With stranger(s) |
|  | 8) Other |
| **What are you doing?** | 1) Eating/drinking |
|  | 2) Watching TV  3) Listening to music  4) Reading |
|  | 5) Working/studying  6) Walking/exercising |
|  | 7) Caring for child(ren)  8) Socialising |
|  | 9) Scrolling on social media |
|  | 10) Relaxing |
|  | 11) Doing chores  12) Other |
| **Where are you?** | 1) At home |
|  | 2) At school/work |
|  | 3) Outside |
|  | 4) In a restaurant/café/bar |
|  | 5) In a public place (e.g., post office) |
|  | 6) On public transport |
|  | 7) In a private vehicle |
|  | 8) In others’ home |
|  | 9) Other |
| **Are cigarettes available to you right now?** | 1) Easily available |
|  | 2) Available with difficulty |
|  | 3) Not available |
| **Have you consumed any caffeine in the last hour?** | 1) No |
|  | 2) Yes |
| **Have you consumed any alcohol in the last hour?** | 1) No |
|  | 2) Yes |
| **Have you used a nicotine product (e.g., e-cigarette, nicotine gum) in the last hour?** | 1) No |
|  | 2) Yes |
| **Have you smoked (even a puff) in the last hour?** | 1) No |
|  | 2) Yes |
| **Participant-specific variable 1** | 1) No |
|  | 2) Yes |
| **Participant-specific variable 2** | 1) No |
|  | 2) Yes |
| **Participant-specific variable 3** | 1) No |
|  | 2) Yes |
