## Supplementary material for "Optimising supervised machine learning algorithms predicting cigarette cravings and lapses for a smoking cessation just-in-time adaptive intervention (JITAI)": S2 Appendix List of predictors for each model

### Models using all predictors

- Between-subject/time-invariant
  1. Age (numeric)
  2. Gender (binary: Man; Woman)
  3. Cigarettes per day (numeric)
  4. Occupation type (three-level factor: Non-manual; Manual, Other (e.g., student, unemployed, retired)
  5. Post-16 education (binary: yes; no)
  6. Ethnicity (five-level factor: Asian or Asian British (any Asian background); Black, Black British, Caribbean or African (any Black, Black British or Caribbean background); Mixed or multiple ethnic groups (e.g., White and Black African, White and Asian); Other ethnic group (i.e., Arab); White (any White background)
  7. Time to first cigarette (four-level factor: Within 5 minutes; 6-30 minutes; 31-60 minutes; After 60 minutes)
  8. Motivation to stop (six-level factor: I think I should stop smoking but don't really want to; I want to stop smoking but haven't thought about when; I really want to stop smoking but don't know when I will; I want to stop smoking and hope to soon; I really want to stop smoking and intend to in the next 3 months; I really want to stop smoking and intend to in the next month)
  9. Past quit attempt (three-level factor: No, never; Yes, but not in the past year; Yes, in the past year)
  10. Ever-use of pharmacological support (binary: yes; no)
  11. Ever-use or behavioural support (binary: yes; no)
  12. Season (four-level factor: spring; summer; autumn; winter)
- Within subject/time-varying
  1. Study day (numeric)
  2. Excited (quasi-numeric: 11-point Likert scale)
  3. Calm (quasi-numeric: 11-point Likert scale)
  4. Enthusiastic (quasi-numeric: 11-point Likert scale)
  5. Contented (quasi-numeric: 11-point Likert scale)
  6. Happy (quasi-numeric: 11-point Likert scale)
  7. Irritable (quasi-numeric: 11-point Likert scale)
  8. Bored (quasi-numeric: 11-point Likert scale)
  9. Anxious (quasi-numeric: 11-point Likert scale)
  10. Stressed (quasi-numeric: 11-point Likert scale)
  11. Sad (quasi-numeric: 11-point Likert scale)
  12. Pain (quasi-numeric: 11-point Likert scale)
  13. Momentary motivation (quasi-numeric: 11-point Likert scale)
  14. Craving (quasi-numeric: 11-point Likert scale)
  15. Self-efficacy (quasi-numeric: 11-point Likert scale)
  16. Cigarette availability (three level factor: Available with difficulty; Easily available; Not available)
  17. Past-hour nicotine use (binary: yes; no)
  18. Past-hour alcohol consumption (binary: yes; no)
  19. Past-hour caffeine consumption (binary: yes; no)
  20. Location: home (binary: yes; no)
  21. Location: school or work (binary: yes; no)
  22. Location: outside (binary: yes; no)
  23. Location: restaurant, café, bar (binary: yes; no)
  24. Location: public place (binary: yes; no)
  25. Location: public transport (binary: yes; no)
  26. Location: private vehicle (binary: yes; no)
  27. Location: other (binary: yes; no)
  28. Activity: eating or drinking (binary: yes; no)
  29. Activity: watching TV (binary: yes; no)
  30. Activity: listening to music (binary: yes; no)
  31. Activity: reading (binary: yes; no)
  32. Activity: working or studying (binary: yes; no)
  33. Activity: walking or exercising (binary: yes; no)
  34. Activity: socialising (binary: yes; no)
  35. Activity: scrolling on social media (binary: yes; no)
  36. Activity: relaxing (binary: yes; no)
  37. Activity: doing chores (binary: yes; no)
  38. Activity: other (binary: yes; no)
  39. Social context: alone (binary: yes; no)
  40. Social context: with partner or spouse (binary: yes; no)
  41. Social context: with relative(s) (binary: yes; no)
  42. Social context: other (binary: yes; no)
  43. Event-contingent prompt (binary: yes; no)
  44. Lapse prior (between current EMA and the one directly before; binary: yes; no)
  45. Time of day (four-level factor: morning; midday; evening; night)

### Models with feature selection

#### Number of features selected for each model

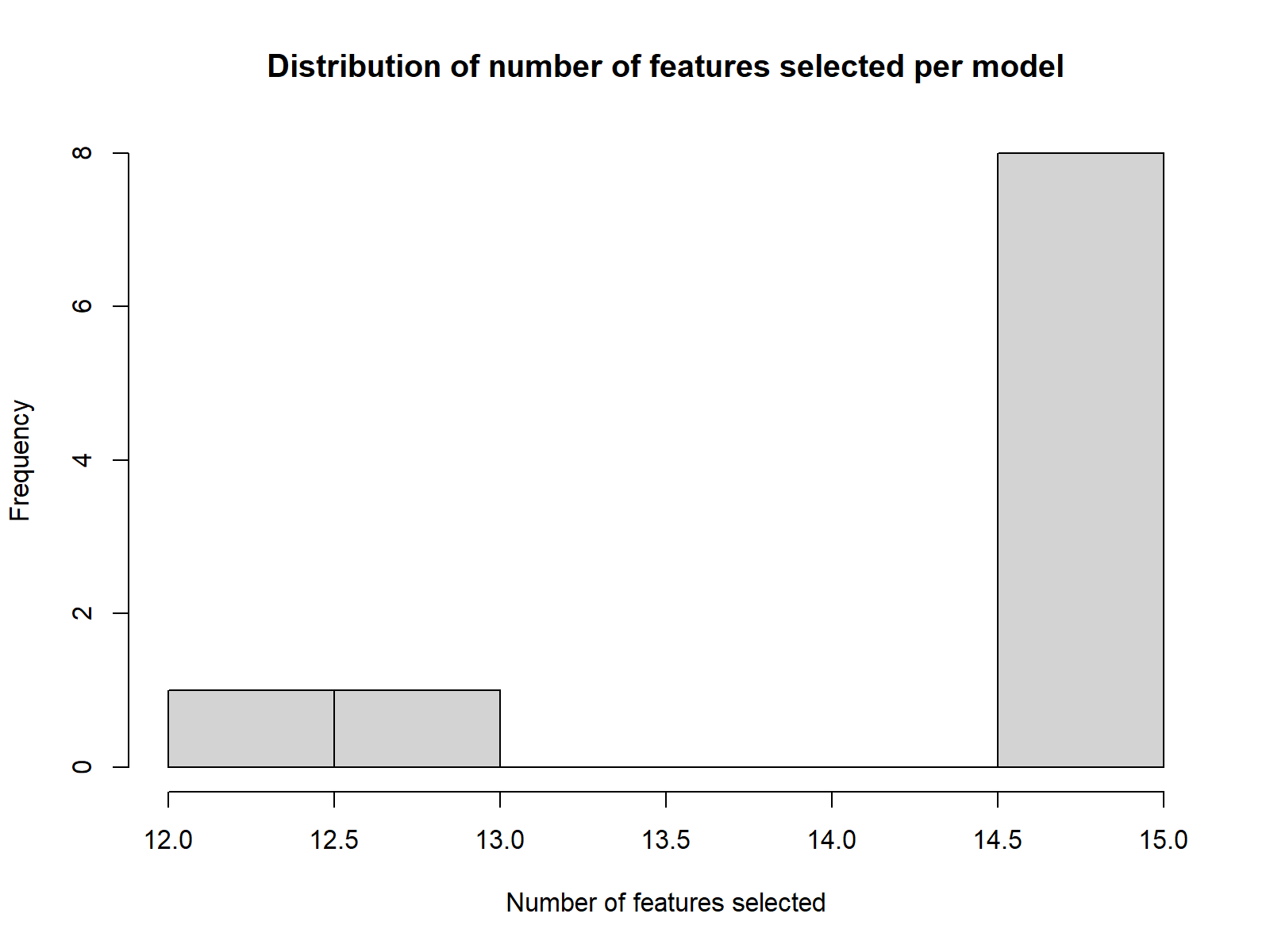

| Outcome | Cravings | | | | | Lapses | | | | |
| --- | --- | --- | --- | --- | --- | --- | --- | --- | --- | --- |
| Prompts per day | 16 | 6 | 5 | 4 | 3 | 16 | 6 | 5 | 4 | 3 |
| Number of features selected | 15 | 15 | 15 | 15 | 15 | 13 | 12 | 15 | 15 | 15 |

#### Number of models each feature is selected in

| **Feature** | **Between- or within-subject?** | **Measurement scale** | **Number of models selected in** | | |
| --- | --- | --- | --- | --- | --- |
|  |  |  | **Overall (out of 10)** | **Craving prediction (out of 5)** | **Lapse prediction (out of 5)** |
| Age | Between subject (time-invariant) | Numeric | 10 | 5 | 5 |
| Cigarettes per day | Between subject (time-invariant) | Numeric | 10 | 5 | 5 |
| Occupation type | Between subject (time-invariant) | Three-level factor | 10 | 5 | 5 |
| Motivation to stop | Between subject (time-invariant) | Six-level factor | 10 | 5 | 5 |
| Season | Between subject (time-invariant) | Four-level factor | 10 | 5 | 5 |
| Irritable | Within-subject (time-varying) | Quasi-numeric (11-point Likert scale) | 9 | 5 | 4 |
| Momentary motivation | Within-subject (time-varying) | Quasi-numeric (11-point Likert scale) | 9 | 4 | 5 |
| Time to first cigarette | Between subject (time-invariant) | Four-level factor | 8 | 3 | 5 |
| Self-efficacy | Within-subject (time-varying) | Quasi-numeric (11-point Likert scale) | 7 | 2 | 5 |
| Stressed | Within-subject (time-varying) | Quasi-numeric (11-point Likert scale) | 7 | 5 | 2 |
| Cigarette availability | Within-subject (time-varying) | Three-level factor | 6 | 1 | 5 |
| Craving | Within-subject (time-varying) | Quasi-numeric (11-point Likert scale) | 6 | 5 | 1 |
| Bored | Within-subject (time-varying) | Quasi-numeric (11-point Likert scale) | 5 | 5 | 0 |
| Enthusiastic | Within-subject (time-varying) | Quasi-numeric (11-point Likert scale) | 5 | 4 | 1 |
| Lapse prior | Within-subject (time-varying) | Binary | 5 | 0 | 5 |
| Pain | Within-subject (time-varying) | Quasi-numeric (11-point Likert scale) | 5 | 5 | 0 |
| Past quit attempt | Between-subject (time-invariant) | Three-level factor | 5 | 0 | 5 |
| Happy | Within-subject (time-varying) | Quasi-numeric (11-point Likert scale) | 4 | 3 | 1 |
| Ever-use of behavioural support | Between-subject (time-invariant) | Binary | 3 | 3 | 0 |
| Ever-use of pharmacological support | Between-subject (time-invariant) | Binary | 3 | 0 | 3 |
| Contented | Within-subject (time-varying) | Quasi-numeric (11-point Likert scale) | 2 | 1 | 1 |
| Ethnicity | Between-subject (time-invariant) | Five-level factor | 2 | 0 | 2 |
| Anxious | Within-subject (time-varying) | Quasi-numeric (11-point Likert scale) | 1 | 1 | 0 |
| Calm | Within-subject (time-varying) | Quasi-numeric (11-point Likert scale) | 1 | 1 | 0 |
| Excited | Within-subject (time-varying) | Quasi-numeric (11-point Likert scale) | 1 | 1 | 0 |
| Study-day | Within-subject (time-varying) | Numeric | 1 | 1 | 0 |

#### Actual features selected for each model

Underlined = time-varying/within-subject

|  | Cravings | Lapses |
| --- | --- | --- |
| 16 prompts per day | 1. Craving 2. Age 3. Study day 4. Cigarettes per day 5. Irritable 6. Season 7. Pain 8. Self-efficacy 9. Bored 10. Stressed 11. Motivation to stop 12. Momentary motivation 13. Occupation type 14. Cigarette availability 15. Enthusiastic | 1. Age 2. Momentary motivation 3. Motivation to stop 4. Lapse prior 5. Self-efficacy 6. Time to first cigarette 7. Occupation type 8. Craving 9. Cigarette availability 10. Season 11. Cigarettes per day 12. Past quit attempt 13. Ever-use of pharmacological support |
| 6 prompts per day | 1. Craving 2. Age 3. Pain 4. Cigarettes per day 5. Stressed 6. Motivation to stop 7. Bored 8. Season 9. Self-efficacy 10. Occupation type 11. Momentary motivation 12. Time to first cigarette 13. Happy 14. Irritable 15. Enthusiastic | 1. Age 2. Motivation to stop 3. Momentary motivation 4. Lapse prior 5. Self-efficacy 6. Cigarette availability 7. Season 8. Occupation type 9. Time to first cigarette 10. Past quit attempt 11. Cigarettes per day 12. Irritable |
| 5 prompts per day | 1. Craving 2. Age 3. Cigarettes per day 4. Pain 5. Motivation to stop 6. Season 7. Bored 8. Stressed 9. Irritable 10. Occupation type 11. Anxious 12. Contented 13. Time to first cigarette 14. Happy 15. Ever-use of behavioural support | 1. Age 2. Motivation to stop 3. Momentary motivation 4. Lapse prior 5. Self-efficacy 6. Season 7. Occupation type 8. Cigarettes per day 9. Time to first cigarette 10. Past quit attempt 11. Cigarette availability 12. Ever-use of pharmacological support 13. Ethnicity 14. Happy 15. Irritable |
| 4 prompts per day | 1. Craving 2. Age 3. Pain 4. Cigarettes per day 5. Motivation to stop 6. Stressed 7. Season 8. Bored 9. Occupation type 10. Irritable 11. Ever-use of behavioural support 12. Momentary motivation 13. Enthusiastic 14. Calm 15. Excited | 1. Motivation to stop 2. Lapse prior 3. Age 4. Cigarette availability 5. Season 6. Self-efficacy 7. Occupation type 8. Momentary motivation 9. Cigarettes per day 10. Past quit attempt 11. Time to first cigarette 12. Ever-use of pharmacological support 13. Irritable 14. Stressed 15. Contented |
| 3 prompts per day | 1. Craving 2. Age 3. Motivation to stop 4. Cigarettes per day 5. Season 6. Pain 7. Bored 8. Time to first cigarette 9. Happy 10. Occupation type 11. Irritable 12. Ever-use of behavioural support 13. Stressed 14. Momentary motivation 15. Enthusiastic | 1. Lapse prior 2. Motivation to stop 3. Cigarette availability 4. Self-efficacy 5. Age 6. Momentary motivation 7. Season 8. Cigarettes per day 9. Occupation type 10. Past quit attempt 11. Time to first cigarette 12. Ethnicity 13. Stressed 14. Enthusiastic 15. Irritable |
