## Supplementary material for "Optimising supervised machine learning algorithms predicting cigarette cravings and lapses for a smoking cessation just-in-time adaptive intervention (JITAI)": S3 Appendix Description of observations lapsers only

Supplementary Table 1 Description of observations among the participants with at least one lapse

| Prompts per day | EMAs | | | | Lapses | | | | High cravings | | | |
| --- | --- | --- | --- | --- | --- | --- | --- | --- | --- | --- | --- | --- |
|  | Number | | | Proportion event contingent | Overall proportion | Across individuals | | | Overall proportion | Across individuals | | |
|  | Total | Event contingent | Signal contingent |  |  | Median proportion | 25th percentile proportion | 75th percentile proportion |  | Median proportion | 25th percentile proportion | 75th percentile proportion |
| 16 | 4,044 | 44 | 4,000 | 1.1% | 10.2% | 6.2% | 1.2% | 13.4% | 34.6% | 29.6% | 13.1% | 44.1% |
| 6 | 1,544 | 44 | 1,500 | 2.8% | 18.4% | 11.8% | 3.3% | 26.7% | 35.1% | 29.5% | 11.3% | 45.9% |
| 5 | 1,294 | 44 | 1,250 | 3.4% | 20.6% | 13.5% | 2.0% | 33.3% | 33.9% | 26.9% | 10.0% | 47.1% |
| 4 | 1,003 | 43 | 960 | 4.3% | 24.1% | 15.5% | 4.3% | 39.0% | 35.1% | 27.2% | 13.6% | 42.3% |
| 3 | 763 | 43 | 720 | 5.6% | 29.1% | 19.9% | 5.7% | 44.6% | 35.3% | 28.6% | 15.4% | 44.6% |
