## Supplementary material for "Optimising supervised machine learning algorithms predicting cigarette cravings and lapses for a smoking cessation just-in-time adaptive intervention (JITAI)": S4 Appendix F1-Score performance summaries

### 1. Descriptive performance summaries: F1-Score

#### 1.1. By specific predictors

##### 1.1.1. By prompts per day

F1-Score by prompts per day

| **Prompts per day** | **n** | **Median** | **Mean** | **Standard deviation** | **25th percentile** | **75th percentile** | **Minimum** | **Maximum** |
| --- | --- | --- | --- | --- | --- | --- | --- | --- |
| 16 | 450 | 0.383 | 0.410 | 0.322 | 0.111 | 0.683 | 0.000 | 0.997 |
| 6 | 436 | 0.444 | 0.434 | 0.298 | 0.222 | 0.633 | 0.000 | 0.992 |
| 5 | 430 | 0.400 | 0.394 | 0.307 | 0.101 | 0.611 | 0.000 | 0.990 |
| 4 | 446 | 0.400 | 0.398 | 0.315 | 0.000 | 0.645 | 0.000 | 0.987 |
| 3 | 442 | 0.429 | 0.411 | 0.322 | 0.000 | 0.650 | 0.000 | 0.984 |

##### 1.1.2. By prompts per day - proportion below threshold

F1-Score by prompts per day (proportion below acceptable threshold of 0.5)

| **Prompts per day** | **n** | **N below threshold** | **Proportion below threshold** |
| --- | --- | --- | --- |
| 16 | 450 | 270 | 0.600 |
| 6 | 436 | 257 | 0.589 |
| 5 | 430 | 262 | 0.609 |
| 4 | 446 | 263 | 0.590 |
| 3 | 442 | 247 | 0.559 |

F1-Score by prompts per day × outcome (outcome: lapses)

| **Prompts per day** | **Outcome** | **n** | **Median** | **Mean** | **Standard deviation** | **25th percentile** | **75th percentile** | **Minimum** | **Maximum** |
| --- | --- | --- | --- | --- | --- | --- | --- | --- | --- |
| 16 | Lapses | 158 | 0.254 | 0.308 | 0.274 | 0.081 | 0.500 | 0.000 | 0.857 |
| 6 | Lapses | 156 | 0.440 | 0.400 | 0.262 | 0.222 | 0.587 | 0.000 | 0.945 |
| 5 | Lapses | 156 | 0.444 | 0.397 | 0.277 | 0.143 | 0.573 | 0.000 | 0.955 |
| 4 | Lapses | 156 | 0.475 | 0.432 | 0.285 | 0.218 | 0.640 | 0.000 | 0.986 |
| 3 | Lapses | 154 | 0.588 | 0.508 | 0.281 | 0.353 | 0.667 | 0.000 | 0.962 |

F1-Score by prompts per day × outcome (outcome: cravings)

| **Prompts per day** | **Outcome** | **n** | **Median** | **Mean** | **Standard deviation** | **25th percentile** | **75th percentile** | **Minimum** | **Maximum** |
| --- | --- | --- | --- | --- | --- | --- | --- | --- | --- |
| 16 | Cravings | 292 | 0.470 | 0.465 | 0.332 | 0.141 | 0.745 | 0.000 | 0.997 |
| 6 | Cravings | 280 | 0.447 | 0.453 | 0.315 | 0.211 | 0.658 | 0.000 | 0.992 |
| 5 | Cravings | 274 | 0.368 | 0.393 | 0.323 | 0.000 | 0.624 | 0.000 | 0.990 |
| 4 | Cravings | 290 | 0.361 | 0.379 | 0.329 | 0.000 | 0.647 | 0.000 | 0.987 |
| 3 | Cravings | 288 | 0.333 | 0.358 | 0.331 | 0.000 | 0.600 | 0.000 | 0.984 |

F1-Score by prompts per day × outcome (proportion below acceptable threshold of 0.5) (outcome: lapses)

| **Prompts per day** | **Outcome** | **n** | **N below threshold** | **Proportion below threshold** |
| --- | --- | --- | --- | --- |
| 16 | Lapses | 158 | 116 | 0.734 |
| 6 | Lapses | 156 | 99 | 0.635 |
| 5 | Lapses | 156 | 86 | 0.551 |
| 4 | Lapses | 156 | 79 | 0.506 |
| 3 | Lapses | 154 | 61 | 0.396 |

F1-Score by prompts per day × outcome (proportion below acceptable threshold of 0.5) (outcome: cravings)

| **Prompts per day** | **Outcome** | **n** | **N below threshold** | **Proportion below threshold** |
| --- | --- | --- | --- | --- |
| 16 | Cravings | 292 | 154 | 0.527 |
| 6 | Cravings | 280 | 158 | 0.564 |
| 5 | Cravings | 274 | 176 | 0.642 |
| 4 | Cravings | 290 | 184 | 0.634 |
| 3 | Cravings | 288 | 186 | 0.646 |

##### 1.1.3. By feature selection

F1-Score by feature selection

| **Feature selection** | **n** | **Median** | **Mean** | **Standard deviation** | **25th percentile** | **75th percentile** | **Minimum** | **Maximum** |
| --- | --- | --- | --- | --- | --- | --- | --- | --- |
| All features | 1,102 | 0.420 | 0.411 | 0.307 | 0.144 | 0.636 | 0.000 | 0.997 |
| Selected features | 1,102 | 0.400 | 0.407 | 0.318 | 0.080 | 0.645 | 0.000 | 0.997 |

##### 1.1.4. By feature selection - proportion below threshold

F1-Score by feature selection (proportion below acceptable threshold of 0.5)

| **Feature selection** | **n** | **N below threshold** | **Proportion below threshold** |
| --- | --- | --- | --- |
| All features | 1,102 | 643 | 0.583 |
| Selected features | 1,102 | 656 | 0.595 |

F1-Score by feature selection × outcome (outcome: lapses)

| **Feature selection** | **Outcome** | **n** | **Median** | **Mean** | **Standard deviation** | **25th percentile** | **75th percentile** | **Minimum** | **Maximum** |
| --- | --- | --- | --- | --- | --- | --- | --- | --- | --- |
| All features | Lapses | 390 | 0.435 | 0.404 | 0.278 | 0.188 | 0.620 | 0.000 | 0.986 |
| Selected features | Lapses | 390 | 0.441 | 0.413 | 0.287 | 0.168 | 0.632 | 0.000 | 0.986 |

F1-Score by feature selection × outcome (outcome: cravings)

| **Feature selection** | **Outcome** | **n** | **Median** | **Mean** | **Standard deviation** | **25th percentile** | **75th percentile** | **Minimum** | **Maximum** |
| --- | --- | --- | --- | --- | --- | --- | --- | --- | --- |
| All features | Cravings | 712 | 0.410 | 0.415 | 0.322 | 0.117 | 0.646 | 0.000 | 0.997 |
| Selected features | Cravings | 712 | 0.400 | 0.404 | 0.334 | 0.000 | 0.650 | 0.000 | 0.997 |

F1-Score by feature selection × outcome (proportion below acceptable threshold of 0.5) (outcome: lapses)

| **Feature selection** | **Outcome** | **n** | **N below threshold** | **Proportion below threshold** |
| --- | --- | --- | --- | --- |
| All features | Lapses | 390 | 224 | 0.574 |
| Selected features | Lapses | 390 | 217 | 0.556 |

F1-Score by feature selection × outcome (proportion below acceptable threshold of 0.5) (outcome: cravings)

| **Feature selection** | **Outcome** | **n** | **N below threshold** | **Proportion below threshold** |
| --- | --- | --- | --- | --- |
| All features | Cravings | 712 | 419 | 0.588 |
| Selected features | Cravings | 712 | 439 | 0.617 |

##### 1.1.5. By share of own data in training set

F1-Score by share of own data in training set

| **Share of own data in training set** | **n** | **Median** | **Mean** | **Standard deviation** | **25th percentile** | **75th percentile** | **Minimum** | **Maximum** |
| --- | --- | --- | --- | --- | --- | --- | --- | --- |
| None | 602 | 0.388 | 0.381 | 0.304 | 0.069 | 0.624 | 0.000 | 0.997 |
| 10% | 554 | 0.412 | 0.402 | 0.304 | 0.139 | 0.619 | 0.000 | 0.997 |
| 20% | 524 | 0.415 | 0.419 | 0.314 | 0.154 | 0.643 | 0.000 | 0.996 |
| 30% | 524 | 0.455 | 0.439 | 0.328 | 0.117 | 0.698 | 0.000 | 0.996 |

##### 1.1.6. By share of own data in training set - proportion below threshold

F1-Score by share of own data in training set (proportion below acceptable threshold of 0.5)

| **Share of own data in training set** | **n** | **N below threshold** | **Proportion below threshold** |
| --- | --- | --- | --- |
| None | 602 | 379 | 0.630 |
| 10% | 554 | 334 | 0.603 |
| 20% | 524 | 307 | 0.586 |
| 30% | 524 | 279 | 0.532 |

F1-Score by share of own data in training set × outcome (outcome: lapses)

| **Share of own data in training set** | **Outcome** | **n** | **Median** | **Mean** | **Standard deviation** | **25th percentile** | **75th percentile** | **Minimum** | **Maximum** |
| --- | --- | --- | --- | --- | --- | --- | --- | --- | --- |
| None | Lapses | 242 | 0.286 | 0.319 | 0.286 | 0.000 | 0.571 | 0.000 | 0.963 |
| 10% | Lapses | 198 | 0.400 | 0.384 | 0.282 | 0.155 | 0.607 | 0.000 | 0.986 |
| 20% | Lapses | 170 | 0.500 | 0.484 | 0.238 | 0.308 | 0.650 | 0.000 | 0.984 |
| 30% | Lapses | 170 | 0.542 | 0.490 | 0.278 | 0.329 | 0.667 | 0.000 | 0.982 |

F1-Score by share of own data in training set × outcome (outcome: cravings)

| **Share of own data in training set** | **Outcome** | **n** | **Median** | **Mean** | **Standard deviation** | **25th percentile** | **75th percentile** | **Minimum** | **Maximum** |
| --- | --- | --- | --- | --- | --- | --- | --- | --- | --- |
| None | Cravings | 360 | 0.424 | 0.422 | 0.309 | 0.143 | 0.649 | 0.000 | 0.997 |
| 10% | Cravings | 356 | 0.419 | 0.412 | 0.315 | 0.123 | 0.633 | 0.000 | 0.997 |
| 20% | Cravings | 354 | 0.333 | 0.388 | 0.341 | 0.000 | 0.640 | 0.000 | 0.996 |
| 30% | Cravings | 354 | 0.400 | 0.415 | 0.348 | 0.000 | 0.703 | 0.000 | 0.996 |

F1-Score by share of own data in training set × outcome (proportion below acceptable threshold of 0.5) (outcome: lapses)

| **Share of own data in training set** | **Outcome** | **n** | **N below threshold** | **Proportion below threshold** |
| --- | --- | --- | --- | --- |
| None | Lapses | 242 | 170 | 0.702 |
| 10% | Lapses | 198 | 120 | 0.606 |
| 20% | Lapses | 170 | 81 | 0.476 |
| 30% | Lapses | 170 | 70 | 0.412 |

F1-Score by share of own data in training set × outcome (proportion below acceptable threshold of 0.5) (outcome: cravings)

| **Share of own data in training set** | **Outcome** | **n** | **N below threshold** | **Proportion below threshold** |
| --- | --- | --- | --- | --- |
| None | Cravings | 360 | 209 | 0.581 |
| 10% | Cravings | 356 | 214 | 0.601 |
| 20% | Cravings | 354 | 226 | 0.638 |
| 30% | Cravings | 354 | 209 | 0.590 |

##### 1.1.7. By outcome

F1-Score by outcome

| **Outcome** | **n** | **Median** | **Mean** | **Standard deviation** | **25th percentile** | **75th percentile** | **Minimum** | **Maximum** |
| --- | --- | --- | --- | --- | --- | --- | --- | --- |
| Lapses | 780 | 0.436 | 0.409 | 0.283 | 0.180 | 0.625 | 0.000 | 0.986 |
| Cravings | 1,424 | 0.400 | 0.410 | 0.328 | 0.048 | 0.649 | 0.000 | 0.997 |

##### 1.1.8. By outcome - proportion below threshold

F1-Score by outcome (proportion below acceptable threshold of 0.5)

| **Outcome** | **n** | **N below threshold** | **Proportion below threshold** |
| --- | --- | --- | --- |
| Lapses | 780 | 441 | 0.565 |
| Cravings | 1,424 | 858 | 0.603 |

#### 1.2. By specification

##### 1.2.1. By specification

F1-Score by specification (prompts per day × use of feature selection × share of own data in training set × outcome)

| **Prompts per day** | **Feature selection** | **Share of own data in training set** | **Outcome** | **n** | **Median** | **Mean** | **Standard deviation** | **25th percentile** | **75th percentile** | **Minimum** | **Maximum** |
| --- | --- | --- | --- | --- | --- | --- | --- | --- | --- | --- | --- |
| 16 | All features | None | Lapses | 25 | 0.161 | 0.227 | 0.250 | 0.000 | 0.348 | 0.000 | 0.833 |
| 16 | All features | None | Cravings | 37 | 0.493 | 0.482 | 0.303 | 0.204 | 0.704 | 0.000 | 0.997 |
| 16 | All features | 10% | Lapses | 20 | 0.221 | 0.267 | 0.231 | 0.122 | 0.388 | 0.000 | 0.837 |
| 16 | All features | 10% | Cravings | 37 | 0.456 | 0.461 | 0.315 | 0.222 | 0.644 | 0.000 | 0.997 |
| 16 | All features | 20% | Lapses | 17 | 0.281 | 0.371 | 0.271 | 0.187 | 0.533 | 0.000 | 0.842 |
| 16 | All features | 20% | Cravings | 36 | 0.476 | 0.461 | 0.336 | 0.171 | 0.712 | 0.000 | 0.996 |
| 16 | All features | 30% | Lapses | 17 | 0.327 | 0.357 | 0.303 | 0.083 | 0.533 | 0.000 | 0.857 |
| 16 | All features | 30% | Cravings | 36 | 0.500 | 0.481 | 0.349 | 0.146 | 0.774 | 0.000 | 0.996 |
| 16 | Selected features | None | Lapses | 25 | 0.171 | 0.267 | 0.274 | 0.000 | 0.424 | 0.000 | 0.833 |
| 16 | Selected features | None | Cravings | 37 | 0.533 | 0.484 | 0.324 | 0.222 | 0.716 | 0.000 | 0.994 |
| 16 | Selected features | 10% | Lapses | 20 | 0.255 | 0.315 | 0.283 | 0.115 | 0.516 | 0.000 | 0.834 |
| 16 | Selected features | 10% | Cravings | 37 | 0.475 | 0.450 | 0.346 | 0.095 | 0.737 | 0.000 | 0.997 |
| 16 | Selected features | 20% | Lapses | 17 | 0.270 | 0.359 | 0.280 | 0.202 | 0.600 | 0.000 | 0.842 |
| 16 | Selected features | 20% | Cravings | 36 | 0.450 | 0.454 | 0.351 | 0.079 | 0.728 | 0.000 | 0.996 |
| 16 | Selected features | 30% | Lapses | 17 | 0.333 | 0.370 | 0.314 | 0.138 | 0.588 | 0.000 | 0.857 |
| 16 | Selected features | 30% | Cravings | 36 | 0.419 | 0.445 | 0.361 | 0.068 | 0.774 | 0.000 | 0.996 |
| 6 | All features | None | Lapses | 24 | 0.273 | 0.310 | 0.279 | 0.000 | 0.450 | 0.000 | 0.940 |
| 6 | All features | None | Cravings | 35 | 0.494 | 0.472 | 0.285 | 0.310 | 0.627 | 0.000 | 0.992 |
| 6 | All features | 10% | Lapses | 20 | 0.389 | 0.368 | 0.264 | 0.195 | 0.563 | 0.000 | 0.941 |
| 6 | All features | 10% | Cravings | 35 | 0.467 | 0.457 | 0.304 | 0.221 | 0.662 | 0.000 | 0.991 |
| 6 | All features | 20% | Lapses | 17 | 0.444 | 0.464 | 0.221 | 0.333 | 0.600 | 0.000 | 0.945 |
| 6 | All features | 20% | Cravings | 35 | 0.444 | 0.438 | 0.334 | 0.167 | 0.655 | 0.000 | 0.989 |
| 6 | All features | 30% | Lapses | 17 | 0.500 | 0.468 | 0.241 | 0.381 | 0.566 | 0.000 | 0.937 |
| 6 | All features | 30% | Cravings | 35 | 0.449 | 0.463 | 0.324 | 0.258 | 0.666 | 0.000 | 0.988 |
| 6 | Selected features | None | Lapses | 24 | 0.319 | 0.321 | 0.281 | 0.000 | 0.541 | 0.000 | 0.901 |
| 6 | Selected features | None | Cravings | 35 | 0.483 | 0.487 | 0.305 | 0.341 | 0.674 | 0.000 | 0.992 |
| 6 | Selected features | 10% | Lapses | 20 | 0.435 | 0.390 | 0.280 | 0.188 | 0.598 | 0.000 | 0.941 |
| 6 | Selected features | 10% | Cravings | 35 | 0.444 | 0.456 | 0.311 | 0.276 | 0.650 | 0.000 | 0.991 |
| 6 | Selected features | 20% | Lapses | 17 | 0.476 | 0.457 | 0.227 | 0.327 | 0.600 | 0.000 | 0.945 |
| 6 | Selected features | 20% | Cravings | 35 | 0.308 | 0.410 | 0.343 | 0.144 | 0.648 | 0.000 | 0.989 |
| 6 | Selected features | 30% | Lapses | 17 | 0.508 | 0.499 | 0.240 | 0.444 | 0.667 | 0.000 | 0.937 |
| 6 | Selected features | 30% | Cravings | 35 | 0.415 | 0.438 | 0.332 | 0.229 | 0.669 | 0.000 | 0.988 |
| 5 | All features | None | Lapses | 24 | 0.219 | 0.300 | 0.306 | 0.000 | 0.588 | 0.000 | 0.938 |
| 5 | All features | None | Cravings | 35 | 0.381 | 0.403 | 0.291 | 0.194 | 0.635 | 0.000 | 0.990 |
| 5 | All features | 10% | Lapses | 20 | 0.296 | 0.322 | 0.280 | 0.000 | 0.512 | 0.000 | 0.953 |
| 5 | All features | 10% | Cravings | 34 | 0.395 | 0.395 | 0.301 | 0.223 | 0.583 | 0.000 | 0.989 |
| 5 | All features | 20% | Lapses | 17 | 0.500 | 0.493 | 0.202 | 0.378 | 0.571 | 0.190 | 0.947 |
| 5 | All features | 20% | Cravings | 34 | 0.333 | 0.397 | 0.336 | 0.042 | 0.649 | 0.000 | 0.987 |
| 5 | All features | 30% | Lapses | 17 | 0.560 | 0.500 | 0.216 | 0.400 | 0.600 | 0.087 | 0.955 |
| 5 | All features | 30% | Cravings | 34 | 0.325 | 0.377 | 0.351 | 0.000 | 0.622 | 0.000 | 0.986 |
| 5 | Selected features | None | Lapses | 24 | 0.288 | 0.315 | 0.282 | 0.071 | 0.497 | 0.000 | 0.917 |
| 5 | Selected features | None | Cravings | 35 | 0.375 | 0.392 | 0.323 | 0.083 | 0.634 | 0.000 | 0.990 |
| 5 | Selected features | 10% | Lapses | 20 | 0.382 | 0.351 | 0.293 | 0.000 | 0.568 | 0.000 | 0.953 |
| 5 | Selected features | 10% | Cravings | 34 | 0.404 | 0.393 | 0.318 | 0.091 | 0.564 | 0.000 | 0.989 |
| 5 | Selected features | 20% | Lapses | 17 | 0.562 | 0.492 | 0.262 | 0.267 | 0.689 | 0.000 | 0.947 |
| 5 | Selected features | 20% | Cravings | 34 | 0.333 | 0.385 | 0.347 | 0.000 | 0.591 | 0.000 | 0.987 |
| 5 | Selected features | 30% | Lapses | 17 | 0.549 | 0.502 | 0.262 | 0.353 | 0.727 | 0.000 | 0.955 |
| 5 | Selected features | 30% | Cravings | 34 | 0.374 | 0.398 | 0.346 | 0.000 | 0.624 | 0.000 | 0.986 |
| 4 | All features | None | Lapses | 24 | 0.268 | 0.332 | 0.307 | 0.000 | 0.658 | 0.000 | 0.892 |
| 4 | All features | None | Cravings | 37 | 0.435 | 0.402 | 0.314 | 0.105 | 0.643 | 0.000 | 0.987 |
| 4 | All features | 10% | Lapses | 20 | 0.414 | 0.396 | 0.279 | 0.198 | 0.582 | 0.000 | 0.986 |
| 4 | All features | 10% | Cravings | 36 | 0.400 | 0.393 | 0.313 | 0.115 | 0.570 | 0.000 | 0.986 |
| 4 | All features | 20% | Lapses | 17 | 0.556 | 0.522 | 0.197 | 0.400 | 0.609 | 0.207 | 0.984 |
| 4 | All features | 20% | Cravings | 36 | 0.258 | 0.342 | 0.329 | 0.000 | 0.617 | 0.000 | 0.984 |
| 4 | All features | 30% | Lapses | 17 | 0.556 | 0.471 | 0.297 | 0.308 | 0.634 | 0.000 | 0.982 |
| 4 | All features | 30% | Cravings | 36 | 0.422 | 0.412 | 0.347 | 0.000 | 0.682 | 0.000 | 0.985 |
| 4 | Selected features | None | Lapses | 24 | 0.333 | 0.368 | 0.300 | 0.000 | 0.611 | 0.000 | 0.963 |
| 4 | Selected features | None | Cravings | 37 | 0.414 | 0.404 | 0.324 | 0.000 | 0.654 | 0.000 | 0.987 |
| 4 | Selected features | 10% | Lapses | 20 | 0.509 | 0.433 | 0.303 | 0.213 | 0.628 | 0.000 | 0.986 |
| 4 | Selected features | 10% | Cravings | 36 | 0.369 | 0.375 | 0.322 | 0.000 | 0.560 | 0.000 | 0.986 |
| 4 | Selected features | 20% | Lapses | 17 | 0.500 | 0.492 | 0.244 | 0.375 | 0.609 | 0.000 | 0.984 |
| 4 | Selected features | 20% | Cravings | 36 | 0.333 | 0.336 | 0.345 | 0.000 | 0.576 | 0.000 | 0.987 |
| 4 | Selected features | 30% | Lapses | 17 | 0.600 | 0.518 | 0.301 | 0.333 | 0.750 | 0.000 | 0.982 |
| 4 | Selected features | 30% | Cravings | 36 | 0.305 | 0.367 | 0.360 | 0.000 | 0.709 | 0.000 | 0.985 |
| 3 | All features | None | Lapses | 24 | 0.385 | 0.386 | 0.305 | 0.000 | 0.612 | 0.000 | 0.951 |
| 3 | All features | None | Cravings | 36 | 0.312 | 0.355 | 0.303 | 0.000 | 0.577 | 0.000 | 0.983 |
| 3 | All features | 10% | Lapses | 19 | 0.571 | 0.509 | 0.266 | 0.434 | 0.667 | 0.000 | 0.941 |
| 3 | All features | 10% | Cravings | 36 | 0.388 | 0.378 | 0.307 | 0.060 | 0.582 | 0.000 | 0.981 |
| 3 | All features | 20% | Lapses | 17 | 0.640 | 0.608 | 0.164 | 0.500 | 0.667 | 0.286 | 0.957 |
| 3 | All features | 20% | Cravings | 36 | 0.235 | 0.334 | 0.350 | 0.000 | 0.588 | 0.000 | 0.984 |
| 3 | All features | 30% | Lapses | 17 | 0.667 | 0.656 | 0.218 | 0.600 | 0.800 | 0.000 | 0.950 |
| 3 | All features | 30% | Cravings | 36 | 0.319 | 0.392 | 0.364 | 0.000 | 0.711 | 0.000 | 0.976 |
| 3 | Selected features | None | Lapses | 24 | 0.408 | 0.368 | 0.292 | 0.000 | 0.600 | 0.000 | 0.857 |
| 3 | Selected features | None | Cravings | 36 | 0.369 | 0.342 | 0.303 | 0.000 | 0.538 | 0.000 | 0.966 |
| 3 | Selected features | 10% | Lapses | 19 | 0.571 | 0.507 | 0.296 | 0.330 | 0.711 | 0.000 | 0.962 |
| 3 | Selected features | 10% | Cravings | 36 | 0.348 | 0.361 | 0.330 | 0.000 | 0.579 | 0.000 | 0.981 |
| 3 | Selected features | 20% | Lapses | 17 | 0.588 | 0.579 | 0.233 | 0.483 | 0.750 | 0.000 | 0.957 |
| 3 | Selected features | 20% | Cravings | 36 | 0.245 | 0.324 | 0.344 | 0.000 | 0.561 | 0.000 | 0.984 |
| 3 | Selected features | 30% | Lapses | 17 | 0.632 | 0.559 | 0.311 | 0.429 | 0.800 | 0.000 | 0.950 |
| 3 | Selected features | 30% | Cravings | 36 | 0.354 | 0.381 | 0.366 | 0.000 | 0.611 | 0.000 | 0.980 |

##### 1.2.2. By specification - proportion below threshold

F1-Score by specification (prompts per day × use of feature selection × share of own data in training set × outcome) - proportion below acceptable threshold of 0.5

| **Prompts per day** | **Feature selection** | **Share of own data in training set** | **Outcome** | **n** | **N below threshold** | **Proportion below threshold** |
| --- | --- | --- | --- | --- | --- | --- |
| 16 | All features | None | Lapses | 25 | 21 | 0.840 |
| 16 | All features | None | Cravings | 37 | 19 | 0.514 |
| 16 | All features | 10% | Lapses | 20 | 16 | 0.800 |
| 16 | All features | 10% | Cravings | 37 | 20 | 0.541 |
| 16 | All features | 20% | Lapses | 17 | 11 | 0.647 |
| 16 | All features | 20% | Cravings | 36 | 19 | 0.528 |
| 16 | All features | 30% | Lapses | 17 | 11 | 0.647 |
| 16 | All features | 30% | Cravings | 36 | 17 | 0.472 |
| 16 | Selected features | None | Lapses | 25 | 19 | 0.760 |
| 16 | Selected features | None | Cravings | 37 | 18 | 0.486 |
| 16 | Selected features | 10% | Lapses | 20 | 15 | 0.750 |
| 16 | Selected features | 10% | Cravings | 37 | 20 | 0.541 |
| 16 | Selected features | 20% | Lapses | 17 | 12 | 0.706 |
| 16 | Selected features | 20% | Cravings | 36 | 20 | 0.556 |
| 16 | Selected features | 30% | Lapses | 17 | 11 | 0.647 |
| 16 | Selected features | 30% | Cravings | 36 | 21 | 0.583 |
| 6 | All features | None | Lapses | 24 | 19 | 0.792 |
| 6 | All features | None | Cravings | 35 | 18 | 0.514 |
| 6 | All features | 10% | Lapses | 20 | 14 | 0.700 |
| 6 | All features | 10% | Cravings | 35 | 18 | 0.514 |
| 6 | All features | 20% | Lapses | 17 | 11 | 0.647 |
| 6 | All features | 20% | Cravings | 35 | 20 | 0.571 |
| 6 | All features | 30% | Lapses | 17 | 8 | 0.471 |
| 6 | All features | 30% | Cravings | 35 | 20 | 0.571 |
| 6 | Selected features | None | Lapses | 24 | 17 | 0.708 |
| 6 | Selected features | None | Cravings | 35 | 18 | 0.514 |
| 6 | Selected features | 10% | Lapses | 20 | 13 | 0.650 |
| 6 | Selected features | 10% | Cravings | 35 | 18 | 0.514 |
| 6 | Selected features | 20% | Lapses | 17 | 10 | 0.588 |
| 6 | Selected features | 20% | Cravings | 35 | 23 | 0.657 |
| 6 | Selected features | 30% | Lapses | 17 | 7 | 0.412 |
| 6 | Selected features | 30% | Cravings | 35 | 23 | 0.657 |
| 5 | All features | None | Lapses | 24 | 17 | 0.708 |
| 5 | All features | None | Cravings | 35 | 22 | 0.629 |
| 5 | All features | 10% | Lapses | 20 | 13 | 0.650 |
| 5 | All features | 10% | Cravings | 34 | 23 | 0.676 |
| 5 | All features | 20% | Lapses | 17 | 8 | 0.471 |
| 5 | All features | 20% | Cravings | 34 | 22 | 0.647 |
| 5 | All features | 30% | Lapses | 17 | 6 | 0.353 |
| 5 | All features | 30% | Cravings | 34 | 21 | 0.618 |
| 5 | Selected features | None | Lapses | 24 | 18 | 0.750 |
| 5 | Selected features | None | Cravings | 35 | 22 | 0.629 |
| 5 | Selected features | 10% | Lapses | 20 | 12 | 0.600 |
| 5 | Selected features | 10% | Cravings | 34 | 22 | 0.647 |
| 5 | Selected features | 20% | Lapses | 17 | 6 | 0.353 |
| 5 | Selected features | 20% | Cravings | 34 | 22 | 0.647 |
| 5 | Selected features | 30% | Lapses | 17 | 6 | 0.353 |
| 5 | Selected features | 30% | Cravings | 34 | 22 | 0.647 |
| 4 | All features | None | Lapses | 24 | 16 | 0.667 |
| 4 | All features | None | Cravings | 37 | 21 | 0.568 |
| 4 | All features | 10% | Lapses | 20 | 11 | 0.550 |
| 4 | All features | 10% | Cravings | 36 | 23 | 0.639 |
| 4 | All features | 20% | Lapses | 17 | 7 | 0.412 |
| 4 | All features | 20% | Cravings | 36 | 25 | 0.694 |
| 4 | All features | 30% | Lapses | 17 | 8 | 0.471 |
| 4 | All features | 30% | Cravings | 36 | 20 | 0.556 |
| 4 | Selected features | None | Lapses | 24 | 14 | 0.583 |
| 4 | Selected features | None | Cravings | 37 | 21 | 0.568 |
| 4 | Selected features | 10% | Lapses | 20 | 10 | 0.500 |
| 4 | Selected features | 10% | Cravings | 36 | 24 | 0.667 |
| 4 | Selected features | 20% | Lapses | 17 | 7 | 0.412 |
| 4 | Selected features | 20% | Cravings | 36 | 26 | 0.722 |
| 4 | Selected features | 30% | Lapses | 17 | 6 | 0.353 |
| 4 | Selected features | 30% | Cravings | 36 | 24 | 0.667 |
| 3 | All features | None | Lapses | 24 | 14 | 0.583 |
| 3 | All features | None | Cravings | 36 | 24 | 0.667 |
| 3 | All features | 10% | Lapses | 19 | 8 | 0.421 |
| 3 | All features | 10% | Cravings | 36 | 23 | 0.639 |
| 3 | All features | 20% | Lapses | 17 | 3 | 0.176 |
| 3 | All features | 20% | Cravings | 36 | 24 | 0.667 |
| 3 | All features | 30% | Lapses | 17 | 2 | 0.118 |
| 3 | All features | 30% | Cravings | 36 | 20 | 0.556 |
| 3 | Selected features | None | Lapses | 24 | 15 | 0.625 |
| 3 | Selected features | None | Cravings | 36 | 26 | 0.722 |
| 3 | Selected features | 10% | Lapses | 19 | 8 | 0.421 |
| 3 | Selected features | 10% | Cravings | 36 | 23 | 0.639 |
| 3 | Selected features | 20% | Lapses | 17 | 6 | 0.353 |
| 3 | Selected features | 20% | Cravings | 36 | 25 | 0.694 |
| 3 | Selected features | 30% | Lapses | 17 | 5 | 0.294 |
| 3 | Selected features | 30% | Cravings | 36 | 21 | 0.583 |

#### 1.3. By participant

##### 1.3.1. By participant (overall)

F1-Score by participant

| **Participant** | **n** | **Median** | **Mean** | **Standard deviation** | **25th percentile** | **75th percentile** | **Minimum** | **Maximum** |
| --- | --- | --- | --- | --- | --- | --- | --- | --- |
| p1 | 80 | 0.353 | 0.351 | 0.152 | 0.235 | 0.460 | 0.000 | 0.652 |
| p10 | 40 | 0.000 | 0.042 | 0.091 | 0.000 | 0.027 | 0.000 | 0.368 |
| p101 | 80 | 0.500 | 0.485 | 0.280 | 0.222 | 0.744 | 0.000 | 0.839 |
| p107 | 80 | 0.310 | 0.270 | 0.227 | 0.000 | 0.469 | 0.000 | 0.667 |
| p12 | 80 | 0.146 | 0.246 | 0.253 | 0.000 | 0.478 | 0.000 | 0.700 |
| p128 | 80 | 0.424 | 0.408 | 0.202 | 0.256 | 0.556 | 0.000 | 0.757 |
| p129 | 40 | 0.000 | 0.062 | 0.116 | 0.000 | 0.115 | 0.000 | 0.500 |
| p131 | 42 | 0.642 | 0.627 | 0.119 | 0.587 | 0.683 | 0.000 | 0.758 |
| p132 | 60 | 0.781 | 0.586 | 0.425 | 0.000 | 0.957 | 0.000 | 0.988 |
| p135 | 80 | 0.477 | 0.479 | 0.138 | 0.422 | 0.567 | 0.000 | 0.667 |
| p141 | 80 | 0.536 | 0.574 | 0.361 | 0.232 | 0.955 | 0.000 | 0.987 |
| p144 | 80 | 0.626 | 0.544 | 0.218 | 0.416 | 0.692 | 0.000 | 0.830 |
| p147 | 40 | 0.449 | 0.444 | 0.124 | 0.373 | 0.551 | 0.143 | 0.617 |
| p15 | 80 | 0.203 | 0.316 | 0.315 | 0.000 | 0.667 | 0.000 | 0.833 |
| p20 | 40 | 0.988 | 0.987 | 0.007 | 0.984 | 0.991 | 0.966 | 0.997 |
| p21 | 80 | 0.544 | 0.498 | 0.132 | 0.397 | 0.609 | 0.250 | 0.711 |
| p22 | 80 | 0.525 | 0.527 | 0.164 | 0.400 | 0.667 | 0.000 | 0.778 |
| p27 | 32 | 0.000 | 0.000 | 0.000 | 0.000 | 0.000 | 0.000 | 0.000 |
| p30 | 50 | 0.195 | 0.193 | 0.156 | 0.074 | 0.307 | 0.000 | 0.516 |
| p39 | 40 | 0.609 | 0.590 | 0.131 | 0.491 | 0.700 | 0.333 | 0.783 |
| p44 | 40 | 0.308 | 0.356 | 0.190 | 0.217 | 0.437 | 0.000 | 0.714 |
| p59 | 40 | 0.429 | 0.340 | 0.220 | 0.163 | 0.493 | 0.000 | 0.654 |
| p6 | 80 | 0.667 | 0.691 | 0.243 | 0.465 | 0.945 | 0.308 | 0.986 |
| p61 | 80 | 0.500 | 0.385 | 0.311 | 0.000 | 0.600 | 0.000 | 0.857 |
| p62 | 60 | 0.000 | 0.071 | 0.114 | 0.000 | 0.127 | 0.000 | 0.462 |
| p63 | 80 | 0.548 | 0.475 | 0.266 | 0.257 | 0.708 | 0.000 | 0.857 |
| p67 | 42 | 0.000 | 0.000 | 0.000 | 0.000 | 0.000 | 0.000 | 0.000 |
| p7 | 80 | 0.250 | 0.230 | 0.198 | 0.000 | 0.400 | 0.000 | 0.769 |
| p70 | 6 | 0.000 | 0.000 | 0.000 | 0.000 | 0.000 | 0.000 | 0.000 |
| p74 | 50 | 0.480 | 0.429 | 0.331 | 0.065 | 0.737 | 0.000 | 0.923 |
| p78 | 40 | 0.927 | 0.879 | 0.130 | 0.884 | 0.945 | 0.392 | 0.968 |
| p79 | 40 | 0.000 | 0.079 | 0.137 | 0.000 | 0.113 | 0.000 | 0.400 |
| p87 | 58 | 0.981 | 0.683 | 0.448 | 0.066 | 0.988 | 0.000 | 0.997 |
| p9 | 50 | 0.282 | 0.251 | 0.170 | 0.122 | 0.398 | 0.000 | 0.500 |
| p93 | 40 | 0.879 | 0.844 | 0.075 | 0.794 | 0.894 | 0.681 | 0.946 |
| p95 | 80 | 0.479 | 0.492 | 0.183 | 0.375 | 0.597 | 0.000 | 0.895 |
| p97 | 74 | 0.000 | 0.140 | 0.169 | 0.000 | 0.286 | 0.000 | 0.667 |

##### 1.3.2. By participant (overall) - proportion below threshold

F1-Score by participant (proportion below acceptable threshold of 0.5)

| **Participant** | **n** | **N below threshold** | **Proportion below threshold** |
| --- | --- | --- | --- |
| p1 | 80 | 67 | 0.838 |
| p10 | 40 | 40 | 1.000 |
| p101 | 80 | 38 | 0.475 |
| p107 | 80 | 63 | 0.787 |
| p12 | 80 | 62 | 0.775 |
| p128 | 80 | 49 | 0.613 |
| p129 | 40 | 39 | 0.975 |
| p131 | 42 | 1 | 0.024 |
| p132 | 60 | 20 | 0.333 |
| p135 | 80 | 41 | 0.512 |
| p141 | 80 | 37 | 0.463 |
| p144 | 80 | 27 | 0.338 |
| p147 | 40 | 24 | 0.600 |
| p15 | 80 | 50 | 0.625 |
| p20 | 40 | 0 | 0.000 |
| p21 | 80 | 36 | 0.450 |
| p22 | 80 | 29 | 0.362 |
| p27 | 32 | 32 | 1.000 |
| p30 | 50 | 49 | 0.980 |
| p39 | 40 | 10 | 0.250 |
| p44 | 40 | 31 | 0.775 |
| p59 | 40 | 30 | 0.750 |
| p6 | 80 | 24 | 0.300 |
| p61 | 80 | 37 | 0.463 |
| p62 | 60 | 60 | 1.000 |
| p63 | 80 | 35 | 0.438 |
| p67 | 42 | 42 | 1.000 |
| p7 | 80 | 73 | 0.912 |
| p70 | 6 | 6 | 1.000 |
| p74 | 50 | 25 | 0.500 |
| p78 | 40 | 1 | 0.025 |
| p79 | 40 | 40 | 1.000 |
| p87 | 58 | 18 | 0.310 |
| p9 | 50 | 48 | 0.960 |
| p93 | 40 | 0 | 0.000 |
| p95 | 80 | 42 | 0.525 |
| p97 | 74 | 73 | 0.986 |

F1-Score by participant × outcome (outcome: lapses)

| **Participant** | **Outcome** | **n** | **Median** | **Mean** | **Standard deviation** | **25th percentile** | **75th percentile** | **Minimum** | **Maximum** |
| --- | --- | --- | --- | --- | --- | --- | --- | --- | --- |
| p1 | Lapses | 40 | 0.429 | 0.413 | 0.160 | 0.324 | 0.542 | 0.000 | 0.652 |
| p101 | Lapses | 40 | 0.222 | 0.241 | 0.174 | 0.156 | 0.349 | 0.000 | 0.727 |
| p107 | Lapses | 40 | 0.000 | 0.136 | 0.214 | 0.000 | 0.238 | 0.000 | 0.667 |
| p12 | Lapses | 40 | 0.478 | 0.451 | 0.189 | 0.367 | 0.600 | 0.000 | 0.700 |
| p128 | Lapses | 40 | 0.494 | 0.459 | 0.204 | 0.276 | 0.616 | 0.071 | 0.757 |
| p131 | Lapses | 2 | 0.250 | 0.250 | 0.354 | 0.125 | 0.375 | 0.000 | 0.500 |
| p132 | Lapses | 20 | 0.000 | 0.000 | 0.000 | 0.000 | 0.000 | 0.000 | 0.000 |
| p135 | Lapses | 40 | 0.560 | 0.535 | 0.114 | 0.467 | 0.625 | 0.118 | 0.667 |
| p141 | Lapses | 40 | 0.229 | 0.250 | 0.169 | 0.140 | 0.353 | 0.000 | 0.571 |
| p144 | Lapses | 40 | 0.667 | 0.649 | 0.104 | 0.600 | 0.712 | 0.375 | 0.800 |
| p15 | Lapses | 40 | 0.667 | 0.590 | 0.200 | 0.516 | 0.735 | 0.128 | 0.833 |
| p21 | Lapses | 40 | 0.548 | 0.522 | 0.134 | 0.475 | 0.610 | 0.270 | 0.711 |
| p22 | Lapses | 40 | 0.500 | 0.499 | 0.168 | 0.398 | 0.667 | 0.000 | 0.778 |
| p30 | Lapses | 10 | 0.000 | 0.000 | 0.000 | 0.000 | 0.000 | 0.000 | 0.000 |
| p6 | Lapses | 40 | 0.945 | 0.921 | 0.064 | 0.899 | 0.956 | 0.714 | 0.986 |
| p61 | Lapses | 40 | 0.600 | 0.648 | 0.128 | 0.571 | 0.707 | 0.333 | 0.857 |
| p62 | Lapses | 20 | 0.000 | 0.000 | 0.000 | 0.000 | 0.000 | 0.000 | 0.000 |
| p63 | Lapses | 40 | 0.710 | 0.631 | 0.197 | 0.571 | 0.750 | 0.176 | 0.857 |
| p67 | Lapses | 10 | 0.000 | 0.000 | 0.000 | 0.000 | 0.000 | 0.000 | 0.000 |
| p7 | Lapses | 40 | 0.278 | 0.260 | 0.157 | 0.164 | 0.365 | 0.000 | 0.600 |
| p74 | Lapses | 10 | 0.000 | 0.000 | 0.000 | 0.000 | 0.000 | 0.000 | 0.000 |
| p87 | Lapses | 18 | 0.000 | 0.023 | 0.036 | 0.000 | 0.052 | 0.000 | 0.095 |
| p9 | Lapses | 10 | 0.000 | 0.000 | 0.000 | 0.000 | 0.000 | 0.000 | 0.000 |
| p95 | Lapses | 40 | 0.448 | 0.485 | 0.241 | 0.295 | 0.679 | 0.000 | 0.895 |
| p97 | Lapses | 40 | 0.276 | 0.258 | 0.149 | 0.165 | 0.373 | 0.000 | 0.667 |

F1-Score by participant × outcome (outcome: cravings)

| **Participant** | **Outcome** | **n** | **Median** | **Mean** | **Standard deviation** | **25th percentile** | **75th percentile** | **Minimum** | **Maximum** |
| --- | --- | --- | --- | --- | --- | --- | --- | --- | --- |
| p1 | Cravings | 40 | 0.310 | 0.290 | 0.115 | 0.234 | 0.356 | 0.000 | 0.480 |
| p10 | Cravings | 40 | 0.000 | 0.042 | 0.091 | 0.000 | 0.027 | 0.000 | 0.368 |
| p101 | Cravings | 40 | 0.745 | 0.729 | 0.082 | 0.718 | 0.777 | 0.429 | 0.839 |
| p107 | Cravings | 40 | 0.445 | 0.405 | 0.146 | 0.315 | 0.500 | 0.000 | 0.631 |
| p12 | Cravings | 40 | 0.000 | 0.040 | 0.084 | 0.000 | 0.021 | 0.000 | 0.400 |
| p128 | Cravings | 40 | 0.400 | 0.358 | 0.190 | 0.230 | 0.502 | 0.000 | 0.688 |
| p129 | Cravings | 40 | 0.000 | 0.062 | 0.116 | 0.000 | 0.115 | 0.000 | 0.500 |
| p131 | Cravings | 40 | 0.643 | 0.646 | 0.063 | 0.588 | 0.684 | 0.539 | 0.758 |
| p132 | Cravings | 40 | 0.879 | 0.880 | 0.090 | 0.782 | 0.972 | 0.761 | 0.988 |
| p135 | Cravings | 40 | 0.444 | 0.422 | 0.138 | 0.405 | 0.504 | 0.000 | 0.643 |
| p141 | Cravings | 40 | 0.957 | 0.898 | 0.143 | 0.904 | 0.973 | 0.356 | 0.987 |
| p144 | Cravings | 40 | 0.433 | 0.440 | 0.251 | 0.200 | 0.643 | 0.000 | 0.830 |
| p147 | Cravings | 40 | 0.449 | 0.444 | 0.124 | 0.373 | 0.551 | 0.143 | 0.617 |
| p15 | Cravings | 40 | 0.000 | 0.042 | 0.083 | 0.000 | 0.000 | 0.000 | 0.286 |
| p20 | Cravings | 40 | 0.988 | 0.987 | 0.007 | 0.984 | 0.991 | 0.966 | 0.997 |
| p21 | Cravings | 40 | 0.444 | 0.475 | 0.127 | 0.384 | 0.600 | 0.250 | 0.702 |
| p22 | Cravings | 40 | 0.612 | 0.555 | 0.157 | 0.443 | 0.667 | 0.222 | 0.769 |
| p27 | Cravings | 32 | 0.000 | 0.000 | 0.000 | 0.000 | 0.000 | 0.000 | 0.000 |
| p30 | Cravings | 40 | 0.230 | 0.241 | 0.136 | 0.129 | 0.345 | 0.000 | 0.516 |
| p39 | Cravings | 40 | 0.609 | 0.590 | 0.131 | 0.491 | 0.700 | 0.333 | 0.783 |
| p44 | Cravings | 40 | 0.308 | 0.356 | 0.190 | 0.217 | 0.437 | 0.000 | 0.714 |
| p59 | Cravings | 40 | 0.429 | 0.340 | 0.220 | 0.163 | 0.493 | 0.000 | 0.654 |
| p6 | Cravings | 40 | 0.464 | 0.461 | 0.084 | 0.393 | 0.516 | 0.308 | 0.621 |
| p61 | Cravings | 40 | 0.000 | 0.121 | 0.195 | 0.000 | 0.250 | 0.000 | 0.667 |
| p62 | Cravings | 40 | 0.069 | 0.106 | 0.126 | 0.000 | 0.203 | 0.000 | 0.462 |
| p63 | Cravings | 40 | 0.394 | 0.319 | 0.235 | 0.105 | 0.505 | 0.000 | 0.706 |
| p67 | Cravings | 32 | 0.000 | 0.000 | 0.000 | 0.000 | 0.000 | 0.000 | 0.000 |
| p7 | Cravings | 40 | 0.071 | 0.199 | 0.230 | 0.000 | 0.400 | 0.000 | 0.769 |
| p70 | Cravings | 6 | 0.000 | 0.000 | 0.000 | 0.000 | 0.000 | 0.000 | 0.000 |
| p74 | Cravings | 40 | 0.587 | 0.536 | 0.281 | 0.284 | 0.760 | 0.000 | 0.923 |
| p78 | Cravings | 40 | 0.927 | 0.879 | 0.130 | 0.884 | 0.945 | 0.392 | 0.968 |
| p79 | Cravings | 40 | 0.000 | 0.079 | 0.137 | 0.000 | 0.113 | 0.000 | 0.400 |
| p87 | Cravings | 40 | 0.986 | 0.980 | 0.025 | 0.981 | 0.989 | 0.847 | 0.997 |
| p9 | Cravings | 40 | 0.308 | 0.313 | 0.127 | 0.236 | 0.405 | 0.000 | 0.500 |
| p93 | Cravings | 40 | 0.879 | 0.844 | 0.075 | 0.794 | 0.894 | 0.681 | 0.946 |
| p95 | Cravings | 40 | 0.515 | 0.500 | 0.099 | 0.444 | 0.566 | 0.211 | 0.667 |
| p97 | Cravings | 34 | 0.000 | 0.000 | 0.000 | 0.000 | 0.000 | 0.000 | 0.000 |

F1-Score by participant × outcome (proportion below acceptable threshold of 0.5) (outcome: lapses)

| **Participant** | **Outcome** | **n** | **N below threshold** | **Proportion below threshold** |
| --- | --- | --- | --- | --- |
| p1 | Lapses | 40 | 27 | 0.675 |
| p101 | Lapses | 40 | 36 | 0.900 |
| p107 | Lapses | 40 | 34 | 0.850 |
| p12 | Lapses | 40 | 22 | 0.550 |
| p128 | Lapses | 40 | 20 | 0.500 |
| p131 | Lapses | 2 | 1 | 0.500 |
| p132 | Lapses | 20 | 20 | 1.000 |
| p135 | Lapses | 40 | 13 | 0.325 |
| p141 | Lapses | 40 | 35 | 0.875 |
| p144 | Lapses | 40 | 4 | 0.100 |
| p15 | Lapses | 40 | 10 | 0.250 |
| p21 | Lapses | 40 | 14 | 0.350 |
| p22 | Lapses | 40 | 17 | 0.425 |
| p30 | Lapses | 10 | 10 | 1.000 |
| p6 | Lapses | 40 | 0 | 0.000 |
| p61 | Lapses | 40 | 1 | 0.025 |
| p62 | Lapses | 20 | 20 | 1.000 |
| p63 | Lapses | 40 | 8 | 0.200 |
| p67 | Lapses | 10 | 10 | 1.000 |
| p7 | Lapses | 40 | 39 | 0.975 |
| p74 | Lapses | 10 | 10 | 1.000 |
| p87 | Lapses | 18 | 18 | 1.000 |
| p9 | Lapses | 10 | 10 | 1.000 |
| p95 | Lapses | 40 | 23 | 0.575 |
| p97 | Lapses | 40 | 39 | 0.975 |

F1-Score by participant × outcome (proportion below acceptable threshold of 0.5) (outcome: cravings)

| **Participant** | **Outcome** | **n** | **N below threshold** | **Proportion below threshold** |
| --- | --- | --- | --- | --- |
| p1 | Cravings | 40 | 40 | 1.000 |
| p10 | Cravings | 40 | 40 | 1.000 |
| p101 | Cravings | 40 | 2 | 0.050 |
| p107 | Cravings | 40 | 29 | 0.725 |
| p12 | Cravings | 40 | 40 | 1.000 |
| p128 | Cravings | 40 | 29 | 0.725 |
| p129 | Cravings | 40 | 39 | 0.975 |
| p131 | Cravings | 40 | 0 | 0.000 |
| p132 | Cravings | 40 | 0 | 0.000 |
| p135 | Cravings | 40 | 28 | 0.700 |
| p141 | Cravings | 40 | 2 | 0.050 |
| p144 | Cravings | 40 | 23 | 0.575 |
| p147 | Cravings | 40 | 24 | 0.600 |
| p15 | Cravings | 40 | 40 | 1.000 |
| p20 | Cravings | 40 | 0 | 0.000 |
| p21 | Cravings | 40 | 22 | 0.550 |
| p22 | Cravings | 40 | 12 | 0.300 |
| p27 | Cravings | 32 | 32 | 1.000 |
| p30 | Cravings | 40 | 39 | 0.975 |
| p39 | Cravings | 40 | 10 | 0.250 |
| p44 | Cravings | 40 | 31 | 0.775 |
| p59 | Cravings | 40 | 30 | 0.750 |
| p6 | Cravings | 40 | 24 | 0.600 |
| p61 | Cravings | 40 | 36 | 0.900 |
| p62 | Cravings | 40 | 40 | 1.000 |
| p63 | Cravings | 40 | 27 | 0.675 |
| p67 | Cravings | 32 | 32 | 1.000 |
| p7 | Cravings | 40 | 34 | 0.850 |
| p70 | Cravings | 6 | 6 | 1.000 |
| p74 | Cravings | 40 | 15 | 0.375 |
| p78 | Cravings | 40 | 1 | 0.025 |
| p79 | Cravings | 40 | 40 | 1.000 |
| p87 | Cravings | 40 | 0 | 0.000 |
| p9 | Cravings | 40 | 38 | 0.950 |
| p93 | Cravings | 40 | 0 | 0.000 |
| p95 | Cravings | 40 | 19 | 0.475 |
| p97 | Cravings | 34 | 34 | 1.000 |
