## Supplementary material for "Optimising supervised machine learning algorithms predicting cigarette cravings and lapses for a smoking cessation just-in-time adaptive intervention (JITAI)": S5 Appendix ROC-AUC performance summaries

### 1. Descriptive performance summaries: ROC-AUC

#### 1.1. By specific predictors

##### 1.1.1. By prompts per day

ROC-AUC by prompts per day

| **Prompts per day** | **n** | **Median** | **Mean** | **Standard deviation** | **25th percentile** | **75th percentile** | **Minimum** | **Maximum** |
| --- | --- | --- | --- | --- | --- | --- | --- | --- |
| 16 | 450 | 0.693 | 0.680 | 0.183 | 0.556 | 0.822 | 0.052 | 0.998 |
| 6 | 436 | 0.649 | 0.640 | 0.167 | 0.536 | 0.749 | 0.000 | 0.983 |
| 5 | 430 | 0.614 | 0.615 | 0.163 | 0.515 | 0.717 | 0.068 | 0.971 |
| 4 | 446 | 0.619 | 0.606 | 0.180 | 0.500 | 0.710 | 0.025 | 1.000 |
| 3 | 442 | 0.600 | 0.584 | 0.207 | 0.454 | 0.739 | 0.000 | 1.000 |

##### 1.1.2. By prompts per day - proportion below threshold

ROC-AUC by prompts per day (proportion below acceptable threshold of 0.5)

| **Prompts per day** | **n** | **N below threshold** | **Proportion below threshold** |
| --- | --- | --- | --- |
| 16 | 450 | 62 | 0.138 |
| 6 | 436 | 85 | 0.195 |
| 5 | 430 | 93 | 0.216 |
| 4 | 446 | 111 | 0.249 |
| 3 | 442 | 143 | 0.324 |

ROC-AUC by prompts per day × outcome (outcome: lapses)

| **Prompts per day** | **Outcome** | **n** | **Median** | **Mean** | **Standard deviation** | **25th percentile** | **75th percentile** | **Minimum** | **Maximum** |
| --- | --- | --- | --- | --- | --- | --- | --- | --- | --- |
| 16 | Lapses | 158 | 0.661 | 0.680 | 0.214 | 0.520 | 0.876 | 0.052 | 0.998 |
| 6 | Lapses | 156 | 0.628 | 0.637 | 0.197 | 0.498 | 0.791 | 0.017 | 0.968 |
| 5 | Lapses | 156 | 0.660 | 0.646 | 0.180 | 0.510 | 0.795 | 0.111 | 0.969 |
| 4 | Lapses | 156 | 0.613 | 0.631 | 0.206 | 0.494 | 0.786 | 0.028 | 0.975 |
| 3 | Lapses | 154 | 0.704 | 0.665 | 0.192 | 0.567 | 0.809 | 0.000 | 0.967 |

ROC-AUC by prompts per day × outcome (outcome: cravings)

| **Prompts per day** | **Outcome** | **n** | **Median** | **Mean** | **Standard deviation** | **25th percentile** | **75th percentile** | **Minimum** | **Maximum** |
| --- | --- | --- | --- | --- | --- | --- | --- | --- | --- |
| 16 | Cravings | 292 | 0.700 | 0.681 | 0.165 | 0.582 | 0.811 | 0.122 | 0.986 |
| 6 | Cravings | 280 | 0.652 | 0.641 | 0.148 | 0.575 | 0.736 | 0.000 | 0.983 |
| 5 | Cravings | 274 | 0.606 | 0.598 | 0.151 | 0.517 | 0.696 | 0.068 | 0.971 |
| 4 | Cravings | 290 | 0.622 | 0.593 | 0.164 | 0.500 | 0.695 | 0.025 | 1.000 |
| 3 | Cravings | 288 | 0.544 | 0.541 | 0.201 | 0.421 | 0.676 | 0.000 | 1.000 |

ROC-AUC by prompts per day × outcome (proportion below acceptable threshold of 0.5) (outcome: lapses)

| **Prompts per day** | **Outcome** | **n** | **N below threshold** | **Proportion below threshold** |
| --- | --- | --- | --- | --- |
| 16 | Lapses | 158 | 26 | 0.165 |
| 6 | Lapses | 156 | 40 | 0.256 |
| 5 | Lapses | 156 | 32 | 0.205 |
| 4 | Lapses | 156 | 40 | 0.256 |
| 3 | Lapses | 154 | 27 | 0.175 |

ROC-AUC by prompts per day × outcome (proportion below acceptable threshold of 0.5) (outcome: cravings)

| **Prompts per day** | **Outcome** | **n** | **N below threshold** | **Proportion below threshold** |
| --- | --- | --- | --- | --- |
| 16 | Cravings | 292 | 36 | 0.123 |
| 6 | Cravings | 280 | 45 | 0.161 |
| 5 | Cravings | 274 | 61 | 0.223 |
| 4 | Cravings | 290 | 71 | 0.245 |
| 3 | Cravings | 288 | 116 | 0.403 |

##### 1.1.3. By feature selection

ROC-AUC by feature selection

| **Feature selection** | **n** | **Median** | **Mean** | **Standard deviation** | **25th percentile** | **75th percentile** | **Minimum** | **Maximum** |
| --- | --- | --- | --- | --- | --- | --- | --- | --- |
| All features | 1,102 | 0.641 | 0.635 | 0.180 | 0.518 | 0.762 | 0.000 | 1.000 |
| Selected features | 1,102 | 0.628 | 0.615 | 0.187 | 0.504 | 0.740 | 0.000 | 0.994 |

##### 1.1.4. By feature selection - proportion below threshold

ROC-AUC by feature selection (proportion below acceptable threshold of 0.5)

| **Feature selection** | **n** | **N below threshold** | **Proportion below threshold** |
| --- | --- | --- | --- |
| All features | 1,102 | 229 | 0.208 |
| Selected features | 1,102 | 265 | 0.240 |

ROC-AUC by feature selection × outcome (outcome: lapses)

| **Feature selection** | **Outcome** | **n** | **Median** | **Mean** | **Standard deviation** | **25th percentile** | **75th percentile** | **Minimum** | **Maximum** |
| --- | --- | --- | --- | --- | --- | --- | --- | --- | --- |
| All features | Lapses | 390 | 0.660 | 0.656 | 0.197 | 0.518 | 0.811 | 0.000 | 0.998 |
| Selected features | Lapses | 390 | 0.657 | 0.648 | 0.200 | 0.508 | 0.808 | 0.000 | 0.994 |

ROC-AUC by feature selection × outcome (outcome: cravings)

| **Feature selection** | **Outcome** | **n** | **Median** | **Mean** | **Standard deviation** | **25th percentile** | **75th percentile** | **Minimum** | **Maximum** |
| --- | --- | --- | --- | --- | --- | --- | --- | --- | --- |
| All features | Cravings | 712 | 0.632 | 0.624 | 0.170 | 0.516 | 0.744 | 0.032 | 1.000 |
| Selected features | Cravings | 712 | 0.622 | 0.598 | 0.177 | 0.502 | 0.717 | 0.000 | 0.986 |

ROC-AUC by feature selection × outcome (proportion below acceptable threshold of 0.5) (outcome: lapses)

| **Feature selection** | **Outcome** | **n** | **N below threshold** | **Proportion below threshold** |
| --- | --- | --- | --- | --- |
| All features | Lapses | 390 | 75 | 0.192 |
| Selected features | Lapses | 390 | 90 | 0.231 |

ROC-AUC by feature selection × outcome (proportion below acceptable threshold of 0.5) (outcome: cravings)

| **Feature selection** | **Outcome** | **n** | **N below threshold** | **Proportion below threshold** |
| --- | --- | --- | --- | --- |
| All features | Cravings | 712 | 154 | 0.216 |
| Selected features | Cravings | 712 | 175 | 0.246 |

##### 1.1.5. By share of own data in training set

ROC-AUC by share of own data in training set

| **Share of own data in training set** | **n** | **Median** | **Mean** | **Standard deviation** | **25th percentile** | **75th percentile** | **Minimum** | **Maximum** |
| --- | --- | --- | --- | --- | --- | --- | --- | --- |
| None | 602 | 0.652 | 0.636 | 0.177 | 0.531 | 0.755 | 0.017 | 0.994 |
| 10% | 554 | 0.618 | 0.614 | 0.189 | 0.509 | 0.739 | 0.000 | 1.000 |
| 20% | 524 | 0.640 | 0.628 | 0.181 | 0.514 | 0.759 | 0.032 | 1.000 |
| 30% | 524 | 0.622 | 0.623 | 0.189 | 0.500 | 0.754 | 0.000 | 1.000 |

##### 1.1.6. By share of own data in training set - proportion below threshold

ROC-AUC by share of own data in training set (proportion below acceptable threshold of 0.5)

| **Share of own data in training set** | **n** | **N below threshold** | **Proportion below threshold** |
| --- | --- | --- | --- |
| None | 602 | 117 | 0.194 |
| 10% | 554 | 131 | 0.236 |
| 20% | 524 | 119 | 0.227 |
| 30% | 524 | 127 | 0.242 |

ROC-AUC by share of own data in training set × outcome (outcome: lapses)

| **Share of own data in training set** | **Outcome** | **n** | **Median** | **Mean** | **Standard deviation** | **25th percentile** | **75th percentile** | **Minimum** | **Maximum** |
| --- | --- | --- | --- | --- | --- | --- | --- | --- | --- |
| None | Lapses | 242 | 0.655 | 0.644 | 0.199 | 0.512 | 0.786 | 0.017 | 0.994 |
| 10% | Lapses | 198 | 0.607 | 0.627 | 0.207 | 0.510 | 0.765 | 0.000 | 0.994 |
| 20% | Lapses | 170 | 0.673 | 0.674 | 0.181 | 0.532 | 0.819 | 0.258 | 0.998 |
| 30% | Lapses | 170 | 0.694 | 0.670 | 0.201 | 0.513 | 0.852 | 0.206 | 0.997 |

ROC-AUC by share of own data in training set × outcome (outcome: cravings)

| **Share of own data in training set** | **Outcome** | **n** | **Median** | **Mean** | **Standard deviation** | **25th percentile** | **75th percentile** | **Minimum** | **Maximum** |
| --- | --- | --- | --- | --- | --- | --- | --- | --- | --- |
| None | Cravings | 360 | 0.650 | 0.630 | 0.160 | 0.544 | 0.734 | 0.025 | 0.983 |
| 10% | Cravings | 356 | 0.619 | 0.607 | 0.178 | 0.504 | 0.722 | 0.000 | 1.000 |
| 20% | Cravings | 354 | 0.632 | 0.606 | 0.177 | 0.503 | 0.728 | 0.032 | 1.000 |
| 30% | Cravings | 354 | 0.614 | 0.600 | 0.178 | 0.493 | 0.730 | 0.000 | 1.000 |

ROC-AUC by share of own data in training set × outcome (proportion below acceptable threshold of 0.5) (outcome: lapses)

| **Share of own data in training set** | **Outcome** | **n** | **N below threshold** | **Proportion below threshold** |
| --- | --- | --- | --- | --- |
| None | Lapses | 242 | 49 | 0.202 |
| 10% | Lapses | 198 | 44 | 0.222 |
| 20% | Lapses | 170 | 35 | 0.206 |
| 30% | Lapses | 170 | 37 | 0.218 |

ROC-AUC by share of own data in training set × outcome (proportion below acceptable threshold of 0.5) (outcome: cravings)

| **Share of own data in training set** | **Outcome** | **n** | **N below threshold** | **Proportion below threshold** |
| --- | --- | --- | --- | --- |
| None | Cravings | 360 | 68 | 0.189 |
| 10% | Cravings | 356 | 87 | 0.244 |
| 20% | Cravings | 354 | 84 | 0.237 |
| 30% | Cravings | 354 | 90 | 0.254 |

##### 1.1.7. By outcome

ROC-AUC by outcome

| **Outcome** | **n** | **Median** | **Mean** | **Standard deviation** | **25th percentile** | **75th percentile** | **Minimum** | **Maximum** |
| --- | --- | --- | --- | --- | --- | --- | --- | --- |
| Lapses | 780 | 0.659 | 0.652 | 0.198 | 0.514 | 0.809 | 0.000 | 0.998 |
| Cravings | 1,424 | 0.628 | 0.611 | 0.174 | 0.510 | 0.729 | 0.000 | 1.000 |

##### 1.1.8. By outcome - proportion below threshold

ROC-AUC by outcome (proportion below acceptable threshold of 0.5)

| **Outcome** | **n** | **N below threshold** | **Proportion below threshold** |
| --- | --- | --- | --- |
| Lapses | 780 | 165 | 0.212 |
| Cravings | 1,424 | 329 | 0.231 |

#### 1.2. By specification

##### 1.2.1. By specification

ROC-AUC by specification (prompts per day × use of feature selection × share of own data in training set × outcome)

| **Prompts per day** | **Feature selection** | **Share of own data in training set** | **Outcome** | **n** | **Median** | **Mean** | **Standard deviation** | **25th percentile** | **75th percentile** | **Minimum** | **Maximum** |
| --- | --- | --- | --- | --- | --- | --- | --- | --- | --- | --- | --- |
| 16 | All features | None | Lapses | 25 | 0.652 | 0.661 | 0.222 | 0.523 | 0.811 | 0.125 | 0.975 |
| 16 | All features | None | Cravings | 37 | 0.724 | 0.711 | 0.141 | 0.587 | 0.824 | 0.369 | 0.929 |
| 16 | All features | 10% | Lapses | 20 | 0.623 | 0.655 | 0.236 | 0.507 | 0.818 | 0.056 | 0.994 |
| 16 | All features | 10% | Cravings | 37 | 0.699 | 0.686 | 0.143 | 0.588 | 0.809 | 0.296 | 0.905 |
| 16 | All features | 20% | Lapses | 17 | 0.655 | 0.706 | 0.191 | 0.555 | 0.873 | 0.398 | 0.998 |
| 16 | All features | 20% | Cravings | 36 | 0.690 | 0.672 | 0.172 | 0.578 | 0.776 | 0.139 | 0.969 |
| 16 | All features | 30% | Lapses | 17 | 0.652 | 0.686 | 0.212 | 0.536 | 0.881 | 0.372 | 0.997 |
| 16 | All features | 30% | Cravings | 36 | 0.675 | 0.671 | 0.167 | 0.574 | 0.783 | 0.150 | 0.962 |
| 16 | Selected features | None | Lapses | 25 | 0.754 | 0.706 | 0.215 | 0.561 | 0.878 | 0.181 | 0.994 |
| 16 | Selected features | None | Cravings | 37 | 0.728 | 0.703 | 0.164 | 0.626 | 0.827 | 0.209 | 0.927 |
| 16 | Selected features | 10% | Lapses | 20 | 0.612 | 0.651 | 0.247 | 0.507 | 0.864 | 0.052 | 0.975 |
| 16 | Selected features | 10% | Cravings | 37 | 0.709 | 0.686 | 0.164 | 0.587 | 0.801 | 0.262 | 0.986 |
| 16 | Selected features | 20% | Lapses | 17 | 0.697 | 0.697 | 0.196 | 0.526 | 0.877 | 0.408 | 0.968 |
| 16 | Selected features | 20% | Cravings | 36 | 0.693 | 0.672 | 0.171 | 0.594 | 0.777 | 0.175 | 0.934 |
| 16 | Selected features | 30% | Lapses | 17 | 0.732 | 0.683 | 0.207 | 0.505 | 0.897 | 0.393 | 0.968 |
| 16 | Selected features | 30% | Cravings | 36 | 0.661 | 0.643 | 0.198 | 0.542 | 0.785 | 0.122 | 0.943 |
| 6 | All features | None | Lapses | 24 | 0.567 | 0.625 | 0.193 | 0.520 | 0.747 | 0.133 | 0.949 |
| 6 | All features | None | Cravings | 35 | 0.678 | 0.667 | 0.122 | 0.606 | 0.706 | 0.397 | 0.983 |
| 6 | All features | 10% | Lapses | 20 | 0.569 | 0.622 | 0.197 | 0.501 | 0.765 | 0.204 | 0.940 |
| 6 | All features | 10% | Cravings | 35 | 0.661 | 0.659 | 0.152 | 0.591 | 0.750 | 0.132 | 0.915 |
| 6 | All features | 20% | Lapses | 17 | 0.673 | 0.665 | 0.206 | 0.467 | 0.873 | 0.365 | 0.957 |
| 6 | All features | 20% | Cravings | 35 | 0.650 | 0.651 | 0.174 | 0.558 | 0.768 | 0.085 | 0.979 |
| 6 | All features | 30% | Lapses | 17 | 0.661 | 0.663 | 0.196 | 0.513 | 0.866 | 0.379 | 0.968 |
| 6 | All features | 30% | Cravings | 35 | 0.648 | 0.648 | 0.157 | 0.539 | 0.780 | 0.317 | 0.939 |
| 6 | Selected features | None | Lapses | 24 | 0.596 | 0.620 | 0.209 | 0.506 | 0.790 | 0.017 | 0.914 |
| 6 | Selected features | None | Cravings | 35 | 0.672 | 0.648 | 0.133 | 0.591 | 0.726 | 0.169 | 0.870 |
| 6 | Selected features | 10% | Lapses | 20 | 0.604 | 0.604 | 0.209 | 0.489 | 0.729 | 0.056 | 0.925 |
| 6 | Selected features | 10% | Cravings | 35 | 0.639 | 0.614 | 0.165 | 0.575 | 0.724 | 0.000 | 0.858 |
| 6 | Selected features | 20% | Lapses | 17 | 0.666 | 0.666 | 0.184 | 0.555 | 0.819 | 0.314 | 0.918 |
| 6 | Selected features | 20% | Cravings | 35 | 0.628 | 0.624 | 0.144 | 0.572 | 0.710 | 0.085 | 0.894 |
| 6 | Selected features | 30% | Lapses | 17 | 0.718 | 0.655 | 0.204 | 0.495 | 0.829 | 0.239 | 0.927 |
| 6 | Selected features | 30% | Cravings | 35 | 0.618 | 0.620 | 0.137 | 0.548 | 0.722 | 0.244 | 0.915 |
| 5 | All features | None | Lapses | 24 | 0.626 | 0.628 | 0.154 | 0.503 | 0.718 | 0.260 | 0.929 |
| 5 | All features | None | Cravings | 35 | 0.640 | 0.637 | 0.118 | 0.549 | 0.708 | 0.420 | 0.902 |
| 5 | All features | 10% | Lapses | 20 | 0.575 | 0.621 | 0.167 | 0.514 | 0.737 | 0.359 | 0.923 |
| 5 | All features | 10% | Cravings | 34 | 0.585 | 0.592 | 0.155 | 0.463 | 0.657 | 0.280 | 0.932 |
| 5 | All features | 20% | Lapses | 17 | 0.731 | 0.694 | 0.183 | 0.562 | 0.846 | 0.361 | 0.950 |
| 5 | All features | 20% | Cravings | 34 | 0.646 | 0.624 | 0.149 | 0.531 | 0.734 | 0.288 | 0.897 |
| 5 | All features | 30% | Lapses | 17 | 0.709 | 0.699 | 0.204 | 0.556 | 0.865 | 0.362 | 0.969 |
| 5 | All features | 30% | Cravings | 34 | 0.582 | 0.578 | 0.176 | 0.449 | 0.694 | 0.182 | 0.971 |
| 5 | Selected features | None | Lapses | 24 | 0.622 | 0.619 | 0.191 | 0.503 | 0.781 | 0.200 | 0.919 |
| 5 | Selected features | None | Cravings | 35 | 0.633 | 0.593 | 0.139 | 0.531 | 0.680 | 0.160 | 0.812 |
| 5 | Selected features | 10% | Lapses | 20 | 0.603 | 0.610 | 0.193 | 0.490 | 0.726 | 0.111 | 0.937 |
| 5 | Selected features | 10% | Cravings | 34 | 0.616 | 0.587 | 0.177 | 0.518 | 0.697 | 0.068 | 0.886 |
| 5 | Selected features | 20% | Lapses | 17 | 0.667 | 0.652 | 0.165 | 0.537 | 0.770 | 0.388 | 0.950 |
| 5 | Selected features | 20% | Cravings | 34 | 0.603 | 0.589 | 0.149 | 0.541 | 0.675 | 0.179 | 0.846 |
| 5 | Selected features | 30% | Lapses | 17 | 0.724 | 0.672 | 0.190 | 0.483 | 0.833 | 0.391 | 0.968 |
| 5 | Selected features | 30% | Cravings | 34 | 0.595 | 0.583 | 0.143 | 0.504 | 0.685 | 0.235 | 0.848 |
| 4 | All features | None | Lapses | 24 | 0.618 | 0.607 | 0.207 | 0.443 | 0.727 | 0.175 | 0.953 |
| 4 | All features | None | Cravings | 37 | 0.645 | 0.638 | 0.163 | 0.576 | 0.760 | 0.125 | 0.974 |
| 4 | All features | 10% | Lapses | 20 | 0.605 | 0.621 | 0.216 | 0.552 | 0.720 | 0.028 | 0.963 |
| 4 | All features | 10% | Cravings | 36 | 0.617 | 0.594 | 0.167 | 0.499 | 0.696 | 0.086 | 1.000 |
| 4 | All features | 20% | Lapses | 17 | 0.613 | 0.641 | 0.221 | 0.485 | 0.789 | 0.258 | 0.957 |
| 4 | All features | 20% | Cravings | 36 | 0.627 | 0.582 | 0.178 | 0.483 | 0.681 | 0.032 | 1.000 |
| 4 | All features | 30% | Lapses | 17 | 0.667 | 0.667 | 0.205 | 0.519 | 0.821 | 0.276 | 0.967 |
| 4 | All features | 30% | Cravings | 36 | 0.651 | 0.620 | 0.164 | 0.518 | 0.695 | 0.185 | 1.000 |
| 4 | Selected features | None | Lapses | 24 | 0.660 | 0.665 | 0.207 | 0.520 | 0.821 | 0.225 | 0.975 |
| 4 | Selected features | None | Cravings | 37 | 0.633 | 0.600 | 0.178 | 0.544 | 0.697 | 0.025 | 0.897 |
| 4 | Selected features | 10% | Lapses | 20 | 0.594 | 0.601 | 0.204 | 0.508 | 0.688 | 0.028 | 0.933 |
| 4 | Selected features | 10% | Cravings | 36 | 0.552 | 0.556 | 0.145 | 0.507 | 0.649 | 0.057 | 0.837 |
| 4 | Selected features | 20% | Lapses | 17 | 0.596 | 0.636 | 0.178 | 0.562 | 0.710 | 0.274 | 0.921 |
| 4 | Selected features | 20% | Cravings | 36 | 0.641 | 0.585 | 0.161 | 0.503 | 0.694 | 0.065 | 0.789 |
| 4 | Selected features | 30% | Lapses | 17 | 0.608 | 0.615 | 0.229 | 0.447 | 0.821 | 0.259 | 0.904 |
| 4 | Selected features | 30% | Cravings | 36 | 0.571 | 0.563 | 0.156 | 0.472 | 0.687 | 0.222 | 0.818 |
| 3 | All features | None | Lapses | 24 | 0.713 | 0.658 | 0.189 | 0.553 | 0.767 | 0.207 | 0.967 |
| 3 | All features | None | Cravings | 36 | 0.595 | 0.557 | 0.171 | 0.446 | 0.669 | 0.198 | 0.904 |
| 3 | All features | 10% | Lapses | 19 | 0.697 | 0.669 | 0.207 | 0.594 | 0.793 | 0.000 | 0.960 |
| 3 | All features | 10% | Cravings | 36 | 0.568 | 0.576 | 0.217 | 0.466 | 0.664 | 0.040 | 1.000 |
| 3 | All features | 20% | Lapses | 17 | 0.731 | 0.708 | 0.149 | 0.630 | 0.788 | 0.420 | 0.939 |
| 3 | All features | 20% | Cravings | 36 | 0.535 | 0.558 | 0.210 | 0.424 | 0.708 | 0.043 | 0.891 |
| 3 | All features | 30% | Lapses | 17 | 0.715 | 0.690 | 0.207 | 0.667 | 0.850 | 0.206 | 0.895 |
| 3 | All features | 30% | Cravings | 36 | 0.555 | 0.551 | 0.175 | 0.423 | 0.676 | 0.167 | 0.850 |
| 3 | Selected features | None | Lapses | 24 | 0.686 | 0.648 | 0.211 | 0.551 | 0.791 | 0.033 | 0.966 |
| 3 | Selected features | None | Cravings | 36 | 0.548 | 0.541 | 0.181 | 0.455 | 0.681 | 0.100 | 0.862 |
| 3 | Selected features | 10% | Lapses | 19 | 0.646 | 0.619 | 0.217 | 0.548 | 0.766 | 0.000 | 0.871 |
| 3 | Selected features | 10% | Cravings | 36 | 0.491 | 0.514 | 0.212 | 0.423 | 0.656 | 0.038 | 0.962 |
| 3 | Selected features | 20% | Lapses | 17 | 0.714 | 0.672 | 0.161 | 0.569 | 0.810 | 0.352 | 0.871 |
| 3 | Selected features | 20% | Cravings | 36 | 0.499 | 0.509 | 0.200 | 0.361 | 0.667 | 0.068 | 0.826 |
| 3 | Selected features | 30% | Lapses | 17 | 0.704 | 0.674 | 0.199 | 0.571 | 0.825 | 0.265 | 0.935 |
| 3 | Selected features | 30% | Cravings | 36 | 0.549 | 0.518 | 0.243 | 0.362 | 0.701 | 0.000 | 0.920 |

##### 1.2.2. By specification - proportion below threshold

ROC-AUC by specification (prompts per day × use of feature selection × share of own data in training set × outcome) - proportion below acceptable threshold of 0.5

| **Prompts per day** | **Feature selection** | **Share of own data in training set** | **Outcome** | **n** | **N below threshold** | **Proportion below threshold** |
| --- | --- | --- | --- | --- | --- | --- |
| 16 | All features | None | Lapses | 25 | 3 | 0.120 |
| 16 | All features | None | Cravings | 37 | 2 | 0.054 |
| 16 | All features | 10% | Lapses | 20 | 4 | 0.200 |
| 16 | All features | 10% | Cravings | 37 | 3 | 0.081 |
| 16 | All features | 20% | Lapses | 17 | 2 | 0.118 |
| 16 | All features | 20% | Cravings | 36 | 5 | 0.139 |
| 16 | All features | 30% | Lapses | 17 | 3 | 0.176 |
| 16 | All features | 30% | Cravings | 36 | 5 | 0.139 |
| 16 | Selected features | None | Lapses | 25 | 3 | 0.120 |
| 16 | Selected features | None | Cravings | 37 | 4 | 0.108 |
| 16 | Selected features | 10% | Lapses | 20 | 5 | 0.250 |
| 16 | Selected features | 10% | Cravings | 37 | 5 | 0.135 |
| 16 | Selected features | 20% | Lapses | 17 | 2 | 0.118 |
| 16 | Selected features | 20% | Cravings | 36 | 5 | 0.139 |
| 16 | Selected features | 30% | Lapses | 17 | 4 | 0.235 |
| 16 | Selected features | 30% | Cravings | 36 | 7 | 0.194 |
| 6 | All features | None | Lapses | 24 | 5 | 0.208 |
| 6 | All features | None | Cravings | 35 | 5 | 0.143 |
| 6 | All features | 10% | Lapses | 20 | 5 | 0.250 |
| 6 | All features | 10% | Cravings | 35 | 6 | 0.171 |
| 6 | All features | 20% | Lapses | 17 | 6 | 0.353 |
| 6 | All features | 20% | Cravings | 35 | 7 | 0.200 |
| 6 | All features | 30% | Lapses | 17 | 3 | 0.176 |
| 6 | All features | 30% | Cravings | 35 | 6 | 0.171 |
| 6 | Selected features | None | Lapses | 24 | 6 | 0.250 |
| 6 | Selected features | None | Cravings | 35 | 5 | 0.143 |
| 6 | Selected features | 10% | Lapses | 20 | 6 | 0.300 |
| 6 | Selected features | 10% | Cravings | 35 | 5 | 0.143 |
| 6 | Selected features | 20% | Lapses | 17 | 4 | 0.235 |
| 6 | Selected features | 20% | Cravings | 35 | 5 | 0.143 |
| 6 | Selected features | 30% | Lapses | 17 | 5 | 0.294 |
| 6 | Selected features | 30% | Cravings | 35 | 6 | 0.171 |
| 5 | All features | None | Lapses | 24 | 4 | 0.167 |
| 5 | All features | None | Cravings | 35 | 5 | 0.143 |
| 5 | All features | 10% | Lapses | 20 | 3 | 0.150 |
| 5 | All features | 10% | Cravings | 34 | 10 | 0.294 |
| 5 | All features | 20% | Lapses | 17 | 3 | 0.176 |
| 5 | All features | 20% | Cravings | 34 | 6 | 0.176 |
| 5 | All features | 30% | Lapses | 17 | 3 | 0.176 |
| 5 | All features | 30% | Cravings | 34 | 10 | 0.294 |
| 5 | Selected features | None | Lapses | 24 | 5 | 0.208 |
| 5 | Selected features | None | Cravings | 35 | 8 | 0.229 |
| 5 | Selected features | 10% | Lapses | 20 | 6 | 0.300 |
| 5 | Selected features | 10% | Cravings | 34 | 8 | 0.235 |
| 5 | Selected features | 20% | Lapses | 17 | 3 | 0.176 |
| 5 | Selected features | 20% | Cravings | 34 | 6 | 0.176 |
| 5 | Selected features | 30% | Lapses | 17 | 5 | 0.294 |
| 5 | Selected features | 30% | Cravings | 34 | 8 | 0.235 |
| 4 | All features | None | Lapses | 24 | 7 | 0.292 |
| 4 | All features | None | Cravings | 37 | 7 | 0.189 |
| 4 | All features | 10% | Lapses | 20 | 4 | 0.200 |
| 4 | All features | 10% | Cravings | 36 | 9 | 0.250 |
| 4 | All features | 20% | Lapses | 17 | 5 | 0.294 |
| 4 | All features | 20% | Cravings | 36 | 12 | 0.333 |
| 4 | All features | 30% | Lapses | 17 | 3 | 0.176 |
| 4 | All features | 30% | Cravings | 36 | 6 | 0.167 |
| 4 | Selected features | None | Lapses | 24 | 5 | 0.208 |
| 4 | Selected features | None | Cravings | 37 | 7 | 0.189 |
| 4 | Selected features | 10% | Lapses | 20 | 5 | 0.250 |
| 4 | Selected features | 10% | Cravings | 36 | 9 | 0.250 |
| 4 | Selected features | 20% | Lapses | 17 | 4 | 0.235 |
| 4 | Selected features | 20% | Cravings | 36 | 8 | 0.222 |
| 4 | Selected features | 30% | Lapses | 17 | 7 | 0.412 |
| 4 | Selected features | 30% | Cravings | 36 | 13 | 0.361 |
| 3 | All features | None | Lapses | 24 | 5 | 0.208 |
| 3 | All features | None | Cravings | 36 | 12 | 0.333 |
| 3 | All features | 10% | Lapses | 19 | 2 | 0.105 |
| 3 | All features | 10% | Cravings | 36 | 14 | 0.389 |
| 3 | All features | 20% | Lapses | 17 | 3 | 0.176 |
| 3 | All features | 20% | Cravings | 36 | 12 | 0.333 |
| 3 | All features | 30% | Lapses | 17 | 2 | 0.118 |
| 3 | All features | 30% | Cravings | 36 | 12 | 0.333 |
| 3 | Selected features | None | Lapses | 24 | 6 | 0.250 |
| 3 | Selected features | None | Cravings | 36 | 13 | 0.361 |
| 3 | Selected features | 10% | Lapses | 19 | 4 | 0.211 |
| 3 | Selected features | 10% | Cravings | 36 | 18 | 0.500 |
| 3 | Selected features | 20% | Lapses | 17 | 3 | 0.176 |
| 3 | Selected features | 20% | Cravings | 36 | 18 | 0.500 |
| 3 | Selected features | 30% | Lapses | 17 | 2 | 0.118 |
| 3 | Selected features | 30% | Cravings | 36 | 17 | 0.472 |

#### 1.3. By participant

##### 1.3.1. By participant (overall)

ROC-AUC by participant

| **Participant** | **n** | **Median** | **Mean** | **Standard deviation** | **25th percentile** | **75th percentile** | **Minimum** | **Maximum** |
| --- | --- | --- | --- | --- | --- | --- | --- | --- |
| p1 | 80 | 0.581 | 0.588 | 0.116 | 0.505 | 0.680 | 0.336 | 0.885 |
| p10 | 40 | 0.672 | 0.657 | 0.116 | 0.627 | 0.726 | 0.345 | 0.833 |
| p101 | 80 | 0.717 | 0.713 | 0.065 | 0.679 | 0.758 | 0.553 | 0.876 |
| p107 | 80 | 0.667 | 0.670 | 0.102 | 0.602 | 0.738 | 0.436 | 0.865 |
| p12 | 80 | 0.433 | 0.435 | 0.128 | 0.362 | 0.505 | 0.167 | 0.758 |
| p128 | 80 | 0.488 | 0.496 | 0.087 | 0.428 | 0.567 | 0.286 | 0.697 |
| p129 | 40 | 0.545 | 0.521 | 0.234 | 0.350 | 0.687 | 0.040 | 0.927 |
| p131 | 42 | 0.620 | 0.624 | 0.055 | 0.595 | 0.654 | 0.484 | 0.772 |
| p132 | 60 | 0.579 | 0.612 | 0.185 | 0.481 | 0.787 | 0.161 | 0.927 |
| p135 | 80 | 0.683 | 0.673 | 0.168 | 0.505 | 0.829 | 0.385 | 0.935 |
| p141 | 80 | 0.698 | 0.709 | 0.105 | 0.645 | 0.790 | 0.500 | 1.000 |
| p144 | 80 | 0.882 | 0.838 | 0.103 | 0.752 | 0.921 | 0.612 | 0.978 |
| p147 | 40 | 0.650 | 0.586 | 0.122 | 0.491 | 0.686 | 0.323 | 0.728 |
| p15 | 80 | 0.551 | 0.548 | 0.073 | 0.493 | 0.595 | 0.381 | 0.733 |
| p20 | 40 | 0.738 | 0.720 | 0.205 | 0.606 | 0.885 | 0.241 | 1.000 |
| p21 | 80 | 0.557 | 0.562 | 0.082 | 0.508 | 0.625 | 0.316 | 0.755 |
| p22 | 80 | 0.795 | 0.762 | 0.157 | 0.654 | 0.902 | 0.325 | 0.964 |
| p27 | 32 | 0.613 | 0.562 | 0.261 | 0.344 | 0.779 | 0.085 | 0.926 |
| p30 | 50 | 0.619 | 0.610 | 0.196 | 0.465 | 0.739 | 0.130 | 0.994 |
| p39 | 40 | 0.695 | 0.685 | 0.121 | 0.581 | 0.804 | 0.481 | 0.862 |
| p44 | 40 | 0.620 | 0.631 | 0.156 | 0.520 | 0.730 | 0.308 | 0.908 |
| p59 | 40 | 0.644 | 0.650 | 0.155 | 0.531 | 0.801 | 0.263 | 0.875 |
| p6 | 80 | 0.573 | 0.594 | 0.103 | 0.537 | 0.661 | 0.259 | 0.886 |
| p61 | 80 | 0.832 | 0.745 | 0.229 | 0.596 | 0.923 | 0.103 | 0.981 |
| p62 | 60 | 0.534 | 0.478 | 0.229 | 0.328 | 0.649 | 0.000 | 0.812 |
| p63 | 80 | 0.755 | 0.757 | 0.144 | 0.686 | 0.868 | 0.259 | 0.998 |
| p67 | 42 | 0.310 | 0.409 | 0.324 | 0.137 | 0.685 | 0.000 | 0.984 |
| p7 | 80 | 0.677 | 0.667 | 0.148 | 0.593 | 0.751 | 0.307 | 0.969 |
| p70 | 6 | 0.814 | 0.784 | 0.171 | 0.701 | 0.891 | 0.513 | 0.986 |
| p74 | 50 | 0.636 | 0.622 | 0.142 | 0.539 | 0.696 | 0.278 | 0.949 |
| p78 | 40 | 0.654 | 0.617 | 0.153 | 0.538 | 0.714 | 0.130 | 0.904 |
| p79 | 40 | 0.586 | 0.566 | 0.198 | 0.432 | 0.727 | 0.032 | 0.886 |
| p87 | 58 | 0.676 | 0.626 | 0.242 | 0.516 | 0.807 | 0.000 | 0.962 |
| p9 | 50 | 0.639 | 0.563 | 0.207 | 0.563 | 0.695 | 0.017 | 0.789 |
| p93 | 40 | 0.643 | 0.636 | 0.129 | 0.548 | 0.753 | 0.388 | 0.835 |
| p95 | 80 | 0.557 | 0.569 | 0.152 | 0.452 | 0.718 | 0.206 | 0.801 |
| p97 | 74 | 0.615 | 0.592 | 0.200 | 0.477 | 0.753 | 0.043 | 0.927 |

##### 1.3.2. By participant (overall) - proportion below threshold

ROC-AUC by participant (proportion below acceptable threshold of 0.5)

| **Participant** | **n** | **N below threshold** | **Proportion below threshold** |
| --- | --- | --- | --- |
| p1 | 80 | 17 | 0.212 |
| p10 | 40 | 4 | 0.100 |
| p101 | 80 | 0 | 0.000 |
| p107 | 80 | 4 | 0.050 |
| p12 | 80 | 58 | 0.725 |
| p128 | 80 | 43 | 0.537 |
| p129 | 40 | 17 | 0.425 |
| p131 | 42 | 1 | 0.024 |
| p132 | 60 | 16 | 0.267 |
| p135 | 80 | 17 | 0.212 |
| p141 | 80 | 0 | 0.000 |
| p144 | 80 | 0 | 0.000 |
| p147 | 40 | 11 | 0.275 |
| p15 | 80 | 24 | 0.300 |
| p20 | 40 | 4 | 0.100 |
| p21 | 80 | 17 | 0.212 |
| p22 | 80 | 6 | 0.075 |
| p27 | 32 | 12 | 0.375 |
| p30 | 50 | 16 | 0.320 |
| p39 | 40 | 3 | 0.075 |
| p44 | 40 | 7 | 0.175 |
| p59 | 40 | 7 | 0.175 |
| p6 | 80 | 12 | 0.150 |
| p61 | 80 | 16 | 0.200 |
| p62 | 60 | 24 | 0.400 |
| p63 | 80 | 3 | 0.037 |
| p67 | 42 | 26 | 0.619 |
| p7 | 80 | 8 | 0.100 |
| p70 | 6 | 0 | 0.000 |
| p74 | 50 | 11 | 0.220 |
| p78 | 40 | 10 | 0.250 |
| p79 | 40 | 14 | 0.350 |
| p87 | 58 | 14 | 0.241 |
| p9 | 50 | 10 | 0.200 |
| p93 | 40 | 8 | 0.200 |
| p95 | 80 | 30 | 0.375 |
| p97 | 74 | 24 | 0.324 |

ROC-AUC by participant × outcome (outcome: lapses)

| **Participant** | **Outcome** | **n** | **Median** | **Mean** | **Standard deviation** | **25th percentile** | **75th percentile** | **Minimum** | **Maximum** |
| --- | --- | --- | --- | --- | --- | --- | --- | --- | --- |
| p1 | Lapses | 40 | 0.531 | 0.541 | 0.074 | 0.501 | 0.570 | 0.391 | 0.715 |
| p101 | Lapses | 40 | 0.697 | 0.699 | 0.067 | 0.666 | 0.736 | 0.553 | 0.876 |
| p107 | Lapses | 40 | 0.667 | 0.676 | 0.098 | 0.610 | 0.738 | 0.436 | 0.865 |
| p12 | Lapses | 40 | 0.392 | 0.393 | 0.079 | 0.350 | 0.446 | 0.239 | 0.544 |
| p128 | Lapses | 40 | 0.455 | 0.476 | 0.090 | 0.410 | 0.506 | 0.362 | 0.697 |
| p131 | Lapses | 2 | 0.726 | 0.726 | 0.066 | 0.702 | 0.749 | 0.679 | 0.772 |
| p132 | Lapses | 20 | 0.506 | 0.506 | 0.118 | 0.444 | 0.567 | 0.250 | 0.800 |
| p135 | Lapses | 40 | 0.830 | 0.827 | 0.053 | 0.785 | 0.867 | 0.731 | 0.935 |
| p141 | Lapses | 40 | 0.691 | 0.677 | 0.075 | 0.644 | 0.714 | 0.512 | 0.829 |
| p144 | Lapses | 40 | 0.917 | 0.910 | 0.045 | 0.894 | 0.940 | 0.812 | 0.978 |
| p15 | Lapses | 40 | 0.567 | 0.553 | 0.066 | 0.495 | 0.597 | 0.434 | 0.674 |
| p21 | Lapses | 40 | 0.541 | 0.546 | 0.077 | 0.499 | 0.575 | 0.344 | 0.755 |
| p22 | Lapses | 40 | 0.904 | 0.884 | 0.062 | 0.840 | 0.937 | 0.762 | 0.964 |
| p30 | Lapses | 10 | 0.867 | 0.786 | 0.235 | 0.717 | 0.972 | 0.308 | 0.994 |
| p6 | Lapses | 40 | 0.596 | 0.618 | 0.122 | 0.556 | 0.711 | 0.259 | 0.886 |
| p61 | Lapses | 40 | 0.913 | 0.886 | 0.081 | 0.824 | 0.957 | 0.730 | 0.981 |
| p62 | Lapses | 20 | 0.205 | 0.259 | 0.221 | 0.055 | 0.457 | 0.000 | 0.693 |
| p63 | Lapses | 40 | 0.869 | 0.853 | 0.093 | 0.769 | 0.930 | 0.686 | 0.998 |
| p67 | Lapses | 10 | 0.892 | 0.797 | 0.209 | 0.601 | 0.962 | 0.483 | 0.984 |
| p7 | Lapses | 40 | 0.661 | 0.648 | 0.094 | 0.603 | 0.704 | 0.384 | 0.848 |
| p74 | Lapses | 10 | 0.787 | 0.787 | 0.109 | 0.718 | 0.856 | 0.603 | 0.949 |
| p87 | Lapses | 18 | 0.699 | 0.701 | 0.128 | 0.622 | 0.788 | 0.436 | 0.949 |
| p9 | Lapses | 10 | 0.178 | 0.190 | 0.137 | 0.127 | 0.236 | 0.017 | 0.500 |
| p95 | Lapses | 40 | 0.477 | 0.470 | 0.096 | 0.408 | 0.548 | 0.206 | 0.608 |
| p97 | Lapses | 40 | 0.707 | 0.682 | 0.154 | 0.531 | 0.798 | 0.333 | 0.927 |

ROC-AUC by participant × outcome (outcome: cravings)

| **Participant** | **Outcome** | **n** | **Median** | **Mean** | **Standard deviation** | **25th percentile** | **75th percentile** | **Minimum** | **Maximum** |
| --- | --- | --- | --- | --- | --- | --- | --- | --- | --- |
| p1 | Cravings | 40 | 0.666 | 0.635 | 0.132 | 0.590 | 0.701 | 0.336 | 0.885 |
| p10 | Cravings | 40 | 0.672 | 0.657 | 0.116 | 0.627 | 0.726 | 0.345 | 0.833 |
| p101 | Cravings | 40 | 0.728 | 0.727 | 0.060 | 0.706 | 0.771 | 0.554 | 0.839 |
| p107 | Cravings | 40 | 0.667 | 0.665 | 0.106 | 0.592 | 0.738 | 0.471 | 0.839 |
| p12 | Cravings | 40 | 0.472 | 0.476 | 0.154 | 0.420 | 0.596 | 0.167 | 0.758 |
| p128 | Cravings | 40 | 0.524 | 0.517 | 0.081 | 0.462 | 0.585 | 0.286 | 0.641 |
| p129 | Cravings | 40 | 0.545 | 0.521 | 0.234 | 0.350 | 0.687 | 0.040 | 0.927 |
| p131 | Cravings | 40 | 0.619 | 0.619 | 0.051 | 0.592 | 0.654 | 0.484 | 0.760 |
| p132 | Cravings | 40 | 0.671 | 0.665 | 0.190 | 0.538 | 0.825 | 0.161 | 0.927 |
| p135 | Cravings | 40 | 0.505 | 0.519 | 0.074 | 0.472 | 0.598 | 0.385 | 0.634 |
| p141 | Cravings | 40 | 0.766 | 0.741 | 0.121 | 0.650 | 0.834 | 0.500 | 1.000 |
| p144 | Cravings | 40 | 0.752 | 0.765 | 0.093 | 0.694 | 0.816 | 0.612 | 0.944 |
| p147 | Cravings | 40 | 0.650 | 0.586 | 0.122 | 0.491 | 0.686 | 0.323 | 0.728 |
| p15 | Cravings | 40 | 0.534 | 0.543 | 0.079 | 0.491 | 0.587 | 0.381 | 0.733 |
| p20 | Cravings | 40 | 0.738 | 0.720 | 0.205 | 0.606 | 0.885 | 0.241 | 1.000 |
| p21 | Cravings | 40 | 0.590 | 0.578 | 0.084 | 0.518 | 0.648 | 0.316 | 0.704 |
| p22 | Cravings | 40 | 0.652 | 0.640 | 0.125 | 0.551 | 0.738 | 0.325 | 0.817 |
| p27 | Cravings | 32 | 0.613 | 0.562 | 0.261 | 0.344 | 0.779 | 0.085 | 0.926 |
| p30 | Cravings | 40 | 0.571 | 0.566 | 0.160 | 0.451 | 0.675 | 0.130 | 0.850 |
| p39 | Cravings | 40 | 0.695 | 0.685 | 0.121 | 0.581 | 0.804 | 0.481 | 0.862 |
| p44 | Cravings | 40 | 0.620 | 0.631 | 0.156 | 0.520 | 0.730 | 0.308 | 0.908 |
| p59 | Cravings | 40 | 0.644 | 0.650 | 0.155 | 0.531 | 0.801 | 0.263 | 0.875 |
| p6 | Cravings | 40 | 0.556 | 0.570 | 0.073 | 0.530 | 0.626 | 0.451 | 0.746 |
| p61 | Cravings | 40 | 0.591 | 0.605 | 0.243 | 0.429 | 0.856 | 0.103 | 0.979 |
| p62 | Cravings | 40 | 0.599 | 0.587 | 0.137 | 0.520 | 0.682 | 0.235 | 0.812 |
| p63 | Cravings | 40 | 0.691 | 0.661 | 0.120 | 0.639 | 0.732 | 0.259 | 0.813 |
| p67 | Cravings | 32 | 0.182 | 0.288 | 0.250 | 0.098 | 0.406 | 0.000 | 0.932 |
| p7 | Cravings | 40 | 0.705 | 0.686 | 0.187 | 0.568 | 0.827 | 0.307 | 0.969 |
| p70 | Cravings | 6 | 0.814 | 0.784 | 0.171 | 0.701 | 0.891 | 0.513 | 0.986 |
| p74 | Cravings | 40 | 0.607 | 0.581 | 0.117 | 0.497 | 0.681 | 0.278 | 0.730 |
| p78 | Cravings | 40 | 0.654 | 0.617 | 0.153 | 0.538 | 0.714 | 0.130 | 0.904 |
| p79 | Cravings | 40 | 0.586 | 0.566 | 0.198 | 0.432 | 0.727 | 0.032 | 0.886 |
| p87 | Cravings | 40 | 0.652 | 0.592 | 0.273 | 0.413 | 0.825 | 0.000 | 0.962 |
| p9 | Cravings | 40 | 0.659 | 0.656 | 0.068 | 0.602 | 0.702 | 0.464 | 0.789 |
| p93 | Cravings | 40 | 0.643 | 0.636 | 0.129 | 0.548 | 0.753 | 0.388 | 0.835 |
| p95 | Cravings | 40 | 0.720 | 0.668 | 0.132 | 0.620 | 0.767 | 0.348 | 0.801 |
| p97 | Cravings | 34 | 0.486 | 0.487 | 0.198 | 0.383 | 0.622 | 0.043 | 0.902 |

ROC-AUC by participant × outcome (proportion below acceptable threshold of 0.5) (outcome: lapses)

| **Participant** | **Outcome** | **n** | **N below threshold** | **Proportion below threshold** |
| --- | --- | --- | --- | --- |
| p1 | Lapses | 40 | 9 | 0.225 |
| p101 | Lapses | 40 | 0 | 0.000 |
| p107 | Lapses | 40 | 1 | 0.025 |
| p12 | Lapses | 40 | 35 | 0.875 |
| p128 | Lapses | 40 | 28 | 0.700 |
| p131 | Lapses | 2 | 0 | 0.000 |
| p132 | Lapses | 20 | 9 | 0.450 |
| p135 | Lapses | 40 | 0 | 0.000 |
| p141 | Lapses | 40 | 0 | 0.000 |
| p144 | Lapses | 40 | 0 | 0.000 |
| p15 | Lapses | 40 | 11 | 0.275 |
| p21 | Lapses | 40 | 10 | 0.250 |
| p22 | Lapses | 40 | 0 | 0.000 |
| p30 | Lapses | 10 | 2 | 0.200 |
| p6 | Lapses | 40 | 3 | 0.075 |
| p61 | Lapses | 40 | 0 | 0.000 |
| p62 | Lapses | 20 | 16 | 0.800 |
| p63 | Lapses | 40 | 0 | 0.000 |
| p67 | Lapses | 10 | 1 | 0.100 |
| p7 | Lapses | 40 | 2 | 0.050 |
| p74 | Lapses | 10 | 0 | 0.000 |
| p87 | Lapses | 18 | 1 | 0.056 |
| p9 | Lapses | 10 | 9 | 0.900 |
| p95 | Lapses | 40 | 22 | 0.550 |
| p97 | Lapses | 40 | 6 | 0.150 |

ROC-AUC by participant × outcome (proportion below acceptable threshold of 0.5) (outcome: cravings)

| **Participant** | **Outcome** | **n** | **N below threshold** | **Proportion below threshold** |
| --- | --- | --- | --- | --- |
| p1 | Cravings | 40 | 8 | 0.200 |
| p10 | Cravings | 40 | 4 | 0.100 |
| p101 | Cravings | 40 | 0 | 0.000 |
| p107 | Cravings | 40 | 3 | 0.075 |
| p12 | Cravings | 40 | 23 | 0.575 |
| p128 | Cravings | 40 | 15 | 0.375 |
| p129 | Cravings | 40 | 17 | 0.425 |
| p131 | Cravings | 40 | 1 | 0.025 |
| p132 | Cravings | 40 | 7 | 0.175 |
| p135 | Cravings | 40 | 17 | 0.425 |
| p141 | Cravings | 40 | 0 | 0.000 |
| p144 | Cravings | 40 | 0 | 0.000 |
| p147 | Cravings | 40 | 11 | 0.275 |
| p15 | Cravings | 40 | 13 | 0.325 |
| p20 | Cravings | 40 | 4 | 0.100 |
| p21 | Cravings | 40 | 7 | 0.175 |
| p22 | Cravings | 40 | 6 | 0.150 |
| p27 | Cravings | 32 | 12 | 0.375 |
| p30 | Cravings | 40 | 14 | 0.350 |
| p39 | Cravings | 40 | 3 | 0.075 |
| p44 | Cravings | 40 | 7 | 0.175 |
| p59 | Cravings | 40 | 7 | 0.175 |
| p6 | Cravings | 40 | 9 | 0.225 |
| p61 | Cravings | 40 | 16 | 0.400 |
| p62 | Cravings | 40 | 8 | 0.200 |
| p63 | Cravings | 40 | 3 | 0.075 |
| p67 | Cravings | 32 | 25 | 0.781 |
| p7 | Cravings | 40 | 6 | 0.150 |
| p70 | Cravings | 6 | 0 | 0.000 |
| p74 | Cravings | 40 | 11 | 0.275 |
| p78 | Cravings | 40 | 10 | 0.250 |
| p79 | Cravings | 40 | 14 | 0.350 |
| p87 | Cravings | 40 | 13 | 0.325 |
| p9 | Cravings | 40 | 1 | 0.025 |
| p93 | Cravings | 40 | 8 | 0.200 |
| p95 | Cravings | 40 | 8 | 0.200 |
| p97 | Cravings | 34 | 18 | 0.529 |
