## Supplementary material for "Optimising supervised machine learning algorithms predicting cigarette cravings and lapses for a smoking cessation just-in-time adaptive intervention (JITAI)": S6 Appendix Sensitivity performance summaries

### 1. Descriptive performance summaries: Sensitivity

#### 1.1. By specific predictors

##### 1.1.1. By prompts per day

Sensitivity by prompts per day

| **Prompts per day** | **n** | **Median** | **Mean** | **Standard deviation** | **25th percentile** | **75th percentile** | **Minimum** | **Maximum** |
| --- | --- | --- | --- | --- | --- | --- | --- | --- |
| 16 | 450 | 0.471 | 0.479 | 0.367 | 0.111 | 0.833 | 0.000 | 1.000 |
| 6 | 436 | 0.500 | 0.507 | 0.352 | 0.222 | 0.843 | 0.000 | 1.000 |
| 5 | 430 | 0.425 | 0.465 | 0.378 | 0.098 | 0.862 | 0.000 | 1.000 |
| 4 | 446 | 0.462 | 0.470 | 0.384 | 0.000 | 0.889 | 0.000 | 1.000 |
| 3 | 442 | 0.519 | 0.501 | 0.387 | 0.000 | 0.889 | 0.000 | 1.000 |

##### 1.1.2. By prompts per day - proportion below threshold

Sensitivity by prompts per day (proportion below acceptable threshold of 0.7)

| **Prompts per day** | **n** | **N below threshold** | **Proportion below threshold** |
| --- | --- | --- | --- |
| 16 | 450 | 286 | 0.636 |
| 6 | 436 | 279 | 0.640 |
| 5 | 430 | 287 | 0.667 |
| 4 | 446 | 302 | 0.677 |
| 3 | 442 | 273 | 0.618 |

Sensitivity by prompts per day × outcome (outcome: lapses)

| **Prompts per day** | **Outcome** | **n** | **Median** | **Mean** | **Standard deviation** | **25th percentile** | **75th percentile** | **Minimum** | **Maximum** |
| --- | --- | --- | --- | --- | --- | --- | --- | --- | --- |
| 16 | Lapses | 158 | 0.333 | 0.412 | 0.355 | 0.097 | 0.754 | 0.000 | 1.000 |
| 6 | Lapses | 156 | 0.536 | 0.526 | 0.351 | 0.250 | 0.806 | 0.000 | 1.000 |
| 5 | Lapses | 156 | 0.433 | 0.500 | 0.371 | 0.143 | 0.914 | 0.000 | 1.000 |
| 4 | Lapses | 156 | 0.531 | 0.531 | 0.368 | 0.244 | 0.933 | 0.000 | 1.000 |
| 3 | Lapses | 154 | 0.608 | 0.570 | 0.330 | 0.381 | 0.800 | 0.000 | 1.000 |

Sensitivity by prompts per day × outcome (outcome: cravings)

| **Prompts per day** | **Outcome** | **n** | **Median** | **Mean** | **Standard deviation** | **25th percentile** | **75th percentile** | **Minimum** | **Maximum** |
| --- | --- | --- | --- | --- | --- | --- | --- | --- | --- |
| 16 | Cravings | 292 | 0.579 | 0.516 | 0.369 | 0.113 | 0.861 | 0.000 | 1.000 |
| 6 | Cravings | 280 | 0.455 | 0.497 | 0.353 | 0.200 | 0.854 | 0.000 | 1.000 |
| 5 | Cravings | 274 | 0.400 | 0.446 | 0.381 | 0.000 | 0.846 | 0.000 | 1.000 |
| 4 | Cravings | 290 | 0.400 | 0.438 | 0.389 | 0.000 | 0.833 | 0.000 | 1.000 |
| 3 | Cravings | 288 | 0.429 | 0.463 | 0.410 | 0.000 | 0.899 | 0.000 | 1.000 |

Sensitivity by prompts per day × outcome (proportion below acceptable threshold of 0.7) (outcome: lapses)

| **Prompts per day** | **Outcome** | **n** | **N below threshold** | **Proportion below threshold** |
| --- | --- | --- | --- | --- |
| 16 | Lapses | 158 | 110 | 0.696 |
| 6 | Lapses | 156 | 96 | 0.615 |
| 5 | Lapses | 156 | 99 | 0.635 |
| 4 | Lapses | 156 | 103 | 0.660 |
| 3 | Lapses | 154 | 93 | 0.604 |

Sensitivity by prompts per day × outcome (proportion below acceptable threshold of 0.7) (outcome: cravings)

| **Prompts per day** | **Outcome** | **n** | **N below threshold** | **Proportion below threshold** |
| --- | --- | --- | --- | --- |
| 16 | Cravings | 292 | 176 | 0.603 |
| 6 | Cravings | 280 | 183 | 0.654 |
| 5 | Cravings | 274 | 188 | 0.686 |
| 4 | Cravings | 290 | 199 | 0.686 |
| 3 | Cravings | 288 | 180 | 0.625 |

##### 1.1.3. By feature selection

Sensitivity by feature selection

| **Feature selection** | **n** | **Median** | **Mean** | **Standard deviation** | **25th percentile** | **75th percentile** | **Minimum** | **Maximum** |
| --- | --- | --- | --- | --- | --- | --- | --- | --- |
| All features | 1,102 | 0.500 | 0.478 | 0.363 | 0.125 | 0.818 | 0.000 | 1.000 |
| Selected features | 1,102 | 0.500 | 0.491 | 0.385 | 0.070 | 0.895 | 0.000 | 1.000 |

##### 1.1.4. By feature selection - proportion below threshold

Sensitivity by feature selection (proportion below acceptable threshold of 0.7)

| **Feature selection** | **n** | **N below threshold** | **Proportion below threshold** |
| --- | --- | --- | --- |
| All features | 1,102 | 729 | 0.662 |
| Selected features | 1,102 | 698 | 0.633 |

Sensitivity by feature selection × outcome (outcome: lapses)

| **Feature selection** | **Outcome** | **n** | **Median** | **Mean** | **Standard deviation** | **25th percentile** | **75th percentile** | **Minimum** | **Maximum** |
| --- | --- | --- | --- | --- | --- | --- | --- | --- | --- |
| All features | Lapses | 390 | 0.500 | 0.490 | 0.348 | 0.200 | 0.778 | 0.000 | 1.000 |
| Selected features | Lapses | 390 | 0.542 | 0.525 | 0.368 | 0.211 | 0.908 | 0.000 | 1.000 |

Sensitivity by feature selection × outcome (outcome: cravings)

| **Feature selection** | **Outcome** | **n** | **Median** | **Mean** | **Standard deviation** | **25th percentile** | **75th percentile** | **Minimum** | **Maximum** |
| --- | --- | --- | --- | --- | --- | --- | --- | --- | --- |
| All features | Cravings | 712 | 0.464 | 0.472 | 0.371 | 0.094 | 0.833 | 0.000 | 1.000 |
| Selected features | Cravings | 712 | 0.444 | 0.472 | 0.392 | 0.000 | 0.889 | 0.000 | 1.000 |

Sensitivity by feature selection × outcome (proportion below acceptable threshold of 0.7) (outcome: lapses)

| **Feature selection** | **Outcome** | **n** | **N below threshold** | **Proportion below threshold** |
| --- | --- | --- | --- | --- |
| All features | Lapses | 390 | 259 | 0.664 |
| Selected features | Lapses | 390 | 242 | 0.621 |

Sensitivity by feature selection × outcome (proportion below acceptable threshold of 0.7) (outcome: cravings)

| **Feature selection** | **Outcome** | **n** | **N below threshold** | **Proportion below threshold** |
| --- | --- | --- | --- | --- |
| All features | Cravings | 712 | 470 | 0.660 |
| Selected features | Cravings | 712 | 456 | 0.640 |

##### 1.1.5. By share of own data in training set

Sensitivity by share of own data in training set

| **Share of own data in training set** | **n** | **Median** | **Mean** | **Standard deviation** | **25th percentile** | **75th percentile** | **Minimum** | **Maximum** |
| --- | --- | --- | --- | --- | --- | --- | --- | --- |
| None | 602 | 0.417 | 0.431 | 0.349 | 0.062 | 0.722 | 0.000 | 1.000 |
| 10% | 554 | 0.500 | 0.496 | 0.376 | 0.111 | 0.884 | 0.000 | 1.000 |
| 20% | 524 | 0.462 | 0.486 | 0.373 | 0.125 | 0.857 | 0.000 | 1.000 |
| 30% | 524 | 0.542 | 0.533 | 0.394 | 0.122 | 0.959 | 0.000 | 1.000 |

##### 1.1.6. By share of own data in training set - proportion below threshold

Sensitivity by share of own data in training set (proportion below acceptable threshold of 0.7)

| **Share of own data in training set** | **n** | **N below threshold** | **Proportion below threshold** |
| --- | --- | --- | --- |
| None | 602 | 439 | 0.729 |
| 10% | 554 | 353 | 0.637 |
| 20% | 524 | 337 | 0.643 |
| 30% | 524 | 298 | 0.569 |

Sensitivity by share of own data in training set × outcome (outcome: lapses)

| **Share of own data in training set** | **Outcome** | **n** | **Median** | **Mean** | **Standard deviation** | **25th percentile** | **75th percentile** | **Minimum** | **Maximum** |
| --- | --- | --- | --- | --- | --- | --- | --- | --- | --- |
| None | Lapses | 242 | 0.333 | 0.384 | 0.345 | 0.000 | 0.667 | 0.000 | 1.000 |
| 10% | Lapses | 198 | 0.561 | 0.508 | 0.373 | 0.167 | 0.845 | 0.000 | 1.000 |
| 20% | Lapses | 170 | 0.542 | 0.585 | 0.312 | 0.333 | 0.896 | 0.000 | 1.000 |
| 30% | Lapses | 170 | 0.667 | 0.606 | 0.354 | 0.346 | 1.000 | 0.000 | 1.000 |

Sensitivity by share of own data in training set × outcome (outcome: cravings)

| **Share of own data in training set** | **Outcome** | **n** | **Median** | **Mean** | **Standard deviation** | **25th percentile** | **75th percentile** | **Minimum** | **Maximum** |
| --- | --- | --- | --- | --- | --- | --- | --- | --- | --- |
| None | Cravings | 360 | 0.486 | 0.463 | 0.348 | 0.111 | 0.757 | 0.000 | 1.000 |
| 10% | Cravings | 356 | 0.500 | 0.489 | 0.378 | 0.108 | 0.886 | 0.000 | 1.000 |
| 20% | Cravings | 354 | 0.375 | 0.438 | 0.391 | 0.000 | 0.850 | 0.000 | 1.000 |
| 30% | Cravings | 354 | 0.500 | 0.498 | 0.407 | 0.000 | 0.944 | 0.000 | 1.000 |

Sensitivity by share of own data in training set × outcome (proportion below acceptable threshold of 0.7) (outcome: lapses)

| **Share of own data in training set** | **Outcome** | **n** | **N below threshold** | **Proportion below threshold** |
| --- | --- | --- | --- | --- |
| None | Lapses | 242 | 189 | 0.781 |
| 10% | Lapses | 198 | 123 | 0.621 |
| 20% | Lapses | 170 | 100 | 0.588 |
| 30% | Lapses | 170 | 89 | 0.524 |

Sensitivity by share of own data in training set × outcome (proportion below acceptable threshold of 0.7) (outcome: cravings)

| **Share of own data in training set** | **Outcome** | **n** | **N below threshold** | **Proportion below threshold** |
| --- | --- | --- | --- | --- |
| None | Cravings | 360 | 250 | 0.694 |
| 10% | Cravings | 356 | 230 | 0.646 |
| 20% | Cravings | 354 | 237 | 0.669 |
| 30% | Cravings | 354 | 209 | 0.590 |

##### 1.1.7. By outcome

Sensitivity by outcome

| **Outcome** | **n** | **Median** | **Mean** | **Standard deviation** | **25th percentile** | **75th percentile** | **Minimum** | **Maximum** |
| --- | --- | --- | --- | --- | --- | --- | --- | --- |
| Lapses | 780 | 0.500 | 0.507 | 0.358 | 0.200 | 0.833 | 0.000 | 1.000 |
| Cravings | 1,424 | 0.455 | 0.472 | 0.382 | 0.025 | 0.867 | 0.000 | 1.000 |

##### 1.1.8. By outcome - proportion below threshold

Sensitivity by outcome (proportion below acceptable threshold of 0.7)

| **Outcome** | **n** | **N below threshold** | **Proportion below threshold** |
| --- | --- | --- | --- |
| Lapses | 780 | 501 | 0.642 |
| Cravings | 1,424 | 926 | 0.650 |

#### 1.2. By specification

##### 1.2.1. By specification

Sensitivity by specification (prompts per day × use of feature selection × share of own data in training set × outcome)

| **Prompts per day** | **Feature selection** | **Share of own data in training set** | **Outcome** | **n** | **Median** | **Mean** | **Standard deviation** | **25th percentile** | **75th percentile** | **Minimum** | **Maximum** |
| --- | --- | --- | --- | --- | --- | --- | --- | --- | --- | --- | --- |
| 16 | All features | None | Lapses | 25 | 0.219 | 0.285 | 0.316 | 0.000 | 0.400 | 0.000 | 1.000 |
| 16 | All features | None | Cravings | 37 | 0.588 | 0.511 | 0.337 | 0.143 | 0.762 | 0.000 | 1.000 |
| 16 | All features | 10% | Lapses | 20 | 0.308 | 0.397 | 0.365 | 0.107 | 0.696 | 0.000 | 1.000 |
| 16 | All features | 10% | Cravings | 37 | 0.600 | 0.526 | 0.371 | 0.174 | 0.872 | 0.000 | 1.000 |
| 16 | All features | 20% | Lapses | 17 | 0.333 | 0.432 | 0.341 | 0.176 | 0.700 | 0.000 | 1.000 |
| 16 | All features | 20% | Cravings | 36 | 0.582 | 0.529 | 0.380 | 0.117 | 0.869 | 0.000 | 1.000 |
| 16 | All features | 30% | Lapses | 17 | 0.333 | 0.432 | 0.374 | 0.056 | 0.800 | 0.000 | 1.000 |
| 16 | All features | 30% | Cravings | 36 | 0.595 | 0.535 | 0.379 | 0.197 | 0.866 | 0.000 | 1.000 |
| 16 | Selected features | None | Lapses | 25 | 0.324 | 0.384 | 0.358 | 0.000 | 0.700 | 0.000 | 1.000 |
| 16 | Selected features | None | Cravings | 37 | 0.583 | 0.516 | 0.349 | 0.167 | 0.812 | 0.000 | 1.000 |
| 16 | Selected features | 10% | Lapses | 20 | 0.333 | 0.444 | 0.378 | 0.125 | 0.725 | 0.000 | 1.000 |
| 16 | Selected features | 10% | Cravings | 37 | 0.591 | 0.508 | 0.387 | 0.091 | 0.890 | 0.000 | 1.000 |
| 16 | Selected features | 20% | Lapses | 17 | 0.500 | 0.492 | 0.355 | 0.207 | 0.800 | 0.000 | 1.000 |
| 16 | Selected features | 20% | Cravings | 36 | 0.545 | 0.504 | 0.385 | 0.047 | 0.855 | 0.000 | 1.000 |
| 16 | Selected features | 30% | Lapses | 17 | 0.450 | 0.497 | 0.378 | 0.222 | 0.857 | 0.000 | 1.000 |
| 16 | Selected features | 30% | Cravings | 36 | 0.437 | 0.498 | 0.396 | 0.062 | 0.897 | 0.000 | 1.000 |
| 6 | All features | None | Lapses | 24 | 0.314 | 0.412 | 0.365 | 0.000 | 0.688 | 0.000 | 1.000 |
| 6 | All features | None | Cravings | 35 | 0.538 | 0.509 | 0.298 | 0.317 | 0.710 | 0.000 | 1.000 |
| 6 | All features | 10% | Lapses | 20 | 0.574 | 0.519 | 0.362 | 0.237 | 0.760 | 0.000 | 1.000 |
| 6 | All features | 10% | Cravings | 35 | 0.545 | 0.522 | 0.342 | 0.255 | 0.861 | 0.000 | 1.000 |
| 6 | All features | 20% | Lapses | 17 | 0.500 | 0.572 | 0.296 | 0.333 | 0.750 | 0.000 | 1.000 |
| 6 | All features | 20% | Cravings | 35 | 0.364 | 0.453 | 0.372 | 0.106 | 0.819 | 0.000 | 1.000 |
| 6 | All features | 30% | Lapses | 17 | 0.667 | 0.582 | 0.330 | 0.400 | 0.750 | 0.000 | 1.000 |
| 6 | All features | 30% | Cravings | 35 | 0.500 | 0.536 | 0.374 | 0.261 | 0.920 | 0.000 | 1.000 |
| 6 | Selected features | None | Lapses | 24 | 0.406 | 0.437 | 0.380 | 0.000 | 0.771 | 0.000 | 1.000 |
| 6 | Selected features | None | Cravings | 35 | 0.489 | 0.511 | 0.326 | 0.357 | 0.737 | 0.000 | 1.000 |
| 6 | Selected features | 10% | Lapses | 20 | 0.667 | 0.565 | 0.391 | 0.188 | 0.945 | 0.000 | 1.000 |
| 6 | Selected features | 10% | Cravings | 35 | 0.455 | 0.497 | 0.345 | 0.298 | 0.785 | 0.000 | 1.000 |
| 6 | Selected features | 20% | Lapses | 17 | 0.500 | 0.546 | 0.294 | 0.333 | 0.750 | 0.000 | 1.000 |
| 6 | Selected features | 20% | Cravings | 35 | 0.281 | 0.435 | 0.392 | 0.087 | 0.877 | 0.000 | 1.000 |
| 6 | Selected features | 30% | Lapses | 17 | 0.700 | 0.651 | 0.348 | 0.400 | 1.000 | 0.000 | 1.000 |
| 6 | Selected features | 30% | Cravings | 35 | 0.429 | 0.511 | 0.387 | 0.191 | 0.943 | 0.000 | 1.000 |
| 5 | All features | None | Lapses | 24 | 0.236 | 0.326 | 0.340 | 0.000 | 0.514 | 0.000 | 1.000 |
| 5 | All features | None | Cravings | 35 | 0.500 | 0.456 | 0.339 | 0.155 | 0.764 | 0.000 | 1.000 |
| 5 | All features | 10% | Lapses | 20 | 0.343 | 0.412 | 0.378 | 0.000 | 0.768 | 0.000 | 1.000 |
| 5 | All features | 10% | Cravings | 34 | 0.381 | 0.435 | 0.355 | 0.159 | 0.773 | 0.000 | 1.000 |
| 5 | All features | 20% | Lapses | 17 | 0.500 | 0.593 | 0.299 | 0.400 | 0.882 | 0.125 | 1.000 |
| 5 | All features | 20% | Cravings | 34 | 0.411 | 0.417 | 0.364 | 0.028 | 0.673 | 0.000 | 1.000 |
| 5 | All features | 30% | Lapses | 17 | 0.778 | 0.694 | 0.313 | 0.400 | 1.000 | 0.143 | 1.000 |
| 5 | All features | 30% | Cravings | 34 | 0.437 | 0.439 | 0.408 | 0.000 | 0.906 | 0.000 | 1.000 |
| 5 | Selected features | None | Lapses | 24 | 0.333 | 0.391 | 0.352 | 0.062 | 0.698 | 0.000 | 1.000 |
| 5 | Selected features | None | Cravings | 35 | 0.333 | 0.429 | 0.380 | 0.047 | 0.789 | 0.000 | 1.000 |
| 5 | Selected features | 10% | Lapses | 20 | 0.469 | 0.442 | 0.393 | 0.000 | 0.762 | 0.000 | 1.000 |
| 5 | Selected features | 10% | Cravings | 34 | 0.407 | 0.470 | 0.402 | 0.051 | 0.874 | 0.000 | 1.000 |
| 5 | Selected features | 20% | Lapses | 17 | 0.750 | 0.640 | 0.366 | 0.375 | 1.000 | 0.000 | 1.000 |
| 5 | Selected features | 20% | Cravings | 34 | 0.388 | 0.438 | 0.406 | 0.000 | 0.913 | 0.000 | 1.000 |
| 5 | Selected features | 30% | Lapses | 17 | 0.750 | 0.648 | 0.380 | 0.400 | 1.000 | 0.000 | 1.000 |
| 5 | Selected features | 30% | Cravings | 34 | 0.397 | 0.481 | 0.425 | 0.000 | 1.000 | 0.000 | 1.000 |
| 4 | All features | None | Lapses | 24 | 0.250 | 0.356 | 0.339 | 0.000 | 0.667 | 0.000 | 0.955 |
| 4 | All features | None | Cravings | 37 | 0.489 | 0.459 | 0.374 | 0.062 | 0.800 | 0.000 | 1.000 |
| 4 | All features | 10% | Lapses | 20 | 0.392 | 0.482 | 0.358 | 0.243 | 0.812 | 0.000 | 1.000 |
| 4 | All features | 10% | Cravings | 36 | 0.458 | 0.446 | 0.366 | 0.083 | 0.710 | 0.000 | 1.000 |
| 4 | All features | 20% | Lapses | 17 | 0.500 | 0.639 | 0.307 | 0.385 | 1.000 | 0.190 | 1.000 |
| 4 | All features | 20% | Cravings | 36 | 0.200 | 0.379 | 0.384 | 0.000 | 0.712 | 0.000 | 1.000 |
| 4 | All features | 30% | Lapses | 17 | 0.667 | 0.593 | 0.375 | 0.385 | 0.933 | 0.000 | 1.000 |
| 4 | All features | 30% | Cravings | 36 | 0.500 | 0.485 | 0.410 | 0.000 | 0.941 | 0.000 | 1.000 |
| 4 | Selected features | None | Lapses | 24 | 0.417 | 0.426 | 0.363 | 0.000 | 0.673 | 0.000 | 1.000 |
| 4 | Selected features | None | Cravings | 37 | 0.500 | 0.452 | 0.376 | 0.000 | 0.750 | 0.000 | 1.000 |
| 4 | Selected features | 10% | Lapses | 20 | 0.679 | 0.590 | 0.396 | 0.243 | 0.967 | 0.000 | 1.000 |
| 4 | Selected features | 10% | Cravings | 36 | 0.360 | 0.445 | 0.401 | 0.000 | 0.894 | 0.000 | 1.000 |
| 4 | Selected features | 20% | Lapses | 17 | 0.692 | 0.649 | 0.347 | 0.375 | 1.000 | 0.000 | 1.000 |
| 4 | Selected features | 20% | Cravings | 36 | 0.293 | 0.389 | 0.403 | 0.000 | 0.818 | 0.000 | 1.000 |
| 4 | Selected features | 30% | Lapses | 17 | 0.667 | 0.630 | 0.385 | 0.400 | 1.000 | 0.000 | 1.000 |
| 4 | Selected features | 30% | Cravings | 36 | 0.388 | 0.447 | 0.425 | 0.000 | 0.950 | 0.000 | 1.000 |
| 3 | All features | None | Lapses | 24 | 0.500 | 0.409 | 0.322 | 0.000 | 0.644 | 0.000 | 0.967 |
| 3 | All features | None | Cravings | 36 | 0.388 | 0.397 | 0.352 | 0.000 | 0.681 | 0.000 | 1.000 |
| 3 | All features | 10% | Lapses | 19 | 0.667 | 0.622 | 0.334 | 0.460 | 0.869 | 0.000 | 1.000 |
| 3 | All features | 10% | Cravings | 36 | 0.444 | 0.480 | 0.390 | 0.031 | 0.886 | 0.000 | 1.000 |
| 3 | All features | 20% | Lapses | 17 | 0.667 | 0.660 | 0.182 | 0.500 | 0.750 | 0.333 | 1.000 |
| 3 | All features | 20% | Cravings | 36 | 0.388 | 0.412 | 0.412 | 0.000 | 0.830 | 0.000 | 1.000 |
| 3 | All features | 30% | Lapses | 17 | 0.750 | 0.692 | 0.272 | 0.500 | 0.909 | 0.000 | 1.000 |
| 3 | All features | 30% | Cravings | 36 | 0.536 | 0.505 | 0.432 | 0.000 | 1.000 | 0.000 | 1.000 |
| 3 | Selected features | None | Lapses | 24 | 0.429 | 0.415 | 0.345 | 0.000 | 0.671 | 0.000 | 1.000 |
| 3 | Selected features | None | Cravings | 36 | 0.336 | 0.394 | 0.354 | 0.000 | 0.695 | 0.000 | 1.000 |
| 3 | Selected features | 10% | Lapses | 19 | 0.667 | 0.615 | 0.363 | 0.345 | 0.948 | 0.000 | 1.000 |
| 3 | Selected features | 10% | Cravings | 36 | 0.652 | 0.554 | 0.437 | 0.000 | 1.000 | 0.000 | 1.000 |
| 3 | Selected features | 20% | Lapses | 17 | 0.571 | 0.630 | 0.293 | 0.462 | 0.938 | 0.000 | 1.000 |
| 3 | Selected features | 20% | Cravings | 36 | 0.211 | 0.423 | 0.431 | 0.000 | 0.878 | 0.000 | 1.000 |
| 3 | Selected features | 30% | Lapses | 17 | 0.750 | 0.639 | 0.376 | 0.429 | 1.000 | 0.000 | 1.000 |
| 3 | Selected features | 30% | Cravings | 36 | 0.690 | 0.541 | 0.464 | 0.000 | 1.000 | 0.000 | 1.000 |

##### 1.2.2. By specification - proportion below threshold

Sensitivity by specification (prompts per day × use of feature selection × share of own data in training set × outcome) - proportion below acceptable threshold of 0.7

| **Prompts per day** | **Feature selection** | **Share of own data in training set** | **Outcome** | **n** | **N below threshold** | **Proportion below threshold** |
| --- | --- | --- | --- | --- | --- | --- |
| 16 | All features | None | Lapses | 25 | 21 | 0.840 |
| 16 | All features | None | Cravings | 37 | 25 | 0.676 |
| 16 | All features | 10% | Lapses | 20 | 15 | 0.750 |
| 16 | All features | 10% | Cravings | 37 | 23 | 0.622 |
| 16 | All features | 20% | Lapses | 17 | 12 | 0.706 |
| 16 | All features | 20% | Cravings | 36 | 20 | 0.556 |
| 16 | All features | 30% | Lapses | 17 | 11 | 0.647 |
| 16 | All features | 30% | Cravings | 36 | 20 | 0.556 |
| 16 | Selected features | None | Lapses | 25 | 18 | 0.720 |
| 16 | Selected features | None | Cravings | 37 | 23 | 0.622 |
| 16 | Selected features | 10% | Lapses | 20 | 13 | 0.650 |
| 16 | Selected features | 10% | Cravings | 37 | 23 | 0.622 |
| 16 | Selected features | 20% | Lapses | 17 | 10 | 0.588 |
| 16 | Selected features | 20% | Cravings | 36 | 22 | 0.611 |
| 16 | Selected features | 30% | Lapses | 17 | 10 | 0.588 |
| 16 | Selected features | 30% | Cravings | 36 | 20 | 0.556 |
| 6 | All features | None | Lapses | 24 | 18 | 0.750 |
| 6 | All features | None | Cravings | 35 | 24 | 0.686 |
| 6 | All features | 10% | Lapses | 20 | 11 | 0.550 |
| 6 | All features | 10% | Cravings | 35 | 23 | 0.657 |
| 6 | All features | 20% | Lapses | 17 | 10 | 0.588 |
| 6 | All features | 20% | Cravings | 35 | 23 | 0.657 |
| 6 | All features | 30% | Lapses | 17 | 9 | 0.529 |
| 6 | All features | 30% | Cravings | 35 | 21 | 0.600 |
| 6 | Selected features | None | Lapses | 24 | 17 | 0.708 |
| 6 | Selected features | None | Cravings | 35 | 24 | 0.686 |
| 6 | Selected features | 10% | Lapses | 20 | 11 | 0.550 |
| 6 | Selected features | 10% | Cravings | 35 | 23 | 0.657 |
| 6 | Selected features | 20% | Lapses | 17 | 12 | 0.706 |
| 6 | Selected features | 20% | Cravings | 35 | 24 | 0.686 |
| 6 | Selected features | 30% | Lapses | 17 | 8 | 0.471 |
| 6 | Selected features | 30% | Cravings | 35 | 21 | 0.600 |
| 5 | All features | None | Lapses | 24 | 20 | 0.833 |
| 5 | All features | None | Cravings | 35 | 25 | 0.714 |
| 5 | All features | 10% | Lapses | 20 | 14 | 0.700 |
| 5 | All features | 10% | Cravings | 34 | 24 | 0.706 |
| 5 | All features | 20% | Lapses | 17 | 10 | 0.588 |
| 5 | All features | 20% | Cravings | 34 | 26 | 0.765 |
| 5 | All features | 30% | Lapses | 17 | 7 | 0.412 |
| 5 | All features | 30% | Cravings | 34 | 23 | 0.676 |
| 5 | Selected features | None | Lapses | 24 | 18 | 0.750 |
| 5 | Selected features | None | Cravings | 35 | 25 | 0.714 |
| 5 | Selected features | 10% | Lapses | 20 | 14 | 0.700 |
| 5 | Selected features | 10% | Cravings | 34 | 22 | 0.647 |
| 5 | Selected features | 20% | Lapses | 17 | 8 | 0.471 |
| 5 | Selected features | 20% | Cravings | 34 | 23 | 0.676 |
| 5 | Selected features | 30% | Lapses | 17 | 8 | 0.471 |
| 5 | Selected features | 30% | Cravings | 34 | 20 | 0.588 |
| 4 | All features | None | Lapses | 24 | 20 | 0.833 |
| 4 | All features | None | Cravings | 37 | 25 | 0.676 |
| 4 | All features | 10% | Lapses | 20 | 14 | 0.700 |
| 4 | All features | 10% | Cravings | 36 | 26 | 0.722 |
| 4 | All features | 20% | Lapses | 17 | 10 | 0.588 |
| 4 | All features | 20% | Cravings | 36 | 27 | 0.750 |
| 4 | All features | 30% | Lapses | 17 | 10 | 0.588 |
| 4 | All features | 30% | Cravings | 36 | 22 | 0.611 |
| 4 | Selected features | None | Lapses | 24 | 19 | 0.792 |
| 4 | Selected features | None | Cravings | 37 | 25 | 0.676 |
| 4 | Selected features | 10% | Lapses | 20 | 11 | 0.550 |
| 4 | Selected features | 10% | Cravings | 36 | 25 | 0.694 |
| 4 | Selected features | 20% | Lapses | 17 | 9 | 0.529 |
| 4 | Selected features | 20% | Cravings | 36 | 25 | 0.694 |
| 4 | Selected features | 30% | Lapses | 17 | 10 | 0.588 |
| 4 | Selected features | 30% | Cravings | 36 | 24 | 0.667 |
| 3 | All features | None | Lapses | 24 | 20 | 0.833 |
| 3 | All features | None | Cravings | 36 | 27 | 0.750 |
| 3 | All features | 10% | Lapses | 19 | 10 | 0.526 |
| 3 | All features | 10% | Cravings | 36 | 22 | 0.611 |
| 3 | All features | 20% | Lapses | 17 | 9 | 0.529 |
| 3 | All features | 20% | Cravings | 36 | 24 | 0.667 |
| 3 | All features | 30% | Lapses | 17 | 8 | 0.471 |
| 3 | All features | 30% | Cravings | 36 | 20 | 0.556 |
| 3 | Selected features | None | Lapses | 24 | 18 | 0.750 |
| 3 | Selected features | None | Cravings | 36 | 27 | 0.750 |
| 3 | Selected features | 10% | Lapses | 19 | 10 | 0.526 |
| 3 | Selected features | 10% | Cravings | 36 | 19 | 0.528 |
| 3 | Selected features | 20% | Lapses | 17 | 10 | 0.588 |
| 3 | Selected features | 20% | Cravings | 36 | 23 | 0.639 |
| 3 | Selected features | 30% | Lapses | 17 | 8 | 0.471 |
| 3 | Selected features | 30% | Cravings | 36 | 18 | 0.500 |

#### 1.3. By participant

##### 1.3.1. By participant (overall)

Sensitivity by participant

| **Participant** | **n** | **Median** | **Mean** | **Standard deviation** | **25th percentile** | **75th percentile** | **Minimum** | **Maximum** |
| --- | --- | --- | --- | --- | --- | --- | --- | --- |
| p1 | 80 | 0.364 | 0.433 | 0.280 | 0.222 | 0.618 | 0.000 | 1.000 |
| p10 | 40 | 0.000 | 0.024 | 0.056 | 0.000 | 0.014 | 0.000 | 0.236 |
| p101 | 80 | 0.500 | 0.479 | 0.303 | 0.217 | 0.760 | 0.000 | 1.000 |
| p107 | 80 | 0.400 | 0.337 | 0.290 | 0.000 | 0.507 | 0.000 | 1.000 |
| p12 | 80 | 0.160 | 0.375 | 0.403 | 0.000 | 0.777 | 0.000 | 1.000 |
| p128 | 80 | 0.567 | 0.565 | 0.345 | 0.247 | 0.892 | 0.000 | 1.000 |
| p129 | 40 | 0.000 | 0.075 | 0.183 | 0.000 | 0.098 | 0.000 | 1.000 |
| p131 | 42 | 0.956 | 0.917 | 0.171 | 0.926 | 1.000 | 0.000 | 1.000 |
| p132 | 60 | 0.675 | 0.553 | 0.413 | 0.000 | 0.962 | 0.000 | 1.000 |
| p135 | 80 | 0.679 | 0.631 | 0.256 | 0.443 | 0.818 | 0.000 | 1.000 |
| p141 | 80 | 0.609 | 0.581 | 0.353 | 0.271 | 0.959 | 0.000 | 1.000 |
| p144 | 80 | 0.608 | 0.512 | 0.246 | 0.308 | 0.692 | 0.000 | 0.893 |
| p147 | 40 | 0.500 | 0.543 | 0.231 | 0.403 | 0.753 | 0.091 | 0.944 |
| p15 | 80 | 0.167 | 0.412 | 0.435 | 0.000 | 0.948 | 0.000 | 1.000 |
| p20 | 40 | 1.000 | 0.999 | 0.005 | 1.000 | 1.000 | 0.966 | 1.000 |
| p21 | 80 | 0.947 | 0.769 | 0.288 | 0.518 | 1.000 | 0.158 | 1.000 |
| p22 | 80 | 0.537 | 0.571 | 0.271 | 0.358 | 0.800 | 0.000 | 1.000 |
| p27 | 32 | 0.000 | 0.000 | 0.000 | 0.000 | 0.000 | 0.000 | 0.000 |
| p30 | 50 | 0.722 | 0.614 | 0.411 | 0.152 | 1.000 | 0.000 | 1.000 |
| p39 | 40 | 0.723 | 0.685 | 0.194 | 0.520 | 0.835 | 0.250 | 1.000 |
| p44 | 40 | 0.250 | 0.296 | 0.191 | 0.143 | 0.407 | 0.000 | 0.714 |
| p59 | 40 | 0.369 | 0.319 | 0.214 | 0.135 | 0.510 | 0.000 | 0.654 |
| p6 | 80 | 0.971 | 0.841 | 0.228 | 0.769 | 1.000 | 0.222 | 1.000 |
| p61 | 80 | 0.400 | 0.334 | 0.269 | 0.000 | 0.500 | 0.000 | 0.750 |
| p62 | 60 | 0.000 | 0.156 | 0.288 | 0.000 | 0.188 | 0.000 | 1.000 |
| p63 | 80 | 0.667 | 0.540 | 0.306 | 0.318 | 0.750 | 0.000 | 1.000 |
| p67 | 42 | 0.000 | 0.000 | 0.000 | 0.000 | 0.000 | 0.000 | 0.000 |
| p7 | 80 | 0.268 | 0.298 | 0.297 | 0.000 | 0.433 | 0.000 | 1.000 |
| p70 | 6 | 0.000 | 0.000 | 0.000 | 0.000 | 0.000 | 0.000 | 0.000 |
| p74 | 50 | 0.322 | 0.365 | 0.328 | 0.034 | 0.609 | 0.000 | 1.000 |
| p78 | 40 | 1.000 | 0.889 | 0.200 | 0.906 | 1.000 | 0.244 | 1.000 |
| p79 | 40 | 0.000 | 0.132 | 0.258 | 0.000 | 0.125 | 0.000 | 1.000 |
| p87 | 58 | 1.000 | 0.784 | 0.405 | 0.953 | 1.000 | 0.000 | 1.000 |
| p9 | 50 | 0.276 | 0.304 | 0.263 | 0.075 | 0.421 | 0.000 | 1.000 |
| p93 | 40 | 0.891 | 0.866 | 0.105 | 0.814 | 0.942 | 0.591 | 1.000 |
| p95 | 80 | 0.627 | 0.655 | 0.323 | 0.356 | 1.000 | 0.000 | 1.000 |
| p97 | 74 | 0.000 | 0.236 | 0.301 | 0.000 | 0.458 | 0.000 | 1.000 |

##### 1.3.2. By participant (overall) - proportion below threshold

Sensitivity by participant (proportion below acceptable threshold of 0.7)

| **Participant** | **n** | **N below threshold** | **Proportion below threshold** |
| --- | --- | --- | --- |
| p1 | 80 | 64 | 0.800 |
| p10 | 40 | 40 | 1.000 |
| p101 | 80 | 53 | 0.662 |
| p107 | 80 | 71 | 0.887 |
| p12 | 80 | 57 | 0.713 |
| p128 | 80 | 44 | 0.550 |
| p129 | 40 | 39 | 0.975 |
| p131 | 42 | 2 | 0.048 |
| p132 | 60 | 36 | 0.600 |
| p135 | 80 | 42 | 0.525 |
| p141 | 80 | 42 | 0.525 |
| p144 | 80 | 66 | 0.825 |
| p147 | 40 | 28 | 0.700 |
| p15 | 80 | 51 | 0.637 |
| p20 | 40 | 0 | 0.000 |
| p21 | 80 | 26 | 0.325 |
| p22 | 80 | 49 | 0.613 |
| p27 | 32 | 32 | 1.000 |
| p30 | 50 | 25 | 0.500 |
| p39 | 40 | 18 | 0.450 |
| p44 | 40 | 38 | 0.950 |
| p59 | 40 | 40 | 1.000 |
| p6 | 80 | 16 | 0.200 |
| p61 | 80 | 69 | 0.863 |
| p62 | 60 | 55 | 0.917 |
| p63 | 80 | 51 | 0.637 |
| p67 | 42 | 42 | 1.000 |
| p7 | 80 | 71 | 0.887 |
| p70 | 6 | 6 | 1.000 |
| p74 | 50 | 40 | 0.800 |
| p78 | 40 | 6 | 0.150 |
| p79 | 40 | 38 | 0.950 |
| p87 | 58 | 12 | 0.207 |
| p9 | 50 | 46 | 0.920 |
| p93 | 40 | 3 | 0.075 |
| p95 | 80 | 41 | 0.512 |
| p97 | 74 | 68 | 0.919 |

Sensitivity by participant × outcome (outcome: lapses)

| **Participant** | **Outcome** | **n** | **Median** | **Mean** | **Standard deviation** | **25th percentile** | **75th percentile** | **Minimum** | **Maximum** |
| --- | --- | --- | --- | --- | --- | --- | --- | --- | --- |
| p1 | Lapses | 40 | 0.450 | 0.529 | 0.291 | 0.333 | 0.824 | 0.000 | 1.000 |
| p101 | Lapses | 40 | 0.211 | 0.231 | 0.178 | 0.122 | 0.333 | 0.000 | 0.571 |
| p107 | Lapses | 40 | 0.000 | 0.175 | 0.247 | 0.000 | 0.500 | 0.000 | 0.667 |
| p12 | Lapses | 40 | 0.785 | 0.716 | 0.289 | 0.529 | 1.000 | 0.000 | 1.000 |
| p128 | Lapses | 40 | 0.780 | 0.669 | 0.308 | 0.453 | 0.908 | 0.042 | 1.000 |
| p131 | Lapses | 2 | 0.250 | 0.250 | 0.354 | 0.125 | 0.375 | 0.000 | 0.500 |
| p132 | Lapses | 20 | 0.000 | 0.000 | 0.000 | 0.000 | 0.000 | 0.000 | 0.000 |
| p135 | Lapses | 40 | 0.700 | 0.670 | 0.239 | 0.500 | 0.822 | 0.062 | 1.000 |
| p141 | Lapses | 40 | 0.286 | 0.295 | 0.217 | 0.143 | 0.400 | 0.000 | 0.800 |
| p144 | Lapses | 40 | 0.636 | 0.583 | 0.149 | 0.533 | 0.692 | 0.231 | 0.800 |
| p15 | Lapses | 40 | 0.949 | 0.792 | 0.288 | 0.645 | 1.000 | 0.103 | 1.000 |
| p21 | Lapses | 40 | 1.000 | 0.993 | 0.023 | 1.000 | 1.000 | 0.875 | 1.000 |
| p22 | Lapses | 40 | 0.450 | 0.501 | 0.227 | 0.383 | 0.700 | 0.000 | 1.000 |
| p30 | Lapses | 10 | 0.000 | 0.000 | 0.000 | 0.000 | 0.000 | 0.000 | 0.000 |
| p6 | Lapses | 40 | 1.000 | 0.968 | 0.073 | 0.980 | 1.000 | 0.714 | 1.000 |
| p61 | Lapses | 40 | 0.500 | 0.538 | 0.156 | 0.400 | 0.700 | 0.200 | 0.750 |
| p62 | Lapses | 20 | 0.000 | 0.000 | 0.000 | 0.000 | 0.000 | 0.000 | 0.000 |
| p63 | Lapses | 40 | 0.750 | 0.762 | 0.113 | 0.667 | 0.833 | 0.600 | 1.000 |
| p67 | Lapses | 10 | 0.000 | 0.000 | 0.000 | 0.000 | 0.000 | 0.000 | 0.000 |
| p7 | Lapses | 40 | 0.333 | 0.350 | 0.284 | 0.143 | 0.433 | 0.000 | 1.000 |
| p74 | Lapses | 10 | 0.000 | 0.000 | 0.000 | 0.000 | 0.000 | 0.000 | 0.000 |
| p87 | Lapses | 18 | 0.000 | 0.333 | 0.485 | 0.000 | 1.000 | 0.000 | 1.000 |
| p9 | Lapses | 10 | 0.000 | 0.000 | 0.000 | 0.000 | 0.000 | 0.000 | 0.000 |
| p95 | Lapses | 40 | 0.422 | 0.525 | 0.335 | 0.258 | 0.921 | 0.000 | 1.000 |
| p97 | Lapses | 40 | 0.333 | 0.438 | 0.281 | 0.312 | 0.500 | 0.000 | 1.000 |

Sensitivity by participant × outcome (outcome: cravings)

| **Participant** | **Outcome** | **n** | **Median** | **Mean** | **Standard deviation** | **25th percentile** | **75th percentile** | **Minimum** | **Maximum** |
| --- | --- | --- | --- | --- | --- | --- | --- | --- | --- |
| p1 | Cravings | 40 | 0.286 | 0.337 | 0.235 | 0.200 | 0.381 | 0.000 | 0.950 |
| p10 | Cravings | 40 | 0.000 | 0.024 | 0.056 | 0.000 | 0.014 | 0.000 | 0.236 |
| p101 | Cravings | 40 | 0.761 | 0.727 | 0.168 | 0.624 | 0.839 | 0.281 | 1.000 |
| p107 | Cravings | 40 | 0.483 | 0.500 | 0.234 | 0.375 | 0.647 | 0.000 | 1.000 |
| p12 | Cravings | 40 | 0.000 | 0.035 | 0.089 | 0.000 | 0.015 | 0.000 | 0.500 |
| p128 | Cravings | 40 | 0.407 | 0.462 | 0.353 | 0.174 | 0.865 | 0.000 | 1.000 |
| p129 | Cravings | 40 | 0.000 | 0.075 | 0.183 | 0.000 | 0.098 | 0.000 | 1.000 |
| p131 | Cravings | 40 | 0.959 | 0.950 | 0.061 | 0.930 | 1.000 | 0.750 | 1.000 |
| p132 | Cravings | 40 | 0.848 | 0.830 | 0.149 | 0.675 | 0.978 | 0.643 | 1.000 |
| p135 | Cravings | 40 | 0.609 | 0.591 | 0.269 | 0.423 | 0.805 | 0.000 | 1.000 |
| p141 | Cravings | 40 | 0.962 | 0.867 | 0.196 | 0.856 | 0.985 | 0.222 | 1.000 |
| p144 | Cravings | 40 | 0.450 | 0.442 | 0.299 | 0.125 | 0.677 | 0.000 | 0.893 |
| p147 | Cravings | 40 | 0.500 | 0.543 | 0.231 | 0.403 | 0.753 | 0.091 | 0.944 |
| p15 | Cravings | 40 | 0.000 | 0.032 | 0.062 | 0.000 | 0.000 | 0.000 | 0.222 |
| p20 | Cravings | 40 | 1.000 | 0.999 | 0.005 | 1.000 | 1.000 | 0.966 | 1.000 |
| p21 | Cravings | 40 | 0.498 | 0.546 | 0.255 | 0.347 | 0.737 | 0.158 | 1.000 |
| p22 | Cravings | 40 | 0.691 | 0.642 | 0.296 | 0.358 | 0.930 | 0.127 | 1.000 |
| p27 | Cravings | 32 | 0.000 | 0.000 | 0.000 | 0.000 | 0.000 | 0.000 | 0.000 |
| p30 | Cravings | 40 | 1.000 | 0.768 | 0.302 | 0.589 | 1.000 | 0.000 | 1.000 |
| p39 | Cravings | 40 | 0.723 | 0.685 | 0.194 | 0.520 | 0.835 | 0.250 | 1.000 |
| p44 | Cravings | 40 | 0.250 | 0.296 | 0.191 | 0.143 | 0.407 | 0.000 | 0.714 |
| p59 | Cravings | 40 | 0.369 | 0.319 | 0.214 | 0.135 | 0.510 | 0.000 | 0.654 |
| p6 | Cravings | 40 | 0.800 | 0.714 | 0.259 | 0.452 | 0.936 | 0.222 | 1.000 |
| p61 | Cravings | 40 | 0.000 | 0.129 | 0.191 | 0.000 | 0.250 | 0.000 | 0.500 |
| p62 | Cravings | 40 | 0.091 | 0.234 | 0.327 | 0.000 | 0.333 | 0.000 | 1.000 |
| p63 | Cravings | 40 | 0.303 | 0.318 | 0.276 | 0.062 | 0.556 | 0.000 | 0.923 |
| p67 | Cravings | 32 | 0.000 | 0.000 | 0.000 | 0.000 | 0.000 | 0.000 | 0.000 |
| p7 | Cravings | 40 | 0.100 | 0.246 | 0.305 | 0.000 | 0.425 | 0.000 | 1.000 |
| p70 | Cravings | 6 | 0.000 | 0.000 | 0.000 | 0.000 | 0.000 | 0.000 | 0.000 |
| p74 | Cravings | 40 | 0.453 | 0.456 | 0.305 | 0.171 | 0.667 | 0.000 | 1.000 |
| p78 | Cravings | 40 | 1.000 | 0.889 | 0.200 | 0.906 | 1.000 | 0.244 | 1.000 |
| p79 | Cravings | 40 | 0.000 | 0.132 | 0.258 | 0.000 | 0.125 | 0.000 | 1.000 |
| p87 | Cravings | 40 | 1.000 | 0.986 | 0.043 | 0.998 | 1.000 | 0.750 | 1.000 |
| p9 | Cravings | 40 | 0.344 | 0.380 | 0.239 | 0.200 | 0.500 | 0.000 | 1.000 |
| p93 | Cravings | 40 | 0.891 | 0.866 | 0.105 | 0.814 | 0.942 | 0.591 | 1.000 |
| p95 | Cravings | 40 | 0.931 | 0.786 | 0.252 | 0.573 | 1.000 | 0.143 | 1.000 |
| p97 | Cravings | 34 | 0.000 | 0.000 | 0.000 | 0.000 | 0.000 | 0.000 | 0.000 |

Sensitivity by participant × outcome (proportion below acceptable threshold of 0.7) (outcome: lapses)

| **Participant** | **Outcome** | **n** | **N below threshold** | **Proportion below threshold** |
| --- | --- | --- | --- | --- |
| p1 | Lapses | 40 | 28 | 0.700 |
| p101 | Lapses | 40 | 40 | 1.000 |
| p107 | Lapses | 40 | 40 | 1.000 |
| p12 | Lapses | 40 | 17 | 0.425 |
| p128 | Lapses | 40 | 15 | 0.375 |
| p131 | Lapses | 2 | 2 | 1.000 |
| p132 | Lapses | 20 | 20 | 1.000 |
| p135 | Lapses | 40 | 19 | 0.475 |
| p141 | Lapses | 40 | 36 | 0.900 |
| p144 | Lapses | 40 | 36 | 0.900 |
| p15 | Lapses | 40 | 11 | 0.275 |
| p21 | Lapses | 40 | 0 | 0.000 |
| p22 | Lapses | 40 | 28 | 0.700 |
| p30 | Lapses | 10 | 10 | 1.000 |
| p6 | Lapses | 40 | 0 | 0.000 |
| p61 | Lapses | 40 | 29 | 0.725 |
| p62 | Lapses | 20 | 20 | 1.000 |
| p63 | Lapses | 40 | 14 | 0.350 |
| p67 | Lapses | 10 | 10 | 1.000 |
| p7 | Lapses | 40 | 35 | 0.875 |
| p74 | Lapses | 10 | 10 | 1.000 |
| p87 | Lapses | 18 | 12 | 0.667 |
| p9 | Lapses | 10 | 10 | 1.000 |
| p95 | Lapses | 40 | 25 | 0.625 |
| p97 | Lapses | 40 | 34 | 0.850 |

Sensitivity by participant × outcome (proportion below acceptable threshold of 0.7) (outcome: cravings)

| **Participant** | **Outcome** | **n** | **N below threshold** | **Proportion below threshold** |
| --- | --- | --- | --- | --- |
| p1 | Cravings | 40 | 36 | 0.900 |
| p10 | Cravings | 40 | 40 | 1.000 |
| p101 | Cravings | 40 | 13 | 0.325 |
| p107 | Cravings | 40 | 31 | 0.775 |
| p12 | Cravings | 40 | 40 | 1.000 |
| p128 | Cravings | 40 | 29 | 0.725 |
| p129 | Cravings | 40 | 39 | 0.975 |
| p131 | Cravings | 40 | 0 | 0.000 |
| p132 | Cravings | 40 | 16 | 0.400 |
| p135 | Cravings | 40 | 23 | 0.575 |
| p141 | Cravings | 40 | 6 | 0.150 |
| p144 | Cravings | 40 | 30 | 0.750 |
| p147 | Cravings | 40 | 28 | 0.700 |
| p15 | Cravings | 40 | 40 | 1.000 |
| p20 | Cravings | 40 | 0 | 0.000 |
| p21 | Cravings | 40 | 26 | 0.650 |
| p22 | Cravings | 40 | 21 | 0.525 |
| p27 | Cravings | 32 | 32 | 1.000 |
| p30 | Cravings | 40 | 15 | 0.375 |
| p39 | Cravings | 40 | 18 | 0.450 |
| p44 | Cravings | 40 | 38 | 0.950 |
| p59 | Cravings | 40 | 40 | 1.000 |
| p6 | Cravings | 40 | 16 | 0.400 |
| p61 | Cravings | 40 | 40 | 1.000 |
| p62 | Cravings | 40 | 35 | 0.875 |
| p63 | Cravings | 40 | 37 | 0.925 |
| p67 | Cravings | 32 | 32 | 1.000 |
| p7 | Cravings | 40 | 36 | 0.900 |
| p70 | Cravings | 6 | 6 | 1.000 |
| p74 | Cravings | 40 | 30 | 0.750 |
| p78 | Cravings | 40 | 6 | 0.150 |
| p79 | Cravings | 40 | 38 | 0.950 |
| p87 | Cravings | 40 | 0 | 0.000 |
| p9 | Cravings | 40 | 36 | 0.900 |
| p93 | Cravings | 40 | 3 | 0.075 |
| p95 | Cravings | 40 | 16 | 0.400 |
| p97 | Cravings | 34 | 34 | 1.000 |
