## Supplementary material for "Optimising supervised machine learning algorithms predicting cigarette cravings and lapses for a smoking cessation just-in-time adaptive intervention (JITAI)": S7 Appendix Specificity performance summaries

### 1. Descriptive performance summaries: Specificity

#### 1.1. By specific predictors

##### 1.1.1. By prompts per day

Specificity by prompts per day

| **Prompts per day** | **n** | **Median** | **Mean** | **Standard deviation** | **25th percentile** | **75th percentile** | **Minimum** | **Maximum** |
| --- | --- | --- | --- | --- | --- | --- | --- | --- |
| 16 | 450 | 0.878 | 0.716 | 0.338 | 0.565 | 0.975 | 0.000 | 1.000 |
| 6 | 436 | 0.827 | 0.639 | 0.368 | 0.276 | 0.946 | 0.000 | 1.000 |
| 5 | 430 | 0.779 | 0.636 | 0.377 | 0.294 | 0.970 | 0.000 | 1.000 |
| 4 | 446 | 0.785 | 0.613 | 0.387 | 0.193 | 0.960 | 0.000 | 1.000 |
| 3 | 442 | 0.735 | 0.585 | 0.393 | 0.143 | 0.954 | 0.000 | 1.000 |

##### 1.1.2. By prompts per day - proportion below threshold

Specificity by prompts per day (proportion below acceptable threshold of 0.5)

| **Prompts per day** | **n** | **N below threshold** | **Proportion below threshold** |
| --- | --- | --- | --- |
| 16 | 450 | 102 | 0.227 |
| 6 | 436 | 131 | 0.300 |
| 5 | 430 | 136 | 0.316 |
| 4 | 446 | 146 | 0.327 |
| 3 | 442 | 161 | 0.364 |

Specificity by prompts per day × outcome (outcome: lapses)

| **Prompts per day** | **Outcome** | **n** | **Median** | **Mean** | **Standard deviation** | **25th percentile** | **75th percentile** | **Minimum** | **Maximum** |
| --- | --- | --- | --- | --- | --- | --- | --- | --- | --- |
| 16 | Lapses | 158 | 0.942 | 0.783 | 0.322 | 0.773 | 0.984 | 0.000 | 1.000 |
| 6 | Lapses | 156 | 0.845 | 0.640 | 0.384 | 0.185 | 0.961 | 0.000 | 1.000 |
| 5 | Lapses | 156 | 0.821 | 0.639 | 0.378 | 0.258 | 0.962 | 0.000 | 1.000 |
| 4 | Lapses | 156 | 0.785 | 0.595 | 0.405 | 0.104 | 0.967 | 0.000 | 1.000 |
| 3 | Lapses | 154 | 0.750 | 0.615 | 0.368 | 0.182 | 0.926 | 0.000 | 1.000 |

Specificity by prompts per day × outcome (outcome: cravings)

| **Prompts per day** | **Outcome** | **n** | **Median** | **Mean** | **Standard deviation** | **25th percentile** | **75th percentile** | **Minimum** | **Maximum** |
| --- | --- | --- | --- | --- | --- | --- | --- | --- | --- |
| 16 | Cravings | 292 | 0.840 | 0.680 | 0.341 | 0.476 | 0.953 | 0.000 | 1.000 |
| 6 | Cravings | 280 | 0.796 | 0.639 | 0.359 | 0.333 | 0.938 | 0.000 | 1.000 |
| 5 | Cravings | 274 | 0.772 | 0.634 | 0.377 | 0.320 | 0.974 | 0.000 | 1.000 |
| 4 | Cravings | 290 | 0.782 | 0.623 | 0.377 | 0.256 | 0.952 | 0.000 | 1.000 |
| 3 | Cravings | 288 | 0.692 | 0.569 | 0.405 | 0.079 | 0.960 | 0.000 | 1.000 |

Specificity by prompts per day × outcome (proportion below acceptable threshold of 0.5) (outcome: lapses)

| **Prompts per day** | **Outcome** | **n** | **N below threshold** | **Proportion below threshold** |
| --- | --- | --- | --- | --- |
| 16 | Lapses | 158 | 26 | 0.165 |
| 6 | Lapses | 156 | 50 | 0.321 |
| 5 | Lapses | 156 | 48 | 0.308 |
| 4 | Lapses | 156 | 56 | 0.359 |
| 3 | Lapses | 154 | 45 | 0.292 |

Specificity by prompts per day × outcome (proportion below acceptable threshold of 0.5) (outcome: cravings)

| **Prompts per day** | **Outcome** | **n** | **N below threshold** | **Proportion below threshold** |
| --- | --- | --- | --- | --- |
| 16 | Cravings | 292 | 76 | 0.260 |
| 6 | Cravings | 280 | 81 | 0.289 |
| 5 | Cravings | 274 | 88 | 0.321 |
| 4 | Cravings | 290 | 90 | 0.310 |
| 3 | Cravings | 288 | 116 | 0.403 |

##### 1.1.3. By feature selection

Specificity by feature selection

| **Feature selection** | **n** | **Median** | **Mean** | **Standard deviation** | **25th percentile** | **75th percentile** | **Minimum** | **Maximum** |
| --- | --- | --- | --- | --- | --- | --- | --- | --- |
| All features | 1,102 | 0.804 | 0.650 | 0.361 | 0.355 | 0.960 | 0.000 | 1.000 |
| Selected features | 1,102 | 0.811 | 0.626 | 0.389 | 0.183 | 0.963 | 0.000 | 1.000 |

##### 1.1.4. By feature selection - proportion below threshold

Specificity by feature selection (proportion below acceptable threshold of 0.5)

| **Feature selection** | **n** | **N below threshold** | **Proportion below threshold** |
| --- | --- | --- | --- |
| All features | 1,102 | 325 | 0.295 |
| Selected features | 1,102 | 351 | 0.319 |

Specificity by feature selection × outcome (outcome: lapses)

| **Feature selection** | **Outcome** | **n** | **Median** | **Mean** | **Standard deviation** | **25th percentile** | **75th percentile** | **Minimum** | **Maximum** |
| --- | --- | --- | --- | --- | --- | --- | --- | --- | --- |
| All features | Lapses | 390 | 0.826 | 0.673 | 0.356 | 0.444 | 0.969 | 0.000 | 1.000 |
| Selected features | Lapses | 390 | 0.838 | 0.636 | 0.398 | 0.130 | 0.968 | 0.000 | 1.000 |

Specificity by feature selection × outcome (outcome: cravings)

| **Feature selection** | **Outcome** | **n** | **Median** | **Mean** | **Standard deviation** | **25th percentile** | **75th percentile** | **Minimum** | **Maximum** |
| --- | --- | --- | --- | --- | --- | --- | --- | --- | --- |
| All features | Cravings | 712 | 0.789 | 0.637 | 0.363 | 0.333 | 0.951 | 0.000 | 1.000 |
| Selected features | Cravings | 712 | 0.793 | 0.621 | 0.384 | 0.212 | 0.958 | 0.000 | 1.000 |

Specificity by feature selection × outcome (proportion below acceptable threshold of 0.5) (outcome: lapses)

| **Feature selection** | **Outcome** | **n** | **N below threshold** | **Proportion below threshold** |
| --- | --- | --- | --- | --- |
| All features | Lapses | 390 | 104 | 0.267 |
| Selected features | Lapses | 390 | 121 | 0.310 |

Specificity by feature selection × outcome (proportion below acceptable threshold of 0.5) (outcome: cravings)

| **Feature selection** | **Outcome** | **n** | **N below threshold** | **Proportion below threshold** |
| --- | --- | --- | --- | --- |
| All features | Cravings | 712 | 221 | 0.310 |
| Selected features | Cravings | 712 | 230 | 0.323 |

##### 1.1.5. By share of own data in training set

Specificity by share of own data in training set

| **Share of own data in training set** | **n** | **Median** | **Mean** | **Standard deviation** | **25th percentile** | **75th percentile** | **Minimum** | **Maximum** |
| --- | --- | --- | --- | --- | --- | --- | --- | --- |
| None | 602 | 0.830 | 0.709 | 0.306 | 0.546 | 0.955 | 0.000 | 1.000 |
| 10% | 554 | 0.794 | 0.627 | 0.374 | 0.226 | 0.946 | 0.000 | 1.000 |
| 20% | 524 | 0.815 | 0.632 | 0.387 | 0.215 | 0.970 | 0.000 | 1.000 |
| 30% | 524 | 0.750 | 0.574 | 0.420 | 0.024 | 0.969 | 0.000 | 1.000 |

##### 1.1.6. By share of own data in training set - proportion below threshold

Specificity by share of own data in training set (proportion below acceptable threshold of 0.5)

| **Share of own data in training set** | **n** | **N below threshold** | **Proportion below threshold** |
| --- | --- | --- | --- |
| None | 602 | 126 | 0.209 |
| 10% | 554 | 178 | 0.321 |
| 20% | 524 | 167 | 0.319 |
| 30% | 524 | 205 | 0.391 |

Specificity by share of own data in training set × outcome (outcome: lapses)

| **Share of own data in training set** | **Outcome** | **n** | **Median** | **Mean** | **Standard deviation** | **25th percentile** | **75th percentile** | **Minimum** | **Maximum** |
| --- | --- | --- | --- | --- | --- | --- | --- | --- | --- |
| None | Lapses | 242 | 0.864 | 0.742 | 0.308 | 0.634 | 0.974 | 0.000 | 1.000 |
| 10% | Lapses | 198 | 0.817 | 0.649 | 0.371 | 0.300 | 0.959 | 0.000 | 1.000 |
| 20% | Lapses | 170 | 0.800 | 0.609 | 0.398 | 0.118 | 0.956 | 0.000 | 1.000 |
| 30% | Lapses | 170 | 0.805 | 0.583 | 0.430 | 0.042 | 0.973 | 0.000 | 1.000 |

Specificity by share of own data in training set × outcome (outcome: cravings)

| **Share of own data in training set** | **Outcome** | **n** | **Median** | **Mean** | **Standard deviation** | **25th percentile** | **75th percentile** | **Minimum** | **Maximum** |
| --- | --- | --- | --- | --- | --- | --- | --- | --- | --- |
| None | Cravings | 360 | 0.800 | 0.687 | 0.304 | 0.500 | 0.934 | 0.000 | 1.000 |
| 10% | Cravings | 356 | 0.778 | 0.615 | 0.376 | 0.204 | 0.942 | 0.000 | 1.000 |
| 20% | Cravings | 354 | 0.841 | 0.643 | 0.382 | 0.312 | 0.973 | 0.000 | 1.000 |
| 30% | Cravings | 354 | 0.726 | 0.570 | 0.416 | 0.000 | 0.963 | 0.000 | 1.000 |

Specificity by share of own data in training set × outcome (proportion below acceptable threshold of 0.5) (outcome: lapses)

| **Share of own data in training set** | **Outcome** | **n** | **N below threshold** | **Proportion below threshold** |
| --- | --- | --- | --- | --- |
| None | Lapses | 242 | 43 | 0.178 |
| 10% | Lapses | 198 | 60 | 0.303 |
| 20% | Lapses | 170 | 56 | 0.329 |
| 30% | Lapses | 170 | 66 | 0.388 |

Specificity by share of own data in training set × outcome (proportion below acceptable threshold of 0.5) (outcome: cravings)

| **Share of own data in training set** | **Outcome** | **n** | **N below threshold** | **Proportion below threshold** |
| --- | --- | --- | --- | --- |
| None | Cravings | 360 | 83 | 0.231 |
| 10% | Cravings | 356 | 118 | 0.331 |
| 20% | Cravings | 354 | 111 | 0.314 |
| 30% | Cravings | 354 | 139 | 0.393 |

##### 1.1.7. By outcome

Specificity by outcome

| **Outcome** | **n** | **Median** | **Mean** | **Standard deviation** | **25th percentile** | **75th percentile** | **Minimum** | **Maximum** |
| --- | --- | --- | --- | --- | --- | --- | --- | --- |
| Lapses | 780 | 0.833 | 0.655 | 0.378 | 0.256 | 0.969 | 0.000 | 1.000 |
| Cravings | 1,424 | 0.791 | 0.629 | 0.374 | 0.300 | 0.955 | 0.000 | 1.000 |

##### 1.1.8. By outcome - proportion below threshold

Specificity by outcome (proportion below acceptable threshold of 0.5)

| **Outcome** | **n** | **N below threshold** | **Proportion below threshold** |
| --- | --- | --- | --- |
| Lapses | 780 | 225 | 0.288 |
| Cravings | 1,424 | 451 | 0.317 |

#### 1.2. By specification

##### 1.2.1. By specification

Specificity by specification (prompts per day × use of feature selection × share of own data in training set × outcome)

| **Prompts per day** | **Feature selection** | **Share of own data in training set** | **Outcome** | **n** | **Median** | **Mean** | **Standard deviation** | **25th percentile** | **75th percentile** | **Minimum** | **Maximum** |
| --- | --- | --- | --- | --- | --- | --- | --- | --- | --- | --- | --- |
| 16 | All features | None | Lapses | 25 | 0.975 | 0.874 | 0.234 | 0.859 | 0.987 | 0.109 | 1.000 |
| 16 | All features | None | Cravings | 37 | 0.867 | 0.777 | 0.236 | 0.634 | 0.940 | 0.048 | 1.000 |
| 16 | All features | 10% | Lapses | 20 | 0.932 | 0.785 | 0.322 | 0.799 | 0.987 | 0.000 | 1.000 |
| 16 | All features | 10% | Cravings | 37 | 0.822 | 0.667 | 0.363 | 0.423 | 0.949 | 0.000 | 1.000 |
| 16 | All features | 20% | Lapses | 17 | 0.945 | 0.769 | 0.365 | 0.806 | 0.983 | 0.000 | 1.000 |
| 16 | All features | 20% | Cravings | 36 | 0.739 | 0.652 | 0.363 | 0.439 | 0.973 | 0.000 | 1.000 |
| 16 | All features | 30% | Lapses | 17 | 0.951 | 0.775 | 0.339 | 0.734 | 0.990 | 0.011 | 1.000 |
| 16 | All features | 30% | Cravings | 36 | 0.815 | 0.627 | 0.385 | 0.272 | 0.951 | 0.000 | 1.000 |
| 16 | Selected features | None | Lapses | 25 | 0.967 | 0.801 | 0.309 | 0.817 | 0.988 | 0.000 | 1.000 |
| 16 | Selected features | None | Cravings | 37 | 0.845 | 0.753 | 0.249 | 0.664 | 0.921 | 0.000 | 1.000 |
| 16 | Selected features | 10% | Lapses | 20 | 0.911 | 0.769 | 0.331 | 0.772 | 0.978 | 0.000 | 1.000 |
| 16 | Selected features | 10% | Cravings | 37 | 0.849 | 0.662 | 0.370 | 0.462 | 0.955 | 0.000 | 1.000 |
| 16 | Selected features | 20% | Lapses | 17 | 0.924 | 0.727 | 0.362 | 0.745 | 0.950 | 0.000 | 1.000 |
| 16 | Selected features | 20% | Cravings | 36 | 0.787 | 0.663 | 0.359 | 0.374 | 0.962 | 0.000 | 1.000 |
| 16 | Selected features | 30% | Lapses | 17 | 0.929 | 0.714 | 0.372 | 0.592 | 0.981 | 0.000 | 1.000 |
| 16 | Selected features | 30% | Cravings | 36 | 0.815 | 0.634 | 0.374 | 0.268 | 0.950 | 0.000 | 1.000 |
| 6 | All features | None | Lapses | 24 | 0.859 | 0.720 | 0.315 | 0.620 | 0.950 | 0.000 | 1.000 |
| 6 | All features | None | Cravings | 35 | 0.750 | 0.671 | 0.297 | 0.542 | 0.924 | 0.000 | 1.000 |
| 6 | All features | 10% | Lapses | 20 | 0.833 | 0.668 | 0.351 | 0.432 | 0.891 | 0.000 | 1.000 |
| 6 | All features | 10% | Cravings | 35 | 0.791 | 0.639 | 0.341 | 0.335 | 0.901 | 0.000 | 1.000 |
| 6 | All features | 20% | Lapses | 17 | 0.750 | 0.599 | 0.404 | 0.118 | 0.955 | 0.000 | 1.000 |
| 6 | All features | 20% | Cravings | 35 | 0.882 | 0.676 | 0.371 | 0.450 | 0.976 | 0.000 | 1.000 |
| 6 | All features | 30% | Lapses | 17 | 0.881 | 0.599 | 0.433 | 0.190 | 0.972 | 0.000 | 1.000 |
| 6 | All features | 30% | Cravings | 35 | 0.706 | 0.568 | 0.416 | 0.097 | 0.960 | 0.000 | 1.000 |
| 6 | Selected features | None | Lapses | 24 | 0.864 | 0.707 | 0.353 | 0.612 | 0.963 | 0.000 | 1.000 |
| 6 | Selected features | None | Cravings | 35 | 0.795 | 0.703 | 0.285 | 0.635 | 0.907 | 0.000 | 1.000 |
| 6 | Selected features | 10% | Lapses | 20 | 0.840 | 0.602 | 0.413 | 0.104 | 0.948 | 0.000 | 1.000 |
| 6 | Selected features | 10% | Cravings | 35 | 0.829 | 0.630 | 0.360 | 0.354 | 0.908 | 0.000 | 1.000 |
| 6 | Selected features | 20% | Lapses | 17 | 0.829 | 0.611 | 0.393 | 0.219 | 0.977 | 0.000 | 1.000 |
| 6 | Selected features | 20% | Cravings | 35 | 0.868 | 0.666 | 0.381 | 0.333 | 0.956 | 0.000 | 1.000 |
| 6 | Selected features | 30% | Lapses | 17 | 0.857 | 0.551 | 0.468 | 0.000 | 0.952 | 0.000 | 1.000 |
| 6 | Selected features | 30% | Cravings | 35 | 0.714 | 0.561 | 0.412 | 0.032 | 0.926 | 0.000 | 1.000 |
| 5 | All features | None | Lapses | 24 | 0.869 | 0.767 | 0.268 | 0.670 | 0.985 | 0.071 | 1.000 |
| 5 | All features | None | Cravings | 35 | 0.750 | 0.678 | 0.294 | 0.478 | 0.938 | 0.000 | 1.000 |
| 5 | All features | 10% | Lapses | 20 | 0.776 | 0.666 | 0.331 | 0.440 | 0.952 | 0.000 | 1.000 |
| 5 | All features | 10% | Cravings | 34 | 0.766 | 0.631 | 0.362 | 0.250 | 0.938 | 0.000 | 1.000 |
| 5 | All features | 20% | Lapses | 17 | 0.778 | 0.636 | 0.370 | 0.542 | 0.970 | 0.000 | 1.000 |
| 5 | All features | 20% | Cravings | 34 | 0.883 | 0.669 | 0.359 | 0.357 | 0.966 | 0.000 | 1.000 |
| 5 | All features | 30% | Lapses | 17 | 0.679 | 0.509 | 0.421 | 0.067 | 0.931 | 0.000 | 1.000 |
| 5 | All features | 30% | Cravings | 34 | 0.736 | 0.589 | 0.432 | 0.017 | 1.000 | 0.000 | 1.000 |
| 5 | Selected features | None | Lapses | 24 | 0.869 | 0.728 | 0.326 | 0.618 | 0.961 | 0.000 | 1.000 |
| 5 | Selected features | None | Cravings | 35 | 0.765 | 0.676 | 0.316 | 0.509 | 0.951 | 0.000 | 1.000 |
| 5 | Selected features | 10% | Lapses | 20 | 0.846 | 0.676 | 0.373 | 0.474 | 0.951 | 0.000 | 1.000 |
| 5 | Selected features | 10% | Cravings | 34 | 0.845 | 0.611 | 0.417 | 0.132 | 1.000 | 0.000 | 1.000 |
| 5 | Selected features | 20% | Lapses | 17 | 0.778 | 0.529 | 0.457 | 0.000 | 0.972 | 0.000 | 1.000 |
| 5 | Selected features | 20% | Cravings | 34 | 0.821 | 0.639 | 0.402 | 0.233 | 1.000 | 0.000 | 1.000 |
| 5 | Selected features | 30% | Lapses | 17 | 0.750 | 0.503 | 0.471 | 0.000 | 0.960 | 0.000 | 1.000 |
| 5 | Selected features | 30% | Cravings | 34 | 0.766 | 0.577 | 0.439 | 0.012 | 1.000 | 0.000 | 1.000 |
| 4 | All features | None | Lapses | 24 | 0.803 | 0.720 | 0.289 | 0.594 | 0.972 | 0.000 | 1.000 |
| 4 | All features | None | Cravings | 37 | 0.778 | 0.656 | 0.334 | 0.531 | 0.921 | 0.000 | 1.000 |
| 4 | All features | 10% | Lapses | 20 | 0.783 | 0.634 | 0.389 | 0.278 | 0.964 | 0.000 | 1.000 |
| 4 | All features | 10% | Cravings | 36 | 0.743 | 0.636 | 0.352 | 0.475 | 0.915 | 0.000 | 1.000 |
| 4 | All features | 20% | Lapses | 17 | 0.731 | 0.556 | 0.421 | 0.000 | 0.935 | 0.000 | 1.000 |
| 4 | All features | 20% | Cravings | 36 | 0.837 | 0.648 | 0.376 | 0.298 | 0.962 | 0.000 | 1.000 |
| 4 | All features | 30% | Lapses | 17 | 0.667 | 0.524 | 0.436 | 0.111 | 1.000 | 0.000 | 1.000 |
| 4 | All features | 30% | Cravings | 36 | 0.764 | 0.595 | 0.399 | 0.205 | 0.933 | 0.000 | 1.000 |
| 4 | Selected features | None | Lapses | 24 | 0.806 | 0.671 | 0.365 | 0.578 | 0.955 | 0.000 | 1.000 |
| 4 | Selected features | None | Cravings | 37 | 0.774 | 0.648 | 0.335 | 0.375 | 0.966 | 0.000 | 1.000 |
| 4 | Selected features | 10% | Lapses | 20 | 0.749 | 0.548 | 0.432 | 0.087 | 0.963 | 0.000 | 1.000 |
| 4 | Selected features | 10% | Cravings | 36 | 0.750 | 0.609 | 0.391 | 0.124 | 0.942 | 0.000 | 1.000 |
| 4 | Selected features | 20% | Lapses | 17 | 0.800 | 0.500 | 0.463 | 0.000 | 0.955 | 0.000 | 1.000 |
| 4 | Selected features | 20% | Cravings | 36 | 0.837 | 0.628 | 0.405 | 0.205 | 0.959 | 0.000 | 1.000 |
| 4 | Selected features | 30% | Lapses | 17 | 0.889 | 0.527 | 0.491 | 0.000 | 1.000 | 0.000 | 1.000 |
| 4 | Selected features | 30% | Cravings | 36 | 0.739 | 0.562 | 0.441 | 0.000 | 0.972 | 0.000 | 1.000 |
| 3 | All features | None | Lapses | 24 | 0.804 | 0.718 | 0.267 | 0.625 | 0.902 | 0.000 | 1.000 |
| 3 | All features | None | Cravings | 36 | 0.765 | 0.633 | 0.340 | 0.411 | 0.921 | 0.000 | 1.000 |
| 3 | All features | 10% | Lapses | 19 | 0.654 | 0.577 | 0.352 | 0.341 | 0.850 | 0.000 | 1.000 |
| 3 | All features | 10% | Cravings | 36 | 0.739 | 0.606 | 0.366 | 0.333 | 0.934 | 0.000 | 1.000 |
| 3 | All features | 20% | Lapses | 17 | 0.737 | 0.596 | 0.357 | 0.200 | 0.864 | 0.000 | 1.000 |
| 3 | All features | 20% | Cravings | 36 | 0.783 | 0.609 | 0.419 | 0.140 | 1.000 | 0.000 | 1.000 |
| 3 | All features | 30% | Lapses | 17 | 0.750 | 0.591 | 0.416 | 0.143 | 1.000 | 0.000 | 1.000 |
| 3 | All features | 30% | Cravings | 36 | 0.587 | 0.515 | 0.429 | 0.000 | 0.949 | 0.000 | 1.000 |
| 3 | Selected features | None | Lapses | 24 | 0.793 | 0.704 | 0.335 | 0.619 | 0.963 | 0.000 | 1.000 |
| 3 | Selected features | None | Cravings | 36 | 0.804 | 0.674 | 0.340 | 0.475 | 0.935 | 0.000 | 1.000 |
| 3 | Selected features | 10% | Lapses | 19 | 0.737 | 0.557 | 0.402 | 0.095 | 0.884 | 0.000 | 1.000 |
| 3 | Selected features | 10% | Cravings | 36 | 0.376 | 0.461 | 0.438 | 0.000 | 0.967 | 0.000 | 1.000 |
| 3 | Selected features | 20% | Lapses | 17 | 0.765 | 0.572 | 0.396 | 0.167 | 0.952 | 0.000 | 1.000 |
| 3 | Selected features | 20% | Cravings | 36 | 0.806 | 0.585 | 0.423 | 0.113 | 1.000 | 0.000 | 1.000 |
| 3 | Selected features | 30% | Lapses | 17 | 0.714 | 0.534 | 0.461 | 0.000 | 1.000 | 0.000 | 1.000 |
| 3 | Selected features | 30% | Cravings | 36 | 0.497 | 0.468 | 0.453 | 0.000 | 0.956 | 0.000 | 1.000 |

##### 1.2.2. By specification - proportion below threshold

Specificity by specification (prompts per day × use of feature selection × share of own data in training set × outcome) - proportion below acceptable threshold of 0.5

| **Prompts per day** | **Feature selection** | **Share of own data in training set** | **Outcome** | **n** | **N below threshold** | **Proportion below threshold** |
| --- | --- | --- | --- | --- | --- | --- |
| 16 | All features | None | Lapses | 25 | 2 | 0.080 |
| 16 | All features | None | Cravings | 37 | 6 | 0.162 |
| 16 | All features | 10% | Lapses | 20 | 3 | 0.150 |
| 16 | All features | 10% | Cravings | 37 | 11 | 0.297 |
| 16 | All features | 20% | Lapses | 17 | 3 | 0.176 |
| 16 | All features | 20% | Cravings | 36 | 10 | 0.278 |
| 16 | All features | 30% | Lapses | 17 | 3 | 0.176 |
| 16 | All features | 30% | Cravings | 36 | 12 | 0.333 |
| 16 | Selected features | None | Lapses | 25 | 4 | 0.160 |
| 16 | Selected features | None | Cravings | 37 | 6 | 0.162 |
| 16 | Selected features | 10% | Lapses | 20 | 3 | 0.150 |
| 16 | Selected features | 10% | Cravings | 37 | 10 | 0.270 |
| 16 | Selected features | 20% | Lapses | 17 | 4 | 0.235 |
| 16 | Selected features | 20% | Cravings | 36 | 10 | 0.278 |
| 16 | Selected features | 30% | Lapses | 17 | 4 | 0.235 |
| 16 | Selected features | 30% | Cravings | 36 | 11 | 0.306 |
| 6 | All features | None | Lapses | 24 | 6 | 0.250 |
| 6 | All features | None | Cravings | 35 | 8 | 0.229 |
| 6 | All features | 10% | Lapses | 20 | 6 | 0.300 |
| 6 | All features | 10% | Cravings | 35 | 10 | 0.286 |
| 6 | All features | 20% | Lapses | 17 | 6 | 0.353 |
| 6 | All features | 20% | Cravings | 35 | 9 | 0.257 |
| 6 | All features | 30% | Lapses | 17 | 7 | 0.412 |
| 6 | All features | 30% | Cravings | 35 | 15 | 0.429 |
| 6 | Selected features | None | Lapses | 24 | 5 | 0.208 |
| 6 | Selected features | None | Cravings | 35 | 6 | 0.171 |
| 6 | Selected features | 10% | Lapses | 20 | 7 | 0.350 |
| 6 | Selected features | 10% | Cravings | 35 | 11 | 0.314 |
| 6 | Selected features | 20% | Lapses | 17 | 6 | 0.353 |
| 6 | Selected features | 20% | Cravings | 35 | 9 | 0.257 |
| 6 | Selected features | 30% | Lapses | 17 | 7 | 0.412 |
| 6 | Selected features | 30% | Cravings | 35 | 13 | 0.371 |
| 5 | All features | None | Lapses | 24 | 4 | 0.167 |
| 5 | All features | None | Cravings | 35 | 10 | 0.286 |
| 5 | All features | 10% | Lapses | 20 | 7 | 0.350 |
| 5 | All features | 10% | Cravings | 34 | 10 | 0.294 |
| 5 | All features | 20% | Lapses | 17 | 4 | 0.235 |
| 5 | All features | 20% | Cravings | 34 | 10 | 0.294 |
| 5 | All features | 30% | Lapses | 17 | 8 | 0.471 |
| 5 | All features | 30% | Cravings | 34 | 13 | 0.382 |
| 5 | Selected features | None | Lapses | 24 | 4 | 0.167 |
| 5 | Selected features | None | Cravings | 35 | 8 | 0.229 |
| 5 | Selected features | 10% | Lapses | 20 | 6 | 0.300 |
| 5 | Selected features | 10% | Cravings | 34 | 12 | 0.353 |
| 5 | Selected features | 20% | Lapses | 17 | 7 | 0.412 |
| 5 | Selected features | 20% | Cravings | 34 | 12 | 0.353 |
| 5 | Selected features | 30% | Lapses | 17 | 8 | 0.471 |
| 5 | Selected features | 30% | Cravings | 34 | 13 | 0.382 |
| 4 | All features | None | Lapses | 24 | 3 | 0.125 |
| 4 | All features | None | Cravings | 37 | 9 | 0.243 |
| 4 | All features | 10% | Lapses | 20 | 7 | 0.350 |
| 4 | All features | 10% | Cravings | 36 | 10 | 0.278 |
| 4 | All features | 20% | Lapses | 17 | 7 | 0.412 |
| 4 | All features | 20% | Cravings | 36 | 10 | 0.278 |
| 4 | All features | 30% | Lapses | 17 | 8 | 0.471 |
| 4 | All features | 30% | Cravings | 36 | 14 | 0.389 |
| 4 | Selected features | None | Lapses | 24 | 6 | 0.250 |
| 4 | Selected features | None | Cravings | 37 | 11 | 0.297 |
| 4 | Selected features | 10% | Lapses | 20 | 9 | 0.450 |
| 4 | Selected features | 10% | Cravings | 36 | 11 | 0.306 |
| 4 | Selected features | 20% | Lapses | 17 | 8 | 0.471 |
| 4 | Selected features | 20% | Cravings | 36 | 11 | 0.306 |
| 4 | Selected features | 30% | Lapses | 17 | 8 | 0.471 |
| 4 | Selected features | 30% | Cravings | 36 | 14 | 0.389 |
| 3 | All features | None | Lapses | 24 | 4 | 0.167 |
| 3 | All features | None | Cravings | 36 | 10 | 0.278 |
| 3 | All features | 10% | Lapses | 19 | 5 | 0.263 |
| 3 | All features | 10% | Cravings | 36 | 14 | 0.389 |
| 3 | All features | 20% | Lapses | 17 | 5 | 0.294 |
| 3 | All features | 20% | Cravings | 36 | 14 | 0.389 |
| 3 | All features | 30% | Lapses | 17 | 6 | 0.353 |
| 3 | All features | 30% | Cravings | 36 | 16 | 0.444 |
| 3 | Selected features | None | Lapses | 24 | 5 | 0.208 |
| 3 | Selected features | None | Cravings | 36 | 9 | 0.250 |
| 3 | Selected features | 10% | Lapses | 19 | 7 | 0.368 |
| 3 | Selected features | 10% | Cravings | 36 | 19 | 0.528 |
| 3 | Selected features | 20% | Lapses | 17 | 6 | 0.353 |
| 3 | Selected features | 20% | Cravings | 36 | 16 | 0.444 |
| 3 | Selected features | 30% | Lapses | 17 | 7 | 0.412 |
| 3 | Selected features | 30% | Cravings | 36 | 18 | 0.500 |

#### 1.3. By participant

##### 1.3.1. By participant (overall)

Specificity by participant

| **Participant** | **n** | **Median** | **Mean** | **Standard deviation** | **25th percentile** | **75th percentile** | **Minimum** | **Maximum** |
| --- | --- | --- | --- | --- | --- | --- | --- | --- |
| p1 | 80 | 0.723 | 0.634 | 0.321 | 0.471 | 0.878 | 0.000 | 1.000 |
| p10 | 40 | 1.000 | 0.974 | 0.080 | 1.000 | 1.000 | 0.609 | 1.000 |
| p101 | 80 | 0.800 | 0.733 | 0.251 | 0.645 | 0.923 | 0.000 | 1.000 |
| p107 | 80 | 0.875 | 0.836 | 0.185 | 0.764 | 0.993 | 0.161 | 1.000 |
| p12 | 80 | 0.765 | 0.598 | 0.402 | 0.182 | 0.971 | 0.000 | 1.000 |
| p128 | 80 | 0.404 | 0.422 | 0.354 | 0.090 | 0.752 | 0.000 | 1.000 |
| p129 | 40 | 1.000 | 0.974 | 0.041 | 0.961 | 1.000 | 0.811 | 1.000 |
| p131 | 42 | 0.040 | 0.121 | 0.224 | 0.000 | 0.122 | 0.000 | 0.994 |
| p132 | 60 | 0.574 | 0.502 | 0.358 | 0.000 | 0.783 | 0.000 | 0.994 |
| p135 | 80 | 0.651 | 0.595 | 0.293 | 0.372 | 0.843 | 0.000 | 1.000 |
| p141 | 80 | 0.753 | 0.549 | 0.389 | 0.000 | 0.869 | 0.000 | 1.000 |
| p144 | 80 | 0.913 | 0.903 | 0.069 | 0.864 | 0.955 | 0.636 | 1.000 |
| p147 | 40 | 0.526 | 0.532 | 0.231 | 0.361 | 0.720 | 0.048 | 0.920 |
| p15 | 80 | 0.877 | 0.612 | 0.415 | 0.113 | 0.989 | 0.000 | 1.000 |
| p20 | 40 | 0.000 | 0.012 | 0.079 | 0.000 | 0.000 | 0.000 | 0.500 |
| p21 | 80 | 0.054 | 0.286 | 0.351 | 0.000 | 0.670 | 0.000 | 1.000 |
| p22 | 80 | 0.843 | 0.731 | 0.312 | 0.606 | 0.973 | 0.000 | 1.000 |
| p27 | 32 | 0.945 | 0.935 | 0.041 | 0.900 | 0.969 | 0.846 | 0.991 |
| p30 | 50 | 0.388 | 0.452 | 0.399 | 0.006 | 0.852 | 0.000 | 1.000 |
| p39 | 40 | 0.650 | 0.597 | 0.262 | 0.510 | 0.720 | 0.000 | 1.000 |
| p44 | 40 | 0.907 | 0.896 | 0.076 | 0.870 | 0.936 | 0.692 | 1.000 |
| p59 | 40 | 0.930 | 0.906 | 0.072 | 0.894 | 0.944 | 0.696 | 1.000 |
| p6 | 80 | 0.000 | 0.185 | 0.269 | 0.000 | 0.358 | 0.000 | 0.867 |
| p61 | 80 | 1.000 | 0.979 | 0.045 | 0.976 | 1.000 | 0.744 | 1.000 |
| p62 | 60 | 0.922 | 0.860 | 0.194 | 0.806 | 1.000 | 0.083 | 1.000 |
| p63 | 80 | 0.941 | 0.866 | 0.197 | 0.841 | 0.974 | 0.067 | 1.000 |
| p67 | 42 | 1.000 | 0.991 | 0.015 | 0.984 | 1.000 | 0.933 | 1.000 |
| p7 | 80 | 0.934 | 0.851 | 0.212 | 0.805 | 0.978 | 0.050 | 1.000 |
| p70 | 6 | 0.987 | 0.985 | 0.009 | 0.978 | 0.992 | 0.974 | 0.993 |
| p74 | 50 | 0.818 | 0.726 | 0.301 | 0.600 | 1.000 | 0.000 | 1.000 |
| p78 | 40 | 0.000 | 0.199 | 0.318 | 0.000 | 0.271 | 0.000 | 1.000 |
| p79 | 40 | 0.948 | 0.882 | 0.218 | 0.894 | 1.000 | 0.000 | 1.000 |
| p87 | 58 | 0.000 | 0.286 | 0.392 | 0.000 | 0.634 | 0.000 | 1.000 |
| p9 | 50 | 0.832 | 0.764 | 0.200 | 0.671 | 0.878 | 0.000 | 1.000 |
| p93 | 40 | 0.200 | 0.226 | 0.177 | 0.000 | 0.383 | 0.000 | 0.538 |
| p95 | 80 | 0.341 | 0.408 | 0.343 | 0.067 | 0.781 | 0.000 | 1.000 |
| p97 | 74 | 0.967 | 0.886 | 0.208 | 0.891 | 0.984 | 0.000 | 1.000 |

##### 1.3.2. By participant (overall) - proportion below threshold

Specificity by participant (proportion below acceptable threshold of 0.5)

| **Participant** | **n** | **N below threshold** | **Proportion below threshold** |
| --- | --- | --- | --- |
| p1 | 80 | 21 | 0.263 |
| p10 | 40 | 0 | 0.000 |
| p101 | 80 | 10 | 0.125 |
| p107 | 80 | 4 | 0.050 |
| p12 | 80 | 32 | 0.400 |
| p128 | 80 | 44 | 0.550 |
| p129 | 40 | 0 | 0.000 |
| p131 | 42 | 40 | 0.952 |
| p132 | 60 | 20 | 0.333 |
| p135 | 80 | 26 | 0.325 |
| p141 | 80 | 30 | 0.375 |
| p144 | 80 | 0 | 0.000 |
| p147 | 40 | 18 | 0.450 |
| p15 | 80 | 32 | 0.400 |
| p20 | 40 | 39 | 0.975 |
| p21 | 80 | 53 | 0.662 |
| p22 | 80 | 18 | 0.225 |
| p27 | 32 | 0 | 0.000 |
| p30 | 50 | 28 | 0.560 |
| p39 | 40 | 10 | 0.250 |
| p44 | 40 | 0 | 0.000 |
| p59 | 40 | 0 | 0.000 |
| p6 | 80 | 63 | 0.787 |
| p61 | 80 | 0 | 0.000 |
| p62 | 60 | 5 | 0.083 |
| p63 | 80 | 5 | 0.062 |
| p67 | 42 | 0 | 0.000 |
| p7 | 80 | 6 | 0.075 |
| p70 | 6 | 0 | 0.000 |
| p74 | 50 | 9 | 0.180 |
| p78 | 40 | 34 | 0.850 |
| p79 | 40 | 3 | 0.075 |
| p87 | 58 | 38 | 0.655 |
| p9 | 50 | 3 | 0.060 |
| p93 | 40 | 38 | 0.950 |
| p95 | 80 | 43 | 0.537 |
| p97 | 74 | 4 | 0.054 |

Specificity by participant × outcome (outcome: lapses)

| **Participant** | **Outcome** | **n** | **Median** | **Mean** | **Standard deviation** | **25th percentile** | **75th percentile** | **Minimum** | **Maximum** |
| --- | --- | --- | --- | --- | --- | --- | --- | --- | --- |
| p1 | Lapses | 40 | 0.625 | 0.523 | 0.331 | 0.123 | 0.778 | 0.000 | 0.948 |
| p101 | Lapses | 40 | 0.885 | 0.878 | 0.089 | 0.809 | 0.948 | 0.727 | 1.000 |
| p107 | Lapses | 40 | 0.987 | 0.951 | 0.085 | 0.945 | 1.000 | 0.571 | 1.000 |
| p12 | Lapses | 40 | 0.182 | 0.238 | 0.240 | 0.048 | 0.338 | 0.000 | 0.802 |
| p128 | Lapses | 40 | 0.141 | 0.301 | 0.307 | 0.084 | 0.644 | 0.000 | 0.897 |
| p131 | Lapses | 2 | 0.984 | 0.984 | 0.013 | 0.980 | 0.989 | 0.975 | 0.994 |
| p132 | Lapses | 20 | 0.852 | 0.829 | 0.144 | 0.696 | 0.951 | 0.593 | 0.994 |
| p135 | Lapses | 40 | 0.821 | 0.731 | 0.272 | 0.651 | 0.919 | 0.000 | 1.000 |
| p141 | Lapses | 40 | 0.847 | 0.844 | 0.084 | 0.790 | 0.926 | 0.625 | 0.964 |
| p144 | Lapses | 40 | 0.931 | 0.915 | 0.062 | 0.864 | 0.962 | 0.769 | 1.000 |
| p15 | Lapses | 40 | 0.109 | 0.254 | 0.294 | 0.040 | 0.350 | 0.000 | 0.917 |
| p21 | Lapses | 40 | 0.000 | 0.011 | 0.032 | 0.000 | 0.000 | 0.000 | 0.143 |
| p22 | Lapses | 40 | 0.969 | 0.941 | 0.065 | 0.927 | 0.978 | 0.780 | 1.000 |
| p30 | Lapses | 10 | 0.980 | 0.982 | 0.014 | 0.974 | 0.996 | 0.966 | 1.000 |
| p6 | Lapses | 40 | 0.000 | 0.034 | 0.097 | 0.000 | 0.000 | 0.000 | 0.500 |
| p61 | Lapses | 40 | 1.000 | 0.989 | 0.014 | 0.979 | 1.000 | 0.944 | 1.000 |
| p62 | Lapses | 20 | 1.000 | 1.000 | 0.000 | 1.000 | 1.000 | 1.000 | 1.000 |
| p63 | Lapses | 40 | 0.955 | 0.884 | 0.205 | 0.920 | 0.978 | 0.067 | 1.000 |
| p67 | Lapses | 10 | 0.989 | 0.987 | 0.012 | 0.977 | 0.998 | 0.967 | 1.000 |
| p7 | Lapses | 40 | 0.868 | 0.767 | 0.256 | 0.727 | 0.951 | 0.050 | 1.000 |
| p74 | Lapses | 10 | 0.997 | 0.971 | 0.047 | 0.970 | 1.000 | 0.872 | 1.000 |
| p87 | Lapses | 18 | 0.837 | 0.783 | 0.220 | 0.619 | 0.981 | 0.276 | 1.000 |
| p9 | Lapses | 10 | 0.662 | 0.726 | 0.144 | 0.627 | 0.758 | 0.600 | 0.988 |
| p95 | Lapses | 40 | 0.520 | 0.462 | 0.333 | 0.101 | 0.753 | 0.000 | 1.000 |
| p97 | Lapses | 40 | 0.932 | 0.809 | 0.258 | 0.751 | 0.970 | 0.000 | 1.000 |

Specificity by participant × outcome (outcome: cravings)

| **Participant** | **Outcome** | **n** | **Median** | **Mean** | **Standard deviation** | **25th percentile** | **75th percentile** | **Minimum** | **Maximum** |
| --- | --- | --- | --- | --- | --- | --- | --- | --- | --- |
| p1 | Cravings | 40 | 0.817 | 0.744 | 0.273 | 0.695 | 0.947 | 0.000 | 1.000 |
| p10 | Cravings | 40 | 1.000 | 0.974 | 0.080 | 1.000 | 1.000 | 0.609 | 1.000 |
| p101 | Cravings | 40 | 0.642 | 0.589 | 0.277 | 0.498 | 0.774 | 0.000 | 1.000 |
| p107 | Cravings | 40 | 0.775 | 0.721 | 0.188 | 0.601 | 0.851 | 0.161 | 1.000 |
| p12 | Cravings | 40 | 0.971 | 0.958 | 0.059 | 0.938 | 1.000 | 0.714 | 1.000 |
| p128 | Cravings | 40 | 0.642 | 0.543 | 0.359 | 0.162 | 0.845 | 0.000 | 1.000 |
| p129 | Cravings | 40 | 1.000 | 0.974 | 0.041 | 0.961 | 1.000 | 0.811 | 1.000 |
| p131 | Cravings | 40 | 0.027 | 0.078 | 0.112 | 0.000 | 0.111 | 0.000 | 0.382 |
| p132 | Cravings | 40 | 0.417 | 0.339 | 0.319 | 0.000 | 0.573 | 0.000 | 0.917 |
| p135 | Cravings | 40 | 0.455 | 0.460 | 0.249 | 0.239 | 0.650 | 0.000 | 0.917 |
| p141 | Cravings | 40 | 0.000 | 0.255 | 0.349 | 0.000 | 0.425 | 0.000 | 1.000 |
| p144 | Cravings | 40 | 0.897 | 0.890 | 0.073 | 0.864 | 0.948 | 0.636 | 0.974 |
| p147 | Cravings | 40 | 0.526 | 0.532 | 0.231 | 0.361 | 0.720 | 0.048 | 0.920 |
| p15 | Cravings | 40 | 0.992 | 0.969 | 0.039 | 0.938 | 1.000 | 0.868 | 1.000 |
| p20 | Cravings | 40 | 0.000 | 0.012 | 0.079 | 0.000 | 0.000 | 0.000 | 0.500 |
| p21 | Cravings | 40 | 0.673 | 0.560 | 0.308 | 0.348 | 0.774 | 0.000 | 1.000 |
| p22 | Cravings | 40 | 0.601 | 0.521 | 0.320 | 0.258 | 0.776 | 0.000 | 1.000 |
| p27 | Cravings | 32 | 0.945 | 0.935 | 0.041 | 0.900 | 0.969 | 0.846 | 0.991 |
| p30 | Cravings | 40 | 0.229 | 0.320 | 0.331 | 0.000 | 0.552 | 0.000 | 0.955 |
| p39 | Cravings | 40 | 0.650 | 0.597 | 0.262 | 0.510 | 0.720 | 0.000 | 1.000 |
| p44 | Cravings | 40 | 0.907 | 0.896 | 0.076 | 0.870 | 0.936 | 0.692 | 1.000 |
| p59 | Cravings | 40 | 0.930 | 0.906 | 0.072 | 0.894 | 0.944 | 0.696 | 1.000 |
| p6 | Cravings | 40 | 0.284 | 0.337 | 0.301 | 0.054 | 0.603 | 0.000 | 0.867 |
| p61 | Cravings | 40 | 0.992 | 0.970 | 0.061 | 0.972 | 1.000 | 0.744 | 1.000 |
| p62 | Cravings | 40 | 0.862 | 0.789 | 0.205 | 0.748 | 0.922 | 0.083 | 1.000 |
| p63 | Cravings | 40 | 0.912 | 0.847 | 0.191 | 0.820 | 0.966 | 0.158 | 1.000 |
| p67 | Cravings | 32 | 1.000 | 0.992 | 0.016 | 0.993 | 1.000 | 0.933 | 1.000 |
| p7 | Cravings | 40 | 0.972 | 0.935 | 0.104 | 0.921 | 1.000 | 0.538 | 1.000 |
| p70 | Cravings | 6 | 0.987 | 0.985 | 0.009 | 0.978 | 0.992 | 0.974 | 0.993 |
| p74 | Cravings | 40 | 0.726 | 0.665 | 0.306 | 0.500 | 0.887 | 0.000 | 1.000 |
| p78 | Cravings | 40 | 0.000 | 0.199 | 0.318 | 0.000 | 0.271 | 0.000 | 1.000 |
| p79 | Cravings | 40 | 0.948 | 0.882 | 0.218 | 0.894 | 1.000 | 0.000 | 1.000 |
| p87 | Cravings | 40 | 0.000 | 0.062 | 0.194 | 0.000 | 0.000 | 0.000 | 0.750 |
| p9 | Cravings | 40 | 0.839 | 0.773 | 0.212 | 0.736 | 0.883 | 0.000 | 1.000 |
| p93 | Cravings | 40 | 0.200 | 0.226 | 0.177 | 0.000 | 0.383 | 0.000 | 0.538 |
| p95 | Cravings | 40 | 0.233 | 0.353 | 0.348 | 0.047 | 0.803 | 0.000 | 0.889 |
| p97 | Cravings | 34 | 0.983 | 0.977 | 0.031 | 0.968 | 1.000 | 0.867 | 1.000 |

Specificity by participant × outcome (proportion below acceptable threshold of 0.5) (outcome: lapses)

| **Participant** | **Outcome** | **n** | **N below threshold** | **Proportion below threshold** |
| --- | --- | --- | --- | --- |
| p1 | Lapses | 40 | 16 | 0.400 |
| p101 | Lapses | 40 | 0 | 0.000 |
| p107 | Lapses | 40 | 0 | 0.000 |
| p12 | Lapses | 40 | 32 | 0.800 |
| p128 | Lapses | 40 | 29 | 0.725 |
| p131 | Lapses | 2 | 0 | 0.000 |
| p132 | Lapses | 20 | 0 | 0.000 |
| p135 | Lapses | 40 | 5 | 0.125 |
| p141 | Lapses | 40 | 0 | 0.000 |
| p144 | Lapses | 40 | 0 | 0.000 |
| p15 | Lapses | 40 | 32 | 0.800 |
| p21 | Lapses | 40 | 40 | 1.000 |
| p22 | Lapses | 40 | 0 | 0.000 |
| p30 | Lapses | 10 | 0 | 0.000 |
| p6 | Lapses | 40 | 39 | 0.975 |
| p61 | Lapses | 40 | 0 | 0.000 |
| p62 | Lapses | 20 | 0 | 0.000 |
| p63 | Lapses | 40 | 3 | 0.075 |
| p67 | Lapses | 10 | 0 | 0.000 |
| p7 | Lapses | 40 | 6 | 0.150 |
| p74 | Lapses | 10 | 0 | 0.000 |
| p87 | Lapses | 18 | 2 | 0.111 |
| p9 | Lapses | 10 | 0 | 0.000 |
| p95 | Lapses | 40 | 17 | 0.425 |
| p97 | Lapses | 40 | 4 | 0.100 |

Specificity by participant × outcome (proportion below acceptable threshold of 0.5) (outcome: cravings)

| **Participant** | **Outcome** | **n** | **N below threshold** | **Proportion below threshold** |
| --- | --- | --- | --- | --- |
| p1 | Cravings | 40 | 5 | 0.125 |
| p10 | Cravings | 40 | 0 | 0.000 |
| p101 | Cravings | 40 | 10 | 0.250 |
| p107 | Cravings | 40 | 4 | 0.100 |
| p12 | Cravings | 40 | 0 | 0.000 |
| p128 | Cravings | 40 | 15 | 0.375 |
| p129 | Cravings | 40 | 0 | 0.000 |
| p131 | Cravings | 40 | 40 | 1.000 |
| p132 | Cravings | 40 | 20 | 0.500 |
| p135 | Cravings | 40 | 21 | 0.525 |
| p141 | Cravings | 40 | 30 | 0.750 |
| p144 | Cravings | 40 | 0 | 0.000 |
| p147 | Cravings | 40 | 18 | 0.450 |
| p15 | Cravings | 40 | 0 | 0.000 |
| p20 | Cravings | 40 | 39 | 0.975 |
| p21 | Cravings | 40 | 13 | 0.325 |
| p22 | Cravings | 40 | 18 | 0.450 |
| p27 | Cravings | 32 | 0 | 0.000 |
| p30 | Cravings | 40 | 28 | 0.700 |
| p39 | Cravings | 40 | 10 | 0.250 |
| p44 | Cravings | 40 | 0 | 0.000 |
| p59 | Cravings | 40 | 0 | 0.000 |
| p6 | Cravings | 40 | 24 | 0.600 |
| p61 | Cravings | 40 | 0 | 0.000 |
| p62 | Cravings | 40 | 5 | 0.125 |
| p63 | Cravings | 40 | 2 | 0.050 |
| p67 | Cravings | 32 | 0 | 0.000 |
| p7 | Cravings | 40 | 0 | 0.000 |
| p70 | Cravings | 6 | 0 | 0.000 |
| p74 | Cravings | 40 | 9 | 0.225 |
| p78 | Cravings | 40 | 34 | 0.850 |
| p79 | Cravings | 40 | 3 | 0.075 |
| p87 | Cravings | 40 | 36 | 0.900 |
| p9 | Cravings | 40 | 3 | 0.075 |
| p93 | Cravings | 40 | 38 | 0.950 |
| p95 | Cravings | 40 | 26 | 0.650 |
| p97 | Cravings | 34 | 0 | 0.000 |
