## Supplementary material for "Optimising supervised machine learning algorithms predicting cigarette cravings and lapses for a smoking cessation just-in-time adaptive intervention (JITAI)": S8 Appendix F1-Score models and marginal effects

### 1. Comprehensive analysis: F1-score

#### 1.1. Overall models

##### 1.1.1. Main model results

F1-score (Overall) - zero-inflated beta regression (with random effects)

| **Predictor** | **Estimate (logit scale)** | **Standard error** | **95% confidence interval (logit)** | **z-statistic** | **p-value** | **Sig.** |
| --- | --- | --- | --- | --- | --- | --- |
| Intercept | -0.009 | 0.179 | [-0.359, 0.341] | -0.050 | 0.960 |  |
| Prompts per day: 6 | 0.077 | 0.059 | [-0.038, 0.193] | 1.310 | 0.190 |  |
| Prompts per day: 5 | -0.065 | 0.060 | [-0.183, 0.053] | -1.077 | 0.281 |  |
| Prompts per day: 4 | 0.031 | 0.060 | [-0.086, 0.149] | 0.524 | 0.600 |  |
| Prompts per day: 3 | 0.152 | 0.061 | [0.032, 0.272] | 2.489 | 0.013 | * |
| Feature selection: Selected features | 0.065 | 0.038 | [-0.011, 0.140] | 1.679 | 0.093 |  |
| Share of own data: 10% | 0.070 | 0.054 | [-0.035, 0.176] | 1.306 | 0.191 |  |
| Share of own data: 20% | 0.127 | 0.054 | [0.020, 0.233] | 2.328 | 0.020 | * |
| Share of own data: 30% | 0.267 | 0.055 | [0.160, 0.374] | 4.885 | <0.001 | *** |
| Outcome: Cravings | -0.093 | 0.050 | [-0.191, 0.004] | -1.872 | 0.061 |  |
| Reference levels: Prompts per day = 16; Feature selection = All features; Share of own data = None; Outcome = Lapses | | | | | | |
| Note: Estimate coefficients are on the logit scale (conditional component only). | | | | | | |
| This is a zero-inflated beta model with separate processes for zeros and values between 0 and 1. | | | | | | |
| For interpretable effect sizes (actual differences in predicted values), see pairwise comparisons on the response scale (0-1). | | | | | | |
| Significance levels: *** p<0.001, ** p<0.01, * p<0.05 | | | | | | |

##### 1.1.2. Pairwise comparisons

By prompts per day

F1-score pairwise comparisons by prompts per day (Overall)

| **Contrast** | **Estimate** | **Standard error** | **z-ratio** | **p-value** | **Sig.** |
| --- | --- | --- | --- | --- | --- |
| EMAs16 - EMAs6 | -0.019 | 0.015 | -1.310 | 0.685 |  |
| EMAs16 - EMAs5 | 0.016 | 0.015 | 1.077 | 0.818 |  |
| EMAs16 - EMAs4 | -0.008 | 0.015 | -0.524 | 0.985 |  |
| EMAs16 - EMAs3 | -0.038 | 0.015 | -2.491 | 0.093 |  |
| EMAs6 - EMAs5 | 0.035 | 0.015 | 2.331 | 0.135 |  |
| EMAs6 - EMAs4 | 0.011 | 0.015 | 0.752 | 0.944 |  |
| EMAs6 - EMAs3 | -0.019 | 0.015 | -1.220 | 0.740 |  |
| EMAs5 - EMAs4 | -0.024 | 0.015 | -1.561 | 0.523 |  |
| EMAs5 - EMAs3 | -0.054 | 0.016 | -3.460 | 0.005 | ** |
| EMAs4 - EMAs3 | -0.030 | 0.016 | -1.931 | 0.301 |  |
| Model: Zero-inflated beta regression. Estimates shown are for the conditional component only (values between 0 and 1), excluding the zero-inflation process. | | | | | |
| Note: Estimates are on the response scale (0-1), showing differences in predicted means. | | | | | |
| Positive estimates indicate higher values for the first condition in each contrast; negative estimates indicate higher values for the second condition. | | | | | |
| Significance levels: *** p<0.001, ** p<0.01, * p<0.05; P-values adjusted using Tukey method. | | | | | |

By feature selection

F1-score pairwise comparisons by feature selection (Overall)

| **Contrast** | **Estimate** | **Standard error** | **z-ratio** | **p-value** | **Sig.** |
| --- | --- | --- | --- | --- | --- |
| All features - Selected features | -0.016 | 0.010 | -1.679 | 0.093 |  |
| Model: Zero-inflated beta regression. Estimates shown are for the conditional component only (values between 0 and 1), excluding the zero-inflation process. | | | | | |
| Note: Estimates are on the response scale (0-1), showing differences in predicted means. | | | | | |
| Positive estimates indicate higher values for the first condition in each contrast; negative estimates indicate higher values for the second condition. | | | | | |
| Significance levels: *** p<0.001, ** p<0.01, * p<0.05; P-values adjusted using Tukey method. | | | | | |

By share of own data in training set

F1-score pairwise comparisons by share of own data in training set (Overall)

| **Contrast** | **Estimate** | **Standard error** | **z-ratio** | **p-value** | **Sig.** |
| --- | --- | --- | --- | --- | --- |
| None - 10% | -0.018 | 0.013 | -1.307 | 0.559 |  |
| None - 20% | -0.032 | 0.014 | -2.329 | 0.091 |  |
| None - 30% | -0.066 | 0.014 | -4.892 | <0.001 | *** |
| 10% - 20% | -0.014 | 0.014 | -1.038 | 0.727 |  |
| 10% - 30% | -0.049 | 0.014 | -3.605 | 0.002 | ** |
| 20% - 30% | -0.035 | 0.014 | -2.549 | 0.053 |  |
| Model: Zero-inflated beta regression. Estimates shown are for the conditional component only (values between 0 and 1), excluding the zero-inflation process. | | | | | |
| Note: Estimates are on the response scale (0-1), showing differences in predicted means. | | | | | |
| Positive estimates indicate higher values for the first condition in each contrast; negative estimates indicate higher values for the second condition. | | | | | |
| Significance levels: *** p<0.001, ** p<0.01, * p<0.05; P-values adjusted using Tukey method. | | | | | |

By outcome

F1-score pairwise comparisons by outcome (Overall)

| **Contrast** | **Estimate** | **Standard error** | **z-ratio** | **p-value** | **Sig.** |
| --- | --- | --- | --- | --- | --- |
| Lapses - Cravings | 0.023 | 0.012 | 1.875 | 0.061 |  |
| Model: Zero-inflated beta regression. Estimates shown are for the conditional component only (values between 0 and 1), excluding the zero-inflation process. | | | | | |
| Note: Estimates are on the response scale (0-1), showing differences in predicted means. | | | | | |
| Positive estimates indicate higher values for the first condition in each contrast; negative estimates indicate higher values for the second condition. | | | | | |
| Significance levels: *** p<0.001, ** p<0.01, * p<0.05; P-values adjusted using Tukey method. | | | | | |

#### 1.2. Lapses models

##### 1.2.1. Main model results

F1-score (Lapses) - zero-inflated beta regression (with random effects)

| **Predictor** | **Estimate (logit scale)** | **Standard error** | **95% confidence interval (logit)** | **z-statistic** | **p-value** | **Sig.** |
| --- | --- | --- | --- | --- | --- | --- |
| Intercept | -0.785 | 0.203 | [-1.183, -0.386] | -3.859 | <0.001 | *** |
| Prompts per day: 6 | 0.460 | 0.075 | [0.313, 0.606] | 6.162 | <0.001 | *** |
| Prompts per day: 5 | 0.411 | 0.074 | [0.266, 0.557] | 5.537 | <0.001 | *** |
| Prompts per day: 4 | 0.655 | 0.075 | [0.508, 0.801] | 8.753 | <0.001 | *** |
| Prompts per day: 3 | 0.981 | 0.076 | [0.833, 1.130] | 12.957 | <0.001 | *** |
| Feature selection: Selected features | 0.097 | 0.047 | [0.006, 0.188] | 2.079 | 0.038 | * |
| Share of own data: 10% | 0.091 | 0.065 | [-0.037, 0.219] | 1.395 | 0.163 |  |
| Share of own data: 20% | 0.198 | 0.065 | [0.072, 0.325] | 3.071 | 0.002 | ** |
| Share of own data: 30% | 0.384 | 0.067 | [0.253, 0.514] | 5.767 | <0.001 | *** |
| Reference levels: Prompts per day = 16; Feature selection = All features; Share of own data = None | | | | | | |
| Note: Estimate coefficients are on the logit scale (conditional component only). | | | | | | |
| This is a zero-inflated beta model with separate processes for zeros and values between 0 and 1. | | | | | | |
| For interpretable effect sizes (actual differences in predicted values), see pairwise comparisons on the response scale (0-1). | | | | | | |
| Significance levels: *** p<0.001, ** p<0.01, * p<0.05 | | | | | | |

##### 1.2.2. Pairwise comparisons

By prompts per day

F1-score pairwise comparisons by prompts per day (Lapses)

| **Contrast** | **Estimate** | **Standard error** | **z-ratio** | **p-value** | **Sig.** |
| --- | --- | --- | --- | --- | --- |
| EMAs16 - EMAs6 | -0.111 | 0.018 | -6.095 | <0.001 | *** |
| EMAs16 - EMAs5 | -0.099 | 0.018 | -5.477 | <0.001 | *** |
| EMAs16 - EMAs4 | -0.160 | 0.018 | -8.729 | <0.001 | *** |
| EMAs16 - EMAs3 | -0.240 | 0.018 | -13.418 | <0.001 | *** |
| EMAs6 - EMAs5 | 0.012 | 0.018 | 0.671 | 0.963 |  |
| EMAs6 - EMAs4 | -0.049 | 0.018 | -2.681 | 0.057 |  |
| EMAs6 - EMAs3 | -0.129 | 0.018 | -7.124 | <0.001 | *** |
| EMAs5 - EMAs4 | -0.061 | 0.018 | -3.376 | 0.007 | ** |
| EMAs5 - EMAs3 | -0.141 | 0.018 | -7.853 | <0.001 | *** |
| EMAs4 - EMAs3 | -0.080 | 0.018 | -4.429 | <0.001 | *** |
| Model: Zero-inflated beta regression. Estimates shown are for the conditional component only (values between 0 and 1), excluding the zero-inflation process. | | | | | |
| Note: Estimates are on the response scale (0-1), showing differences in predicted means. | | | | | |
| Positive estimates indicate higher values for the first condition in each contrast; negative estimates indicate higher values for the second condition. | | | | | |
| Significance levels: *** p<0.001, ** p<0.01, * p<0.05; P-values adjusted using Tukey method. | | | | | |

By feature selection

F1-score pairwise comparisons by feature selection (Lapses)

| **Contrast** | **Estimate** | **Standard error** | **z-ratio** | **p-value** | **Sig.** |
| --- | --- | --- | --- | --- | --- |
| All features - Selected features | -0.024 | 0.012 | -2.080 | 0.038 | * |
| Model: Zero-inflated beta regression. Estimates shown are for the conditional component only (values between 0 and 1), excluding the zero-inflation process. | | | | | |
| Note: Estimates are on the response scale (0-1), showing differences in predicted means. | | | | | |
| Positive estimates indicate higher values for the first condition in each contrast; negative estimates indicate higher values for the second condition. | | | | | |
| Significance levels: *** p<0.001, ** p<0.01, * p<0.05; P-values adjusted using Tukey method. | | | | | |

By share of own data in training set

F1-score pairwise comparisons by share of own data in training set (Lapses)

| **Contrast** | **Estimate** | **Standard error** | **z-ratio** | **p-value** | **Sig.** |
| --- | --- | --- | --- | --- | --- |
| None - 10% | -0.023 | 0.016 | -1.395 | 0.503 |  |
| None - 20% | -0.049 | 0.016 | -3.072 | 0.011 | * |
| None - 30% | -0.096 | 0.016 | -5.797 | <0.001 | *** |
| 10% - 20% | -0.027 | 0.016 | -1.636 | 0.358 |  |
| 10% - 30% | -0.073 | 0.017 | -4.362 | <0.001 | *** |
| 20% - 30% | -0.046 | 0.017 | -2.775 | 0.028 | * |
| Model: Zero-inflated beta regression. Estimates shown are for the conditional component only (values between 0 and 1), excluding the zero-inflation process. | | | | | |
| Note: Estimates are on the response scale (0-1), showing differences in predicted means. | | | | | |
| Positive estimates indicate higher values for the first condition in each contrast; negative estimates indicate higher values for the second condition. | | | | | |
| Significance levels: *** p<0.001, ** p<0.01, * p<0.05; P-values adjusted using Tukey method. | | | | | |

#### 1.3. Cravings models

##### 1.3.1. Main model results

F1-score (Cravings) - zero-inflated beta regression (with random effects)

| **Predictor** | **Estimate (logit scale)** | **Standard error** | **95% confidence interval (logit)** | **z-statistic** | **p-value** | **Sig.** |
| --- | --- | --- | --- | --- | --- | --- |
| Intercept | 0.278 | 0.230 | [-0.173, 0.728] | 1.208 | 0.227 |  |
| Prompts per day: 6 | -0.142 | 0.056 | [-0.253, -0.032] | -2.523 | 0.012 | * |
| Prompts per day: 5 | -0.398 | 0.059 | [-0.514, -0.282] | -6.746 | <0.001 | *** |
| Prompts per day: 4 | -0.346 | 0.059 | [-0.461, -0.231] | -5.882 | <0.001 | *** |
| Prompts per day: 3 | -0.381 | 0.060 | [-0.499, -0.262] | -6.304 | <0.001 | *** |
| Feature selection: Selected features | 0.021 | 0.037 | [-0.053, 0.094] | 0.555 | 0.579 |  |
| Share of own data: 10% | -0.057 | 0.053 | [-0.160, 0.046] | -1.085 | 0.278 |  |
| Share of own data: 20% | -0.076 | 0.054 | [-0.181, 0.029] | -1.423 | 0.155 |  |
| Share of own data: 30% | 0.077 | 0.053 | [-0.027, 0.181] | 1.452 | 0.146 |  |
| Reference levels: Prompts per day = 16; Feature selection = All features; Share of own data = None | | | | | | |
| Note: Estimate coefficients are on the logit scale (conditional component only). | | | | | | |
| This is a zero-inflated beta model with separate processes for zeros and values between 0 and 1. | | | | | | |
| For interpretable effect sizes (actual differences in predicted values), see pairwise comparisons on the response scale (0-1). | | | | | | |
| Significance levels: *** p<0.001, ** p<0.01, * p<0.05 | | | | | | |

##### 1.3.2. Pairwise comparisons

By prompts per day

F1-score pairwise comparisons by prompts per day (Cravings)

| **Contrast** | **Estimate** | **Standard error** | **z-ratio** | **p-value** | **Sig.** |
| --- | --- | --- | --- | --- | --- |
| EMAs16 - EMAs6 | 0.035 | 0.014 | 2.520 | 0.086 |  |
| EMAs16 - EMAs5 | 0.099 | 0.015 | 6.772 | <0.001 | *** |
| EMAs16 - EMAs4 | 0.086 | 0.015 | 5.891 | <0.001 | *** |
| EMAs16 - EMAs3 | 0.095 | 0.015 | 6.321 | <0.001 | *** |
| EMAs6 - EMAs5 | 0.064 | 0.015 | 4.330 | <0.001 | *** |
| EMAs6 - EMAs4 | 0.051 | 0.015 | 3.430 | 0.005 | ** |
| EMAs6 - EMAs3 | 0.060 | 0.015 | 3.953 | <0.001 | *** |
| EMAs5 - EMAs4 | -0.013 | 0.015 | -0.854 | 0.913 |  |
| EMAs5 - EMAs3 | -0.004 | 0.016 | -0.280 | 0.999 |  |
| EMAs4 - EMAs3 | 0.009 | 0.016 | 0.558 | 0.981 |  |
| Model: Zero-inflated beta regression. Estimates shown are for the conditional component only (values between 0 and 1), excluding the zero-inflation process. | | | | | |
| Note: Estimates are on the response scale (0-1), showing differences in predicted means. | | | | | |
| Positive estimates indicate higher values for the first condition in each contrast; negative estimates indicate higher values for the second condition. | | | | | |
| Significance levels: *** p<0.001, ** p<0.01, * p<0.05; P-values adjusted using Tukey method. | | | | | |

By feature selection

F1-score pairwise comparisons by feature selection (Cravings)

| **Contrast** | **Estimate** | **Standard error** | **z-ratio** | **p-value** | **Sig.** |
| --- | --- | --- | --- | --- | --- |
| All features - Selected features | -0.005 | 0.009 | -0.555 | 0.579 |  |
| Model: Zero-inflated beta regression. Estimates shown are for the conditional component only (values between 0 and 1), excluding the zero-inflation process. | | | | | |
| Note: Estimates are on the response scale (0-1), showing differences in predicted means. | | | | | |
| Positive estimates indicate higher values for the first condition in each contrast; negative estimates indicate higher values for the second condition. | | | | | |
| Significance levels: *** p<0.001, ** p<0.01, * p<0.05; P-values adjusted using Tukey method. | | | | | |

By share of own data in training set

F1-score pairwise comparisons by share of own data in training set (Cravings)

| **Contrast** | **Estimate** | **Standard error** | **z-ratio** | **p-value** | **Sig.** |
| --- | --- | --- | --- | --- | --- |
| None - 10% | 0.014 | 0.013 | 1.085 | 0.699 |  |
| None - 20% | 0.019 | 0.013 | 1.424 | 0.485 |  |
| None - 30% | -0.019 | 0.013 | -1.453 | 0.467 |  |
| 10% - 20% | 0.005 | 0.013 | 0.359 | 0.984 |  |
| 10% - 30% | -0.033 | 0.013 | -2.536 | 0.055 |  |
| 20% - 30% | -0.038 | 0.013 | -2.875 | 0.021 | * |
| Model: Zero-inflated beta regression. Estimates shown are for the conditional component only (values between 0 and 1), excluding the zero-inflation process. | | | | | |
| Note: Estimates are on the response scale (0-1), showing differences in predicted means. | | | | | |
| Positive estimates indicate higher values for the first condition in each contrast; negative estimates indicate higher values for the second condition. | | | | | |
| Significance levels: *** p<0.001, ** p<0.01, * p<0.05; P-values adjusted using Tukey method. | | | | | |
