## Supplementary material for "Optimising supervised machine learning algorithms predicting cigarette cravings and lapses for a smoking cessation just-in-time adaptive intervention (JITAI)": S9 Appendix ROC-AUC models and marginal effects

### 1. Comprehensive analysis: ROC-AUC

#### 1.1. Overall models

##### 1.1.1. Main model results

ROC-AUC (Overall) (linear mixed model)

| **Predictor** | **Estimate** | **Standard error** | **95% confidence interval** | **t-statistic** | **p-value** | **Sig.** |
| --- | --- | --- | --- | --- | --- | --- |
| Intercept | 0.726 | 0.018 | [0.690, 0.762] | 39.653 | <0.001 | *** |
| Prompts per day: 6 | -0.044 | 0.010 | [-0.064, -0.023] | -4.206 | <0.001 | *** |
| Prompts per day: 5 | -0.066 | 0.010 | [-0.087, -0.046] | -6.345 | <0.001 | *** |
| Prompts per day: 4 | -0.074 | 0.010 | [-0.094, -0.054] | -7.168 | <0.001 | *** |
| Prompts per day: 3 | -0.095 | 0.010 | [-0.116, -0.075] | -9.239 | <0.001 | *** |
| Feature selection: Selected features | -0.020 | 0.007 | [-0.033, -0.007] | -3.047 | 0.002 | ** |
| Share of own data: 10% | -0.024 | 0.009 | [-0.042, -0.006] | -2.649 | 0.008 | ** |
| Share of own data: 20% | -0.011 | 0.009 | [-0.029, 0.007] | -1.180 | 0.238 |  |
| Share of own data: 30% | -0.017 | 0.009 | [-0.035, 0.002] | -1.793 | 0.073 |  |
| Outcome: Cravings | -0.036 | 0.008 | [-0.051, -0.021] | -4.747 | <0.001 | *** |
| Reference levels: Prompts per day = 16; Feature selection = All features; Share of own data = None; Outcome = Lapses | | | | | | |
| Significance levels: *** p<0.001, ** p<0.01, * p<0.05 | | | | | | |

##### 1.1.2. Pairwise comparisons

By prompts per day

ROC-AUC pairwise comparisons by prompts per day (Overall)

| **Contrast** | **Estimate** | **Standard error** | **t-ratio** | **p-value** | **Sig.** |
| --- | --- | --- | --- | --- | --- |
| EMAs16 - EMAs6 | 0.044 | 0.010 | 4.205 | <0.001 | *** |
| EMAs16 - EMAs5 | 0.066 | 0.010 | 6.345 | <0.001 | *** |
| EMAs16 - EMAs4 | 0.074 | 0.010 | 7.168 | <0.001 | *** |
| EMAs16 - EMAs3 | 0.095 | 0.010 | 9.239 | <0.001 | *** |
| EMAs6 - EMAs5 | 0.022 | 0.011 | 2.140 | 0.203 |  |
| EMAs6 - EMAs4 | 0.030 | 0.010 | 2.906 | 0.030 | * |
| EMAs6 - EMAs3 | 0.052 | 0.010 | 4.977 | <0.001 | *** |
| EMAs5 - EMAs4 | 0.008 | 0.010 | 0.741 | 0.947 |  |
| EMAs5 - EMAs3 | 0.029 | 0.010 | 2.808 | 0.040 | * |
| EMAs4 - EMAs3 | 0.022 | 0.010 | 2.090 | 0.225 |  |
| Note: Estimates are on the response scale (0-1), showing differences in predicted means. | | | | | |
| Positive estimates indicate higher values for the first condition in each contrast; negative estimates indicate higher values for the second condition. | | | | | |
| Significance levels: *** p<0.001, ** p<0.01, * p<0.05; P-values adjusted using Tukey method. | | | | | |

By feature selection

ROC-AUC pairwise comparisons by feature selection (Overall)

| **Contrast** | **Estimate** | **Standard error** | **t-ratio** | **p-value** | **Sig.** |
| --- | --- | --- | --- | --- | --- |
| All features - Selected features | 0.020 | 0.007 | 3.047 | 0.002 | ** |
| Note: Estimates are on the response scale (0-1), showing differences in predicted means. | | | | | |
| Positive estimates indicate higher values for the first condition in each contrast; negative estimates indicate higher values for the second condition. | | | | | |
| Significance levels: *** p<0.001, ** p<0.01, * p<0.05; P-values adjusted using Tukey method. | | | | | |

By share of own data in training set

ROC-AUC pairwise comparisons by share of own data in training set (Overall)

| **Contrast** | **Estimate** | **Standard error** | **t-ratio** | **p-value** | **Sig.** |
| --- | --- | --- | --- | --- | --- |
| None - 10% | 0.024 | 0.009 | 2.649 | 0.041 | * |
| None - 20% | 0.011 | 0.009 | 1.180 | 0.640 |  |
| None - 30% | 0.017 | 0.009 | 1.793 | 0.277 |  |
| 10% - 20% | -0.013 | 0.009 | -1.401 | 0.499 |  |
| 10% - 30% | -0.008 | 0.009 | -0.796 | 0.856 |  |
| 20% - 30% | 0.006 | 0.010 | 0.598 | 0.933 |  |
| Note: Estimates are on the response scale (0-1), showing differences in predicted means. | | | | | |
| Positive estimates indicate higher values for the first condition in each contrast; negative estimates indicate higher values for the second condition. | | | | | |
| Significance levels: *** p<0.001, ** p<0.01, * p<0.05; P-values adjusted using Tukey method. | | | | | |

By outcome

ROC-AUC pairwise comparisons by outcome (Overall)

| **Contrast** | **Estimate** | **Standard error** | **t-ratio** | **p-value** | **Sig.** |
| --- | --- | --- | --- | --- | --- |
| Lapses - Cravings | 0.036 | 0.008 | 4.743 | <0.001 | *** |
| Note: Estimates are on the response scale (0-1), showing differences in predicted means. | | | | | |
| Positive estimates indicate higher values for the first condition in each contrast; negative estimates indicate higher values for the second condition. | | | | | |
| Significance levels: *** p<0.001, ** p<0.01, * p<0.05; P-values adjusted using Tukey method. | | | | | |

#### 1.2. Lapses models

##### 1.2.1. Main model results

ROC-AUC (Lapses) (linear mixed model)

| **Predictor** | **Estimate** | **Standard error** | **95% confidence interval** | **t-statistic** | **p-value** | **Sig.** |
| --- | --- | --- | --- | --- | --- | --- |
| Intercept | 0.677 | 0.039 | [0.601, 0.754] | 17.373 | <0.001 | *** |
| Prompts per day: 6 | -0.042 | 0.011 | [-0.064, -0.020] | -3.749 | <0.001 | *** |
| Prompts per day: 5 | -0.034 | 0.011 | [-0.056, -0.012] | -2.986 | 0.003 | ** |
| Prompts per day: 4 | -0.048 | 0.011 | [-0.070, -0.026] | -4.288 | <0.001 | *** |
| Prompts per day: 3 | -0.014 | 0.011 | [-0.036, 0.008] | -1.212 | 0.226 |  |
| Feature selection: Selected features | -0.009 | 0.007 | [-0.023, 0.005] | -1.215 | 0.224 |  |
| Share of own data: 10% | -0.016 | 0.010 | [-0.036, 0.003] | -1.615 | 0.106 |  |
| Share of own data: 20% | 0.004 | 0.011 | [-0.017, 0.025] | 0.384 | 0.701 |  |
| Share of own data: 30% | 0.001 | 0.011 | [-0.020, 0.021] | 0.057 | 0.954 |  |
| Reference levels: Prompts per day = 16; Feature selection = All features; Share of own data = None | | | | | | |
| Significance levels: *** p<0.001, ** p<0.01, * p<0.05 | | | | | | |

##### 1.2.2. Pairwise comparisons

By prompts per day

ROC-AUC pairwise comparisons by prompts per day (Lapses)

| **Contrast** | **Estimate** | **Standard error** | **t-ratio** | **p-value** | **Sig.** |
| --- | --- | --- | --- | --- | --- |
| EMAs16 - EMAs6 | 0.042 | 0.011 | 3.749 | 0.002 | ** |
| EMAs16 - EMAs5 | 0.034 | 0.011 | 2.986 | 0.024 | * |
| EMAs16 - EMAs4 | 0.048 | 0.011 | 4.288 | <0.001 | *** |
| EMAs16 - EMAs3 | 0.014 | 0.011 | 1.212 | 0.745 |  |
| EMAs6 - EMAs5 | -0.009 | 0.011 | -0.763 | 0.941 |  |
| EMAs6 - EMAs4 | 0.006 | 0.011 | 0.539 | 0.983 |  |
| EMAs6 - EMAs3 | -0.029 | 0.011 | -2.523 | 0.087 |  |
| EMAs5 - EMAs4 | 0.015 | 0.011 | 1.302 | 0.690 |  |
| EMAs5 - EMAs3 | -0.020 | 0.011 | -1.762 | 0.396 |  |
| EMAs4 - EMAs3 | -0.035 | 0.011 | -3.060 | 0.019 | * |
| Note: Estimates are on the response scale (0-1), showing differences in predicted means. | | | | | |
| Positive estimates indicate higher values for the first condition in each contrast; negative estimates indicate higher values for the second condition. | | | | | |
| Significance levels: *** p<0.001, ** p<0.01, * p<0.05; P-values adjusted using Tukey method. | | | | | |

By feature selection

ROC-AUC pairwise comparisons by feature selection (Lapses)

| **Contrast** | **Estimate** | **Standard error** | **t-ratio** | **p-value** | **Sig.** |
| --- | --- | --- | --- | --- | --- |
| All features - Selected features | 0.009 | 0.007 | 1.215 | 0.225 |  |
| Note: Estimates are on the response scale (0-1), showing differences in predicted means. | | | | | |
| Positive estimates indicate higher values for the first condition in each contrast; negative estimates indicate higher values for the second condition. | | | | | |
| Significance levels: *** p<0.001, ** p<0.01, * p<0.05; P-values adjusted using Tukey method. | | | | | |

By share of own data in training set

ROC-AUC pairwise comparisons by share of own data in training set (Lapses)

| **Contrast** | **Estimate** | **Standard error** | **t-ratio** | **p-value** | **Sig.** |
| --- | --- | --- | --- | --- | --- |
| None - 10% | 0.016 | 0.010 | 1.614 | 0.371 |  |
| None - 20% | -0.004 | 0.011 | -0.384 | 0.981 |  |
| None - 30% | -0.001 | 0.011 | -0.057 | 1.000 |  |
| 10% - 20% | -0.020 | 0.011 | -1.902 | 0.228 |  |
| 10% - 30% | -0.017 | 0.011 | -1.576 | 0.393 |  |
| 20% - 30% | 0.003 | 0.011 | 0.320 | 0.989 |  |
| Note: Estimates are on the response scale (0-1), showing differences in predicted means. | | | | | |
| Positive estimates indicate higher values for the first condition in each contrast; negative estimates indicate higher values for the second condition. | | | | | |
| Significance levels: *** p<0.001, ** p<0.01, * p<0.05; P-values adjusted using Tukey method. | | | | | |

#### 1.3. Cravings models

##### 1.3.1. Main model results

ROC-AUC (Cravings) (linear mixed model)

| **Predictor** | **Estimate** | **Standard error** | **95% confidence interval** | **t-statistic** | **p-value** | **Sig.** |
| --- | --- | --- | --- | --- | --- | --- |
| Intercept | 0.714 | 0.018 | [0.678, 0.749] | 39.470 | <0.001 | *** |
| Prompts per day: 6 | -0.046 | 0.012 | [-0.070, -0.023] | -3.812 | <0.001 | *** |
| Prompts per day: 5 | -0.085 | 0.012 | [-0.110, -0.061] | -6.964 | <0.001 | *** |
| Prompts per day: 4 | -0.088 | 0.012 | [-0.111, -0.064] | -7.260 | <0.001 | *** |
| Prompts per day: 3 | -0.139 | 0.012 | [-0.163, -0.115] | -11.489 | <0.001 | *** |
| Feature selection: Selected features | -0.026 | 0.008 | [-0.041, -0.011] | -3.403 | <0.001 | *** |
| Share of own data: 10% | -0.023 | 0.011 | [-0.045, -0.002] | -2.156 | 0.031 | * |
| Share of own data: 20% | -0.023 | 0.011 | [-0.044, -0.002] | -2.105 | 0.035 | * |
| Share of own data: 30% | -0.030 | 0.011 | [-0.051, -0.008] | -2.726 | 0.006 | ** |
| Reference levels: Prompts per day = 16; Feature selection = All features; Share of own data = None | | | | | | |
| Significance levels: *** p<0.001, ** p<0.01, * p<0.05 | | | | | | |

##### 1.3.2. Pairwise comparisons

By prompts per day

ROC-AUC pairwise comparisons by prompts per day (Cravings)

| **Contrast** | **Estimate** | **Standard error** | **t-ratio** | **p-value** | **Sig.** |
| --- | --- | --- | --- | --- | --- |
| EMAs16 - EMAs6 | 0.046 | 0.012 | 3.812 | 0.001 | ** |
| EMAs16 - EMAs5 | 0.085 | 0.012 | 6.964 | <0.001 | *** |
| EMAs16 - EMAs4 | 0.088 | 0.012 | 7.260 | <0.001 | *** |
| EMAs16 - EMAs3 | 0.139 | 0.012 | 11.488 | <0.001 | *** |
| EMAs6 - EMAs5 | 0.039 | 0.012 | 3.144 | 0.015 | * |
| EMAs6 - EMAs4 | 0.041 | 0.012 | 3.362 | 0.007 | ** |
| EMAs6 - EMAs3 | 0.092 | 0.012 | 7.563 | <0.001 | *** |
| EMAs5 - EMAs4 | 0.002 | 0.012 | 0.165 | 1.000 |  |
| EMAs5 - EMAs3 | 0.053 | 0.012 | 4.344 | <0.001 | *** |
| EMAs4 - EMAs3 | 0.051 | 0.012 | 4.248 | <0.001 | *** |
| Note: Estimates are on the response scale (0-1), showing differences in predicted means. | | | | | |
| Positive estimates indicate higher values for the first condition in each contrast; negative estimates indicate higher values for the second condition. | | | | | |
| Significance levels: *** p<0.001, ** p<0.01, * p<0.05; P-values adjusted using Tukey method. | | | | | |

By feature selection

ROC-AUC pairwise comparisons by feature selection (Cravings)

| **Contrast** | **Estimate** | **Standard error** | **t-ratio** | **p-value** | **Sig.** |
| --- | --- | --- | --- | --- | --- |
| All features - Selected features | 0.026 | 0.008 | 3.403 | <0.001 | *** |
| Note: Estimates are on the response scale (0-1), showing differences in predicted means. | | | | | |
| Positive estimates indicate higher values for the first condition in each contrast; negative estimates indicate higher values for the second condition. | | | | | |
| Significance levels: *** p<0.001, ** p<0.01, * p<0.05; P-values adjusted using Tukey method. | | | | | |

By share of own data in training set

ROC-AUC pairwise comparisons by share of own data in training set (Cravings)

| **Contrast** | **Estimate** | **Standard error** | **t-ratio** | **p-value** | **Sig.** |
| --- | --- | --- | --- | --- | --- |
| None - 10% | 0.023 | 0.011 | 2.156 | 0.136 |  |
| None - 20% | 0.023 | 0.011 | 2.105 | 0.152 |  |
| None - 30% | 0.030 | 0.011 | 2.726 | 0.033 | * |
| 10% - 20% | -0.001 | 0.011 | -0.046 | 1.000 |  |
| 10% - 30% | 0.006 | 0.011 | 0.574 | 0.940 |  |
| 20% - 30% | 0.007 | 0.011 | 0.619 | 0.926 |  |
| Note: Estimates are on the response scale (0-1), showing differences in predicted means. | | | | | |
| Positive estimates indicate higher values for the first condition in each contrast; negative estimates indicate higher values for the second condition. | | | | | |
| Significance levels: *** p<0.001, ** p<0.01, * p<0.05; P-values adjusted using Tukey method. | | | | | |
