## Supplementary material for "Optimising supervised machine learning algorithms predicting cigarette cravings and lapses for a smoking cessation just-in-time adaptive intervention (JITAI)": S10 Appendix Sensitivity models and marginal effects

### 1. Comprehensive analysis: Sensitivity

#### 1.1. Overall models

##### 1.1.1. Main model results

Sensitivity (Overall) (linear mixed model)

| **Predictor** | **Estimate** | **Standard error** | **95% confidence interval** | **t-statistic** | **p-value** | **Sig.** |
| --- | --- | --- | --- | --- | --- | --- |
| Intercept | 0.434 | 0.049 | [0.338, 0.529] | 8.897 | <0.001 | *** |
| Prompts per day: 6 | 0.018 | 0.019 | [-0.020, 0.056] | 0.908 | 0.364 |  |
| Prompts per day: 5 | -0.028 | 0.020 | [-0.066, 0.011] | -1.420 | 0.156 |  |
| Prompts per day: 4 | -0.009 | 0.019 | [-0.047, 0.029] | -0.449 | 0.653 |  |
| Prompts per day: 3 | 0.021 | 0.019 | [-0.017, 0.059] | 1.083 | 0.279 |  |
| Feature selection: Selected features | 0.012 | 0.012 | [-0.012, 0.036] | 0.998 | 0.318 |  |
| Share of own data: 10% | 0.056 | 0.017 | [0.023, 0.090] | 3.280 | 0.001 | ** |
| Share of own data: 20% | 0.045 | 0.017 | [0.011, 0.080] | 2.609 | 0.009 | ** |
| Share of own data: 30% | 0.093 | 0.017 | [0.059, 0.127] | 5.328 | <0.001 | *** |
| Outcome: Cravings | -0.031 | 0.014 | [-0.059, -0.003] | -2.175 | 0.030 | * |
| Reference levels: Prompts per day = 16; Feature selection = All features; Share of own data = None; Outcome = Lapses | | | | | | |
| Significance levels: *** p<0.001, ** p<0.01, * p<0.05 | | | | | | |

##### 1.1.2. Pairwise comparisons

By prompts per day

Sensitivity pairwise comparisons by prompts per day (Overall)

| **Contrast** | **Estimate** | **Standard error** | **t-ratio** | **p-value** | **Sig.** |
| --- | --- | --- | --- | --- | --- |
| EMAs16 - EMAs6 | -0.018 | 0.019 | -0.908 | 0.894 |  |
| EMAs16 - EMAs5 | 0.028 | 0.020 | 1.420 | 0.615 |  |
| EMAs16 - EMAs4 | 0.009 | 0.019 | 0.449 | 0.992 |  |
| EMAs16 - EMAs3 | -0.021 | 0.019 | -1.083 | 0.815 |  |
| EMAs6 - EMAs5 | 0.045 | 0.020 | 2.307 | 0.143 |  |
| EMAs6 - EMAs4 | 0.026 | 0.019 | 1.352 | 0.659 |  |
| EMAs6 - EMAs3 | -0.003 | 0.020 | -0.170 | 1.000 |  |
| EMAs5 - EMAs4 | -0.019 | 0.020 | -0.974 | 0.867 |  |
| EMAs5 - EMAs3 | -0.049 | 0.020 | -2.486 | 0.094 |  |
| EMAs4 - EMAs3 | -0.030 | 0.019 | -1.529 | 0.544 |  |
| Note: Estimates are on the response scale (0-1), showing differences in predicted means. | | | | | |
| Positive estimates indicate higher values for the first condition in each contrast; negative estimates indicate higher values for the second condition. | | | | | |
| Significance levels: *** p<0.001, ** p<0.01, * p<0.05; P-values adjusted using Tukey method. | | | | | |

By feature selection

Sensitivity pairwise comparisons by feature selection (Overall)

| **Contrast** | **Estimate** | **Standard error** | **t-ratio** | **p-value** | **Sig.** |
| --- | --- | --- | --- | --- | --- |
| All features - Selected features | -0.012 | 0.012 | -0.998 | 0.318 |  |
| Note: Estimates are on the response scale (0-1), showing differences in predicted means. | | | | | |
| Positive estimates indicate higher values for the first condition in each contrast; negative estimates indicate higher values for the second condition. | | | | | |
| Significance levels: *** p<0.001, ** p<0.01, * p<0.05; P-values adjusted using Tukey method. | | | | | |

By share of own data in training set

Sensitivity pairwise comparisons by share of own data in training set (Overall)

| **Contrast** | **Estimate** | **Standard error** | **t-ratio** | **p-value** | **Sig.** |
| --- | --- | --- | --- | --- | --- |
| None - 10% | -0.056 | 0.017 | -3.280 | 0.006 | ** |
| None - 20% | -0.045 | 0.017 | -2.609 | 0.045 | * |
| None - 30% | -0.093 | 0.017 | -5.328 | <0.001 | *** |
| 10% - 20% | 0.011 | 0.018 | 0.601 | 0.932 |  |
| 10% - 30% | -0.037 | 0.018 | -2.081 | 0.160 |  |
| 20% - 30% | -0.047 | 0.018 | -2.652 | 0.040 | * |
| Note: Estimates are on the response scale (0-1), showing differences in predicted means. | | | | | |
| Positive estimates indicate higher values for the first condition in each contrast; negative estimates indicate higher values for the second condition. | | | | | |
| Significance levels: *** p<0.001, ** p<0.01, * p<0.05; P-values adjusted using Tukey method. | | | | | |

By outcome

Sensitivity pairwise comparisons by outcome (Overall)

| **Contrast** | **Estimate** | **Standard error** | **t-ratio** | **p-value** | **Sig.** |
| --- | --- | --- | --- | --- | --- |
| Lapses - Cravings | 0.031 | 0.014 | 2.175 | 0.030 | * |
| Note: Estimates are on the response scale (0-1), showing differences in predicted means. | | | | | |
| Positive estimates indicate higher values for the first condition in each contrast; negative estimates indicate higher values for the second condition. | | | | | |
| Significance levels: *** p<0.001, ** p<0.01, * p<0.05; P-values adjusted using Tukey method. | | | | | |

#### 1.2. Lapses models

##### 1.2.1. Main model results

Sensitivity (Lapses) (linear mixed model)

| **Predictor** | **Estimate** | **Standard error** | **95% confidence interval** | **t-statistic** | **p-value** | **Sig.** |
| --- | --- | --- | --- | --- | --- | --- |
| Intercept | 0.272 | 0.063 | [0.148, 0.396] | 4.291 | <0.001 | *** |
| Prompts per day: 6 | 0.112 | 0.025 | [0.062, 0.162] | 4.419 | <0.001 | *** |
| Prompts per day: 5 | 0.087 | 0.025 | [0.037, 0.136] | 3.416 | <0.001 | *** |
| Prompts per day: 4 | 0.118 | 0.025 | [0.068, 0.167] | 4.639 | <0.001 | *** |
| Prompts per day: 3 | 0.155 | 0.025 | [0.105, 0.205] | 6.083 | <0.001 | *** |
| Feature selection: Selected features | 0.034 | 0.016 | [0.003, 0.065] | 2.118 | 0.034 | * |
| Share of own data: 10% | 0.049 | 0.022 | [0.006, 0.093] | 2.205 | 0.027 | * |
| Share of own data: 20% | 0.064 | 0.024 | [0.018, 0.111] | 2.699 | 0.007 | ** |
| Share of own data: 30% | 0.085 | 0.024 | [0.038, 0.131] | 3.562 | <0.001 | *** |
| Reference levels: Prompts per day = 16; Feature selection = All features; Share of own data = None | | | | | | |
| Significance levels: *** p<0.001, ** p<0.01, * p<0.05 | | | | | | |

##### 1.2.2. Pairwise comparisons

By prompts per day

Sensitivity pairwise comparisons by prompts per day (Lapses)

| **Contrast** | **Estimate** | **Standard error** | **t-ratio** | **p-value** | **Sig.** |
| --- | --- | --- | --- | --- | --- |
| EMAs16 - EMAs6 | -0.112 | 0.025 | -4.419 | <0.001 | *** |
| EMAs16 - EMAs5 | -0.087 | 0.025 | -3.416 | 0.006 | ** |
| EMAs16 - EMAs4 | -0.118 | 0.025 | -4.638 | <0.001 | *** |
| EMAs16 - EMAs3 | -0.155 | 0.025 | -6.082 | <0.001 | *** |
| EMAs6 - EMAs5 | 0.025 | 0.025 | 1.002 | 0.854 |  |
| EMAs6 - EMAs4 | -0.006 | 0.025 | -0.219 | 0.999 |  |
| EMAs6 - EMAs3 | -0.043 | 0.025 | -1.678 | 0.448 |  |
| EMAs5 - EMAs4 | -0.031 | 0.025 | -1.222 | 0.739 |  |
| EMAs5 - EMAs3 | -0.068 | 0.025 | -2.677 | 0.058 |  |
| EMAs4 - EMAs3 | -0.037 | 0.025 | -1.460 | 0.589 |  |
| Note: Estimates are on the response scale (0-1), showing differences in predicted means. | | | | | |
| Positive estimates indicate higher values for the first condition in each contrast; negative estimates indicate higher values for the second condition. | | | | | |
| Significance levels: *** p<0.001, ** p<0.01, * p<0.05; P-values adjusted using Tukey method. | | | | | |

By feature selection

Sensitivity pairwise comparisons by feature selection (Lapses)

| **Contrast** | **Estimate** | **Standard error** | **t-ratio** | **p-value** | **Sig.** |
| --- | --- | --- | --- | --- | --- |
| All features - Selected features | -0.034 | 0.016 | -2.118 | 0.035 | * |
| Note: Estimates are on the response scale (0-1), showing differences in predicted means. | | | | | |
| Positive estimates indicate higher values for the first condition in each contrast; negative estimates indicate higher values for the second condition. | | | | | |
| Significance levels: *** p<0.001, ** p<0.01, * p<0.05; P-values adjusted using Tukey method. | | | | | |

By share of own data in training set

Sensitivity pairwise comparisons by share of own data in training set (Lapses)

| **Contrast** | **Estimate** | **Standard error** | **t-ratio** | **p-value** | **Sig.** |
| --- | --- | --- | --- | --- | --- |
| None - 10% | -0.049 | 0.022 | -2.204 | 0.123 |  |
| None - 20% | -0.064 | 0.024 | -2.698 | 0.036 | * |
| None - 30% | -0.085 | 0.024 | -3.560 | 0.002 | ** |
| 10% - 20% | -0.015 | 0.024 | -0.620 | 0.926 |  |
| 10% - 30% | -0.035 | 0.024 | -1.481 | 0.450 |  |
| 20% - 30% | -0.021 | 0.024 | -0.844 | 0.833 |  |
| Note: Estimates are on the response scale (0-1), showing differences in predicted means. | | | | | |
| Positive estimates indicate higher values for the first condition in each contrast; negative estimates indicate higher values for the second condition. | | | | | |
| Significance levels: *** p<0.001, ** p<0.01, * p<0.05; P-values adjusted using Tukey method. | | | | | |

#### 1.3. Cravings models

##### 1.3.1. Main model results

Sensitivity (Cravings) (linear mixed model)

| **Predictor** | **Estimate** | **Standard error** | **95% confidence interval** | **t-statistic** | **p-value** | **Sig.** |
| --- | --- | --- | --- | --- | --- | --- |
| Intercept | 0.506 | 0.056 | [0.396, 0.616] | 8.995 | <0.001 | *** |
| Prompts per day: 6 | -0.038 | 0.018 | [-0.073, -0.003] | -2.147 | 0.032 | * |
| Prompts per day: 5 | -0.101 | 0.018 | [-0.136, -0.065] | -5.590 | <0.001 | *** |
| Prompts per day: 4 | -0.081 | 0.018 | [-0.116, -0.047] | -4.603 | <0.001 | *** |
| Prompts per day: 3 | -0.059 | 0.018 | [-0.094, -0.024] | -3.329 | <0.001 | *** |
| Feature selection: Selected features | 0.000 | 0.011 | [-0.022, 0.023] | 0.035 | 0.972 |  |
| Share of own data: 10% | 0.020 | 0.016 | [-0.011, 0.051] | 1.246 | 0.213 |  |
| Share of own data: 20% | -0.034 | 0.016 | [-0.065, -0.002] | -2.103 | 0.035 | * |
| Share of own data: 30% | 0.027 | 0.016 | [-0.005, 0.058] | 1.671 | 0.095 |  |
| Reference levels: Prompts per day = 16; Feature selection = All features; Share of own data = None | | | | | | |
| Significance levels: *** p<0.001, ** p<0.01, * p<0.05 | | | | | | |

##### 1.3.2. Pairwise comparisons

By prompts per day

Sensitivity pairwise comparisons by prompts per day (Cravings)

| **Contrast** | **Estimate** | **Standard error** | **t-ratio** | **p-value** | **Sig.** |
| --- | --- | --- | --- | --- | --- |
| EMAs16 - EMAs6 | 0.038 | 0.018 | 2.147 | 0.201 |  |
| EMAs16 - EMAs5 | 0.101 | 0.018 | 5.590 | <0.001 | *** |
| EMAs16 - EMAs4 | 0.081 | 0.018 | 4.603 | <0.001 | *** |
| EMAs16 - EMAs3 | 0.059 | 0.018 | 3.329 | 0.008 | ** |
| EMAs6 - EMAs5 | 0.062 | 0.018 | 3.422 | 0.006 | ** |
| EMAs6 - EMAs4 | 0.043 | 0.018 | 2.399 | 0.116 |  |
| EMAs6 - EMAs3 | 0.021 | 0.018 | 1.152 | 0.779 |  |
| EMAs5 - EMAs4 | -0.019 | 0.018 | -1.072 | 0.821 |  |
| EMAs5 - EMAs3 | -0.042 | 0.018 | -2.308 | 0.143 |  |
| EMAs4 - EMAs3 | -0.022 | 0.018 | -1.258 | 0.717 |  |
| Note: Estimates are on the response scale (0-1), showing differences in predicted means. | | | | | |
| Positive estimates indicate higher values for the first condition in each contrast; negative estimates indicate higher values for the second condition. | | | | | |
| Significance levels: *** p<0.001, ** p<0.01, * p<0.05; P-values adjusted using Tukey method. | | | | | |

By feature selection

Sensitivity pairwise comparisons by feature selection (Cravings)

| **Contrast** | **Estimate** | **Standard error** | **t-ratio** | **p-value** | **Sig.** |
| --- | --- | --- | --- | --- | --- |
| All features - Selected features | -0.000 | 0.011 | -0.035 | 0.972 |  |
| Note: Estimates are on the response scale (0-1), showing differences in predicted means. | | | | | |
| Positive estimates indicate higher values for the first condition in each contrast; negative estimates indicate higher values for the second condition. | | | | | |
| Significance levels: *** p<0.001, ** p<0.01, * p<0.05; P-values adjusted using Tukey method. | | | | | |

By share of own data in training set

Sensitivity pairwise comparisons by share of own data in training set (Cravings)

| **Contrast** | **Estimate** | **Standard error** | **t-ratio** | **p-value** | **Sig.** |
| --- | --- | --- | --- | --- | --- |
| None - 10% | -0.020 | 0.016 | -1.246 | 0.598 |  |
| None - 20% | 0.034 | 0.016 | 2.103 | 0.153 |  |
| None - 30% | -0.027 | 0.016 | -1.671 | 0.339 |  |
| 10% - 20% | 0.053 | 0.016 | 3.341 | 0.005 | ** |
| 10% - 30% | -0.007 | 0.016 | -0.428 | 0.974 |  |
| 20% - 30% | -0.060 | 0.016 | -3.765 | <0.001 | *** |
| Note: Estimates are on the response scale (0-1), showing differences in predicted means. | | | | | |
| Positive estimates indicate higher values for the first condition in each contrast; negative estimates indicate higher values for the second condition. | | | | | |
| Significance levels: *** p<0.001, ** p<0.01, * p<0.05; P-values adjusted using Tukey method. | | | | | |
