## Supplementary material for "Optimising supervised machine learning algorithms predicting cigarette cravings and lapses for a smoking cessation just-in-time adaptive intervention (JITAI)": S11 Appendix Specificity models and marginal effects

### 1. Comprehensive analysis: Specificity

#### 1.1. Overall models

##### 1.1.1. Main model results

Specificity (Overall) (linear mixed model)

| **Predictor** | **Estimate** | **Standard error** | **95% confidence interval** | **t-statistic** | **p-value** | **Sig.** |
| --- | --- | --- | --- | --- | --- | --- |
| Intercept | 0.802 | 0.049 | [0.706, 0.899] | 16.268 | <0.001 | *** |
| Prompts per day: 6 | -0.070 | 0.018 | [-0.105, -0.035] | -3.943 | <0.001 | *** |
| Prompts per day: 5 | -0.071 | 0.018 | [-0.106, -0.036] | -3.984 | <0.001 | *** |
| Prompts per day: 4 | -0.104 | 0.018 | [-0.139, -0.069] | -5.865 | <0.001 | *** |
| Prompts per day: 3 | -0.132 | 0.018 | [-0.167, -0.098] | -7.456 | <0.001 | *** |
| Feature selection: Selected features | -0.024 | 0.011 | [-0.046, -0.002] | -2.145 | 0.032 | * |
| Share of own data: 10% | -0.077 | 0.016 | [-0.108, -0.047] | -4.930 | <0.001 | *** |
| Share of own data: 20% | -0.075 | 0.016 | [-0.106, -0.044] | -4.698 | <0.001 | *** |
| Share of own data: 30% | -0.133 | 0.016 | [-0.165, -0.102] | -8.353 | <0.001 | *** |
| Outcome: Cravings | -0.005 | 0.013 | [-0.030, 0.021] | -0.350 | 0.726 |  |
| Reference levels: Prompts per day = 16; Feature selection = All features; Share of own data = None; Outcome = Lapses | | | | | | |
| Significance levels: *** p<0.001, ** p<0.01, * p<0.05 | | | | | | |

##### 1.1.2. Pairwise comparisons

By prompts per day

Specificity pairwise comparisons by prompts per day (Overall)

| **Contrast** | **Estimate** | **Standard error** | **t-ratio** | **p-value** | **Sig.** |
| --- | --- | --- | --- | --- | --- |
| EMAs16 - EMAs6 | 0.070 | 0.018 | 3.943 | <0.001 | *** |
| EMAs16 - EMAs5 | 0.071 | 0.018 | 3.984 | <0.001 | *** |
| EMAs16 - EMAs4 | 0.104 | 0.018 | 5.865 | <0.001 | *** |
| EMAs16 - EMAs3 | 0.132 | 0.018 | 7.456 | <0.001 | *** |
| EMAs6 - EMAs5 | 0.001 | 0.018 | 0.056 | 1.000 |  |
| EMAs6 - EMAs4 | 0.034 | 0.018 | 1.875 | 0.331 |  |
| EMAs6 - EMAs3 | 0.062 | 0.018 | 3.470 | 0.005 | ** |
| EMAs5 - EMAs4 | 0.032 | 0.018 | 1.811 | 0.367 |  |
| EMAs5 - EMAs3 | 0.061 | 0.018 | 3.400 | 0.006 | ** |
| EMAs4 - EMAs3 | 0.029 | 0.018 | 1.609 | 0.492 |  |
| Note: Estimates are on the response scale (0-1), showing differences in predicted means. | | | | | |
| Positive estimates indicate higher values for the first condition in each contrast; negative estimates indicate higher values for the second condition. | | | | | |
| Significance levels: *** p<0.001, ** p<0.01, * p<0.05; P-values adjusted using Tukey method. | | | | | |

By feature selection

Specificity pairwise comparisons by feature selection (Overall)

| **Contrast** | **Estimate** | **Standard error** | **t-ratio** | **p-value** | **Sig.** |
| --- | --- | --- | --- | --- | --- |
| All features - Selected features | 0.024 | 0.011 | 2.145 | 0.032 | * |
| Note: Estimates are on the response scale (0-1), showing differences in predicted means. | | | | | |
| Positive estimates indicate higher values for the first condition in each contrast; negative estimates indicate higher values for the second condition. | | | | | |
| Significance levels: *** p<0.001, ** p<0.01, * p<0.05; P-values adjusted using Tukey method. | | | | | |

By share of own data in training set

Specificity pairwise comparisons by share of own data in training set (Overall)

| **Contrast** | **Estimate** | **Standard error** | **t-ratio** | **p-value** | **Sig.** |
| --- | --- | --- | --- | --- | --- |
| None - 10% | 0.077 | 0.016 | 4.930 | <0.001 | *** |
| None - 20% | 0.075 | 0.016 | 4.698 | <0.001 | *** |
| None - 30% | 0.133 | 0.016 | 8.353 | <0.001 | *** |
| 10% - 20% | -0.002 | 0.016 | -0.139 | 0.999 |  |
| 10% - 30% | 0.056 | 0.016 | 3.466 | 0.003 | ** |
| 20% - 30% | 0.058 | 0.016 | 3.566 | 0.002 | ** |
| Note: Estimates are on the response scale (0-1), showing differences in predicted means. | | | | | |
| Positive estimates indicate higher values for the first condition in each contrast; negative estimates indicate higher values for the second condition. | | | | | |
| Significance levels: *** p<0.001, ** p<0.01, * p<0.05; P-values adjusted using Tukey method. | | | | | |

By outcome

Specificity pairwise comparisons by outcome (Overall)

| **Contrast** | **Estimate** | **Standard error** | **t-ratio** | **p-value** | **Sig.** |
| --- | --- | --- | --- | --- | --- |
| Lapses - Cravings | 0.005 | 0.013 | 0.350 | 0.726 |  |
| Note: Estimates are on the response scale (0-1), showing differences in predicted means. | | | | | |
| Positive estimates indicate higher values for the first condition in each contrast; negative estimates indicate higher values for the second condition. | | | | | |
| Significance levels: *** p<0.001, ** p<0.01, * p<0.05; P-values adjusted using Tukey method. | | | | | |

#### 1.2. Lapses models

##### 1.2.1. Main model results

Specificity (Lapses) (linear mixed model)

| **Predictor** | **Estimate** | **Standard error** | **95% confidence interval** | **t-statistic** | **p-value** | **Sig.** |
| --- | --- | --- | --- | --- | --- | --- |
| Intercept | 0.889 | 0.063 | [0.765, 1.013] | 14.047 | <0.001 | *** |
| Prompts per day: 6 | -0.141 | 0.021 | [-0.182, -0.099] | -6.638 | <0.001 | *** |
| Prompts per day: 5 | -0.141 | 0.021 | [-0.183, -0.100] | -6.654 | <0.001 | *** |
| Prompts per day: 4 | -0.185 | 0.021 | [-0.227, -0.144] | -8.741 | <0.001 | *** |
| Prompts per day: 3 | -0.165 | 0.021 | [-0.206, -0.123] | -7.732 | <0.001 | *** |
| Feature selection: Selected features | -0.038 | 0.013 | [-0.064, -0.011] | -2.805 | 0.005 | ** |
| Share of own data: 10% | -0.056 | 0.019 | [-0.093, -0.019] | -2.988 | 0.003 | ** |
| Share of own data: 20% | -0.063 | 0.020 | [-0.102, -0.024] | -3.178 | 0.001 | ** |
| Share of own data: 30% | -0.090 | 0.020 | [-0.129, -0.051] | -4.512 | <0.001 | *** |
| Reference levels: Prompts per day = 16; Feature selection = All features; Share of own data = None | | | | | | |
| Significance levels: *** p<0.001, ** p<0.01, * p<0.05 | | | | | | |

##### 1.2.2. Pairwise comparisons

By prompts per day

Specificity pairwise comparisons by prompts per day (Lapses)

| **Contrast** | **Estimate** | **Standard error** | **t-ratio** | **p-value** | **Sig.** |
| --- | --- | --- | --- | --- | --- |
| EMAs16 - EMAs6 | 0.141 | 0.021 | 6.638 | <0.001 | *** |
| EMAs16 - EMAs5 | 0.141 | 0.021 | 6.653 | <0.001 | *** |
| EMAs16 - EMAs4 | 0.185 | 0.021 | 8.741 | <0.001 | *** |
| EMAs16 - EMAs3 | 0.165 | 0.021 | 7.731 | <0.001 | *** |
| EMAs6 - EMAs5 | 0.000 | 0.021 | 0.015 | 1.000 |  |
| EMAs6 - EMAs4 | 0.045 | 0.021 | 2.102 | 0.220 |  |
| EMAs6 - EMAs3 | 0.024 | 0.021 | 1.117 | 0.798 |  |
| EMAs5 - EMAs4 | 0.044 | 0.021 | 2.086 | 0.227 |  |
| EMAs5 - EMAs3 | 0.023 | 0.021 | 1.101 | 0.806 |  |
| EMAs4 - EMAs3 | -0.021 | 0.021 | -0.978 | 0.865 |  |
| Note: Estimates are on the response scale (0-1), showing differences in predicted means. | | | | | |
| Positive estimates indicate higher values for the first condition in each contrast; negative estimates indicate higher values for the second condition. | | | | | |
| Significance levels: *** p<0.001, ** p<0.01, * p<0.05; P-values adjusted using Tukey method. | | | | | |

By feature selection

Specificity pairwise comparisons by feature selection (Lapses)

| **Contrast** | **Estimate** | **Standard error** | **t-ratio** | **p-value** | **Sig.** |
| --- | --- | --- | --- | --- | --- |
| All features - Selected features | 0.038 | 0.013 | 2.805 | 0.005 | ** |
| Note: Estimates are on the response scale (0-1), showing differences in predicted means. | | | | | |
| Positive estimates indicate higher values for the first condition in each contrast; negative estimates indicate higher values for the second condition. | | | | | |
| Significance levels: *** p<0.001, ** p<0.01, * p<0.05; P-values adjusted using Tukey method. | | | | | |

By share of own data in training set

Specificity pairwise comparisons by share of own data in training set (Lapses)

| **Contrast** | **Estimate** | **Standard error** | **t-ratio** | **p-value** | **Sig.** |
| --- | --- | --- | --- | --- | --- |
| None - 10% | 0.056 | 0.019 | 2.987 | 0.015 | * |
| None - 20% | 0.063 | 0.020 | 3.177 | 0.008 | ** |
| None - 30% | 0.090 | 0.020 | 4.511 | <0.001 | *** |
| 10% - 20% | 0.007 | 0.020 | 0.363 | 0.984 |  |
| 10% - 30% | 0.034 | 0.020 | 1.695 | 0.327 |  |
| 20% - 30% | 0.027 | 0.020 | 1.307 | 0.559 |  |
| Note: Estimates are on the response scale (0-1), showing differences in predicted means. | | | | | |
| Positive estimates indicate higher values for the first condition in each contrast; negative estimates indicate higher values for the second condition. | | | | | |
| Significance levels: *** p<0.001, ** p<0.01, * p<0.05; P-values adjusted using Tukey method. | | | | | |

#### 1.3. Cravings models

##### 1.3.1. Main model results

Specificity (Cravings) (linear mixed model)

| **Predictor** | **Estimate** | **Standard error** | **95% confidence interval** | **t-statistic** | **p-value** | **Sig.** |
| --- | --- | --- | --- | --- | --- | --- |
| Intercept | 0.747 | 0.053 | [0.643, 0.851] | 14.096 | <0.001 | *** |
| Prompts per day: 6 | -0.026 | 0.018 | [-0.062, 0.009] | -1.469 | 0.142 |  |
| Prompts per day: 5 | -0.026 | 0.018 | [-0.062, 0.010] | -1.432 | 0.152 |  |
| Prompts per day: 4 | -0.055 | 0.018 | [-0.090, -0.020] | -3.102 | 0.002 | ** |
| Prompts per day: 3 | -0.107 | 0.018 | [-0.142, -0.072] | -5.987 | <0.001 | *** |
| Feature selection: Selected features | -0.017 | 0.011 | [-0.039, 0.005] | -1.481 | 0.139 |  |
| Share of own data: 10% | -0.069 | 0.016 | [-0.100, -0.037] | -4.268 | <0.001 | *** |
| Share of own data: 20% | -0.039 | 0.016 | [-0.070, -0.007] | -2.419 | 0.016 | * |
| Share of own data: 30% | -0.113 | 0.016 | [-0.144, -0.081] | -6.995 | <0.001 | *** |
| Reference levels: Prompts per day = 16; Feature selection = All features; Share of own data = None | | | | | | |
| Significance levels: *** p<0.001, ** p<0.01, * p<0.05 | | | | | | |

##### 1.3.2. Pairwise comparisons

By prompts per day

Specificity pairwise comparisons by prompts per day (Cravings)

| **Contrast** | **Estimate** | **Standard error** | **t-ratio** | **p-value** | **Sig.** |
| --- | --- | --- | --- | --- | --- |
| EMAs16 - EMAs6 | 0.026 | 0.018 | 1.468 | 0.583 |  |
| EMAs16 - EMAs5 | 0.026 | 0.018 | 1.432 | 0.607 |  |
| EMAs16 - EMAs4 | 0.055 | 0.018 | 3.102 | 0.017 | * |
| EMAs16 - EMAs3 | 0.107 | 0.018 | 5.987 | <0.001 | *** |
| EMAs6 - EMAs5 | -0.000 | 0.018 | -0.027 | 1.000 |  |
| EMAs6 - EMAs4 | 0.029 | 0.018 | 1.595 | 0.501 |  |
| EMAs6 - EMAs3 | 0.080 | 0.018 | 4.460 | <0.001 | *** |
| EMAs5 - EMAs4 | 0.029 | 0.018 | 1.612 | 0.490 |  |
| EMAs5 - EMAs3 | 0.081 | 0.018 | 4.458 | <0.001 | *** |
| EMAs4 - EMAs3 | 0.052 | 0.018 | 2.895 | 0.031 | * |
| Note: Estimates are on the response scale (0-1), showing differences in predicted means. | | | | | |
| Positive estimates indicate higher values for the first condition in each contrast; negative estimates indicate higher values for the second condition. | | | | | |
| Significance levels: *** p<0.001, ** p<0.01, * p<0.05; P-values adjusted using Tukey method. | | | | | |

By feature selection

Specificity pairwise comparisons by feature selection (Cravings)

| **Contrast** | **Estimate** | **Standard error** | **t-ratio** | **p-value** | **Sig.** |
| --- | --- | --- | --- | --- | --- |
| All features - Selected features | 0.017 | 0.011 | 1.481 | 0.139 |  |
| Note: Estimates are on the response scale (0-1), showing differences in predicted means. | | | | | |
| Positive estimates indicate higher values for the first condition in each contrast; negative estimates indicate higher values for the second condition. | | | | | |
| Significance levels: *** p<0.001, ** p<0.01, * p<0.05; P-values adjusted using Tukey method. | | | | | |

By share of own data in training set

Specificity pairwise comparisons by share of own data in training set (Cravings)

| **Contrast** | **Estimate** | **Standard error** | **t-ratio** | **p-value** | **Sig.** |
| --- | --- | --- | --- | --- | --- |
| None - 10% | 0.069 | 0.016 | 4.268 | <0.001 | *** |
| None - 20% | 0.039 | 0.016 | 2.419 | 0.074 |  |
| None - 30% | 0.113 | 0.016 | 6.995 | <0.001 | *** |
| 10% - 20% | -0.030 | 0.016 | -1.835 | 0.257 |  |
| 10% - 30% | 0.044 | 0.016 | 2.734 | 0.032 | * |
| 20% - 30% | 0.074 | 0.016 | 4.565 | <0.001 | *** |
| Note: Estimates are on the response scale (0-1), showing differences in predicted means. | | | | | |
| Positive estimates indicate higher values for the first condition in each contrast; negative estimates indicate higher values for the second condition. | | | | | |
| Significance levels: *** p<0.001, ** p<0.01, * p<0.05; P-values adjusted using Tukey method. | | | | | |
