## Supplementary material for "Optimising supervised machine learning algorithms predicting cigarette cravings and lapses for a smoking cessation just-in-time adaptive intervention (JITAI)": S12 Appendix Time-varying variables distributions

Time-varying variables distributions in full datasets (16 EMA prompts/day)

### Categorical predictors

#### Alcohol use

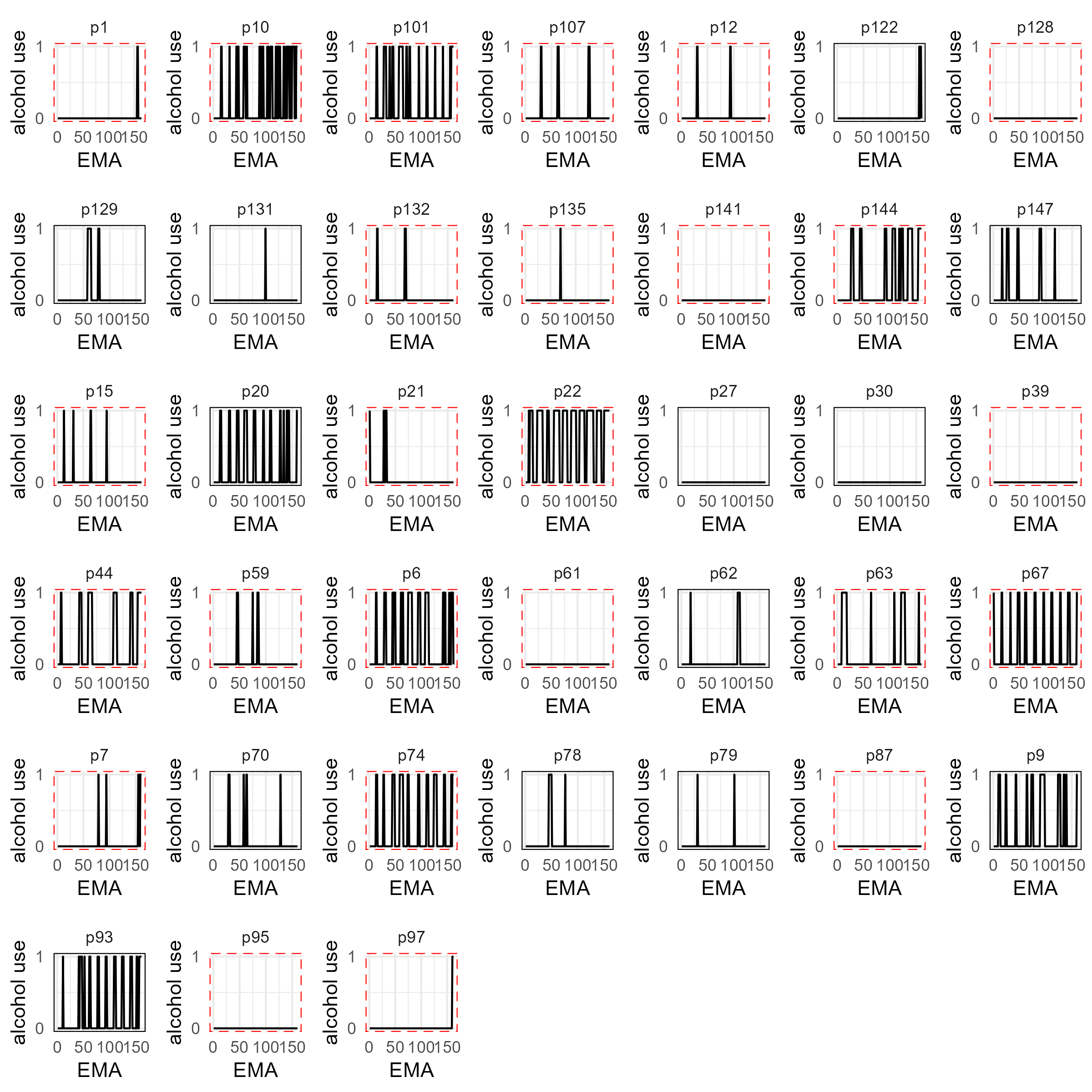

#### Caffeine use

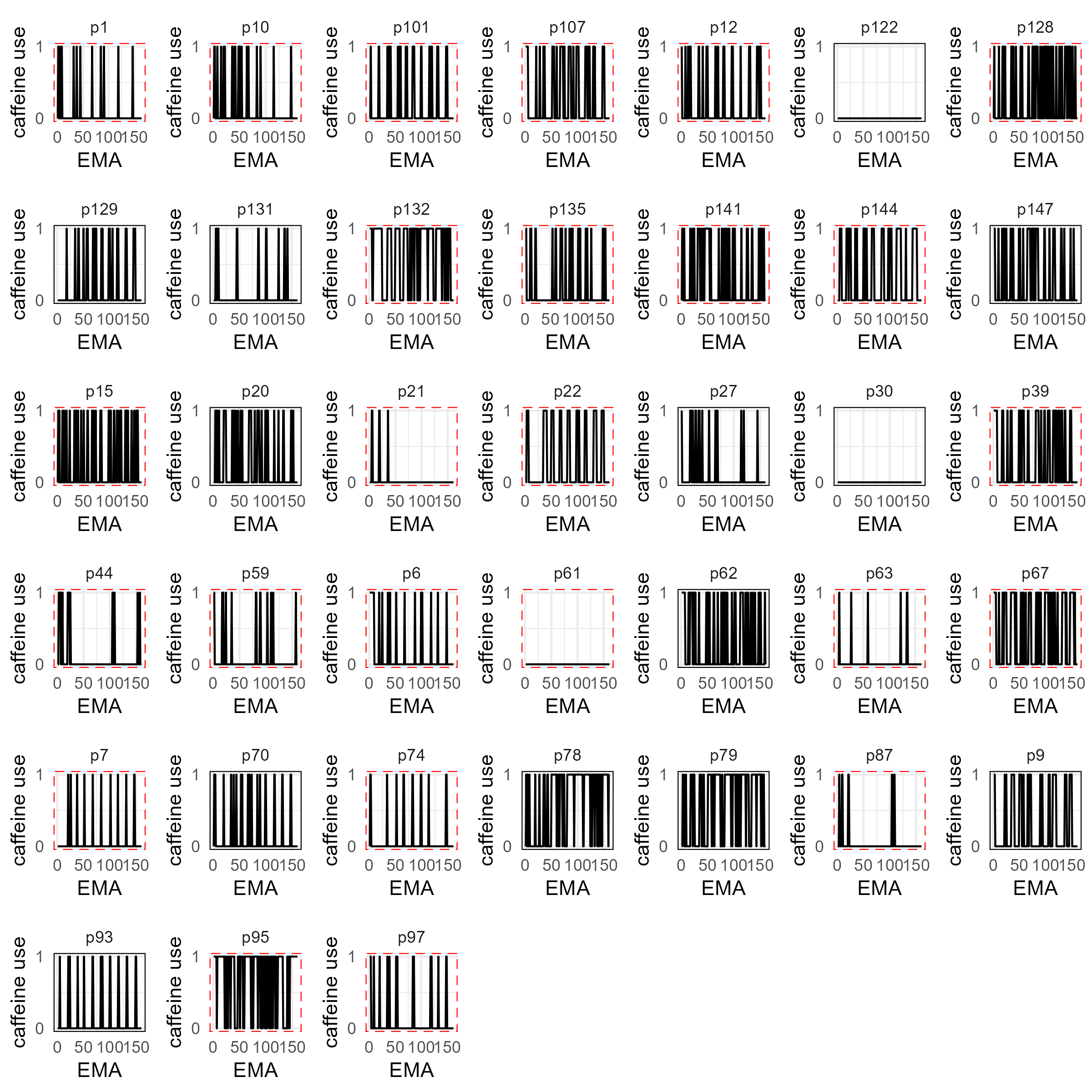

#### Cigarette availability

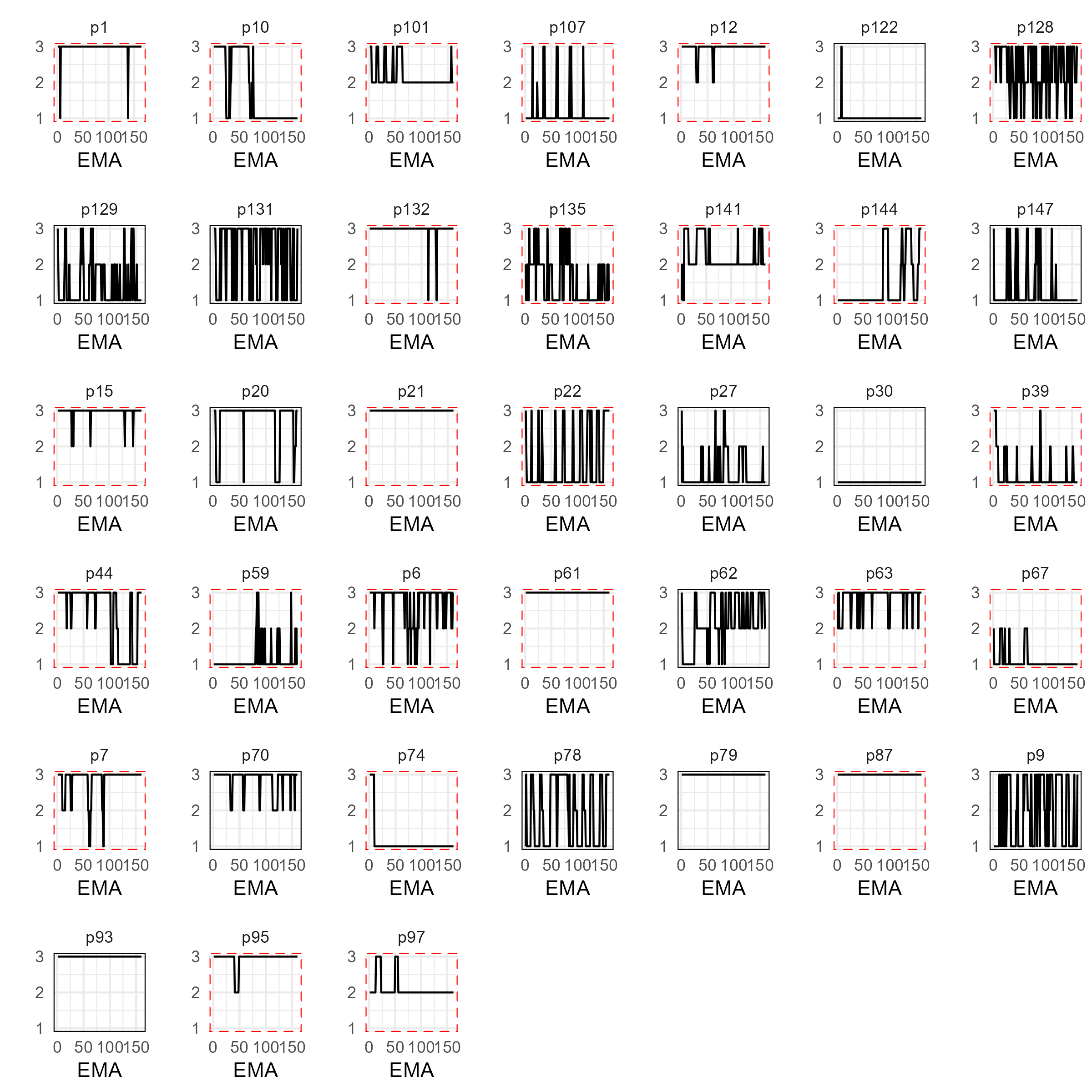

#### Nicotine use

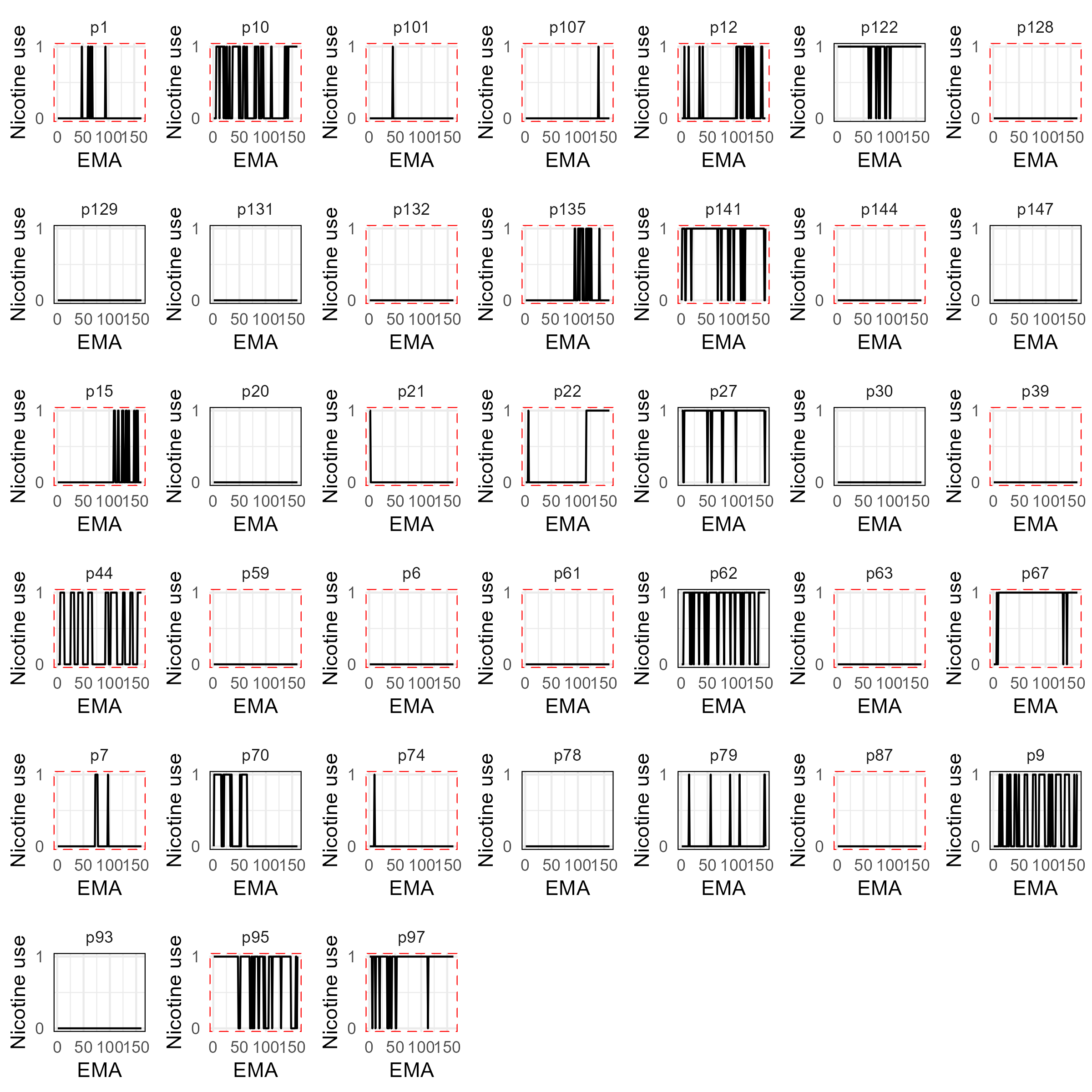

#### High cravings

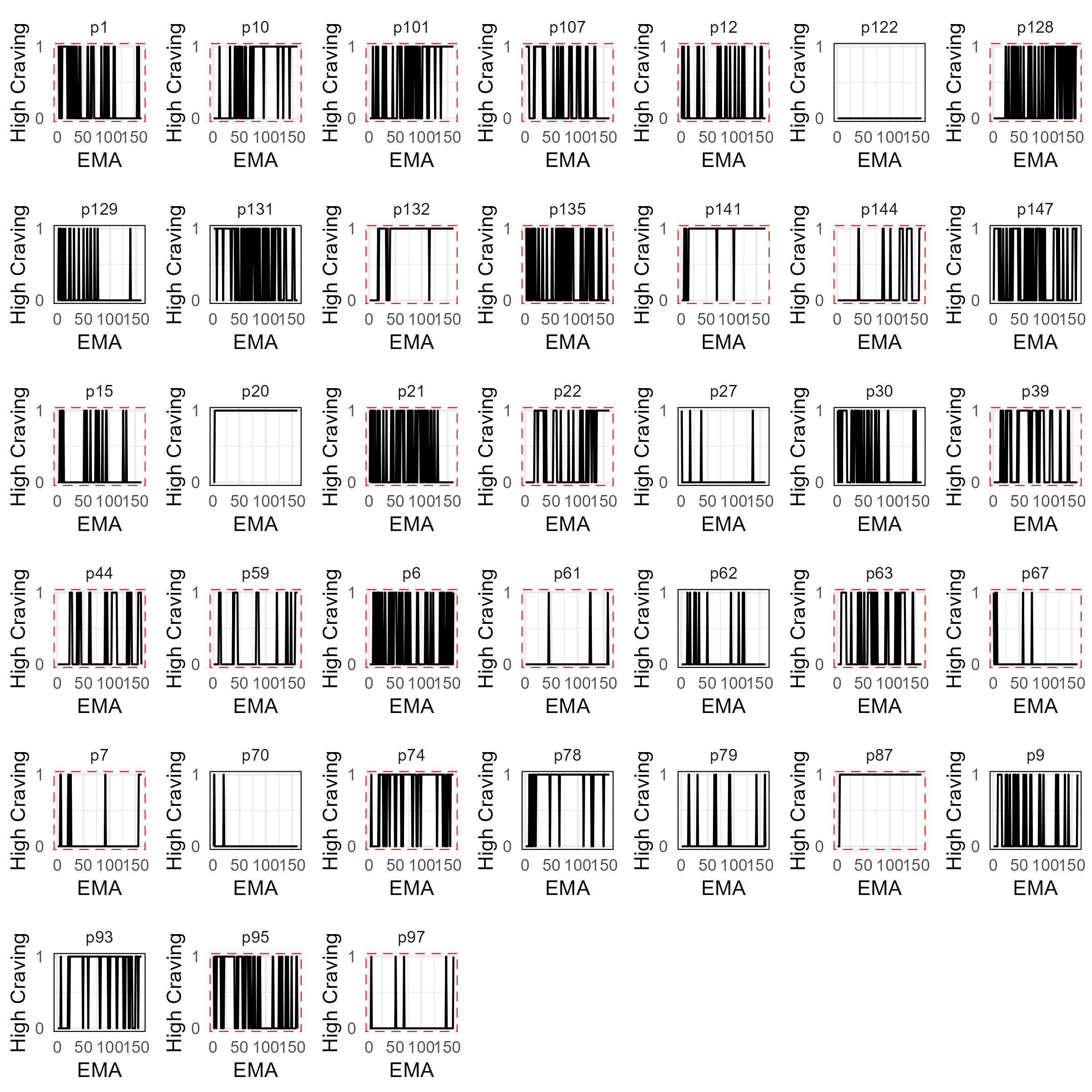

#### Lapses

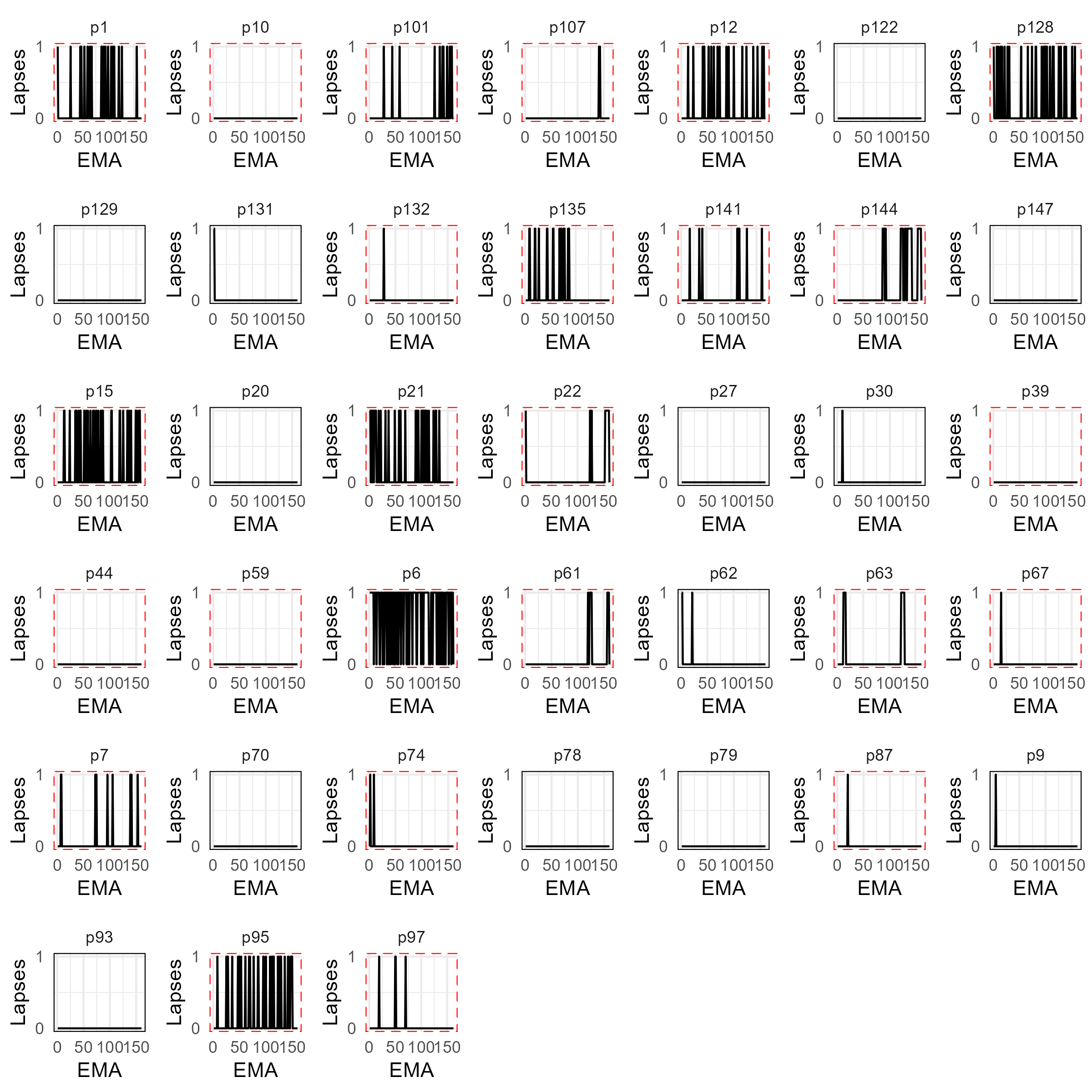

### Continuous predictors

#### Anxiety

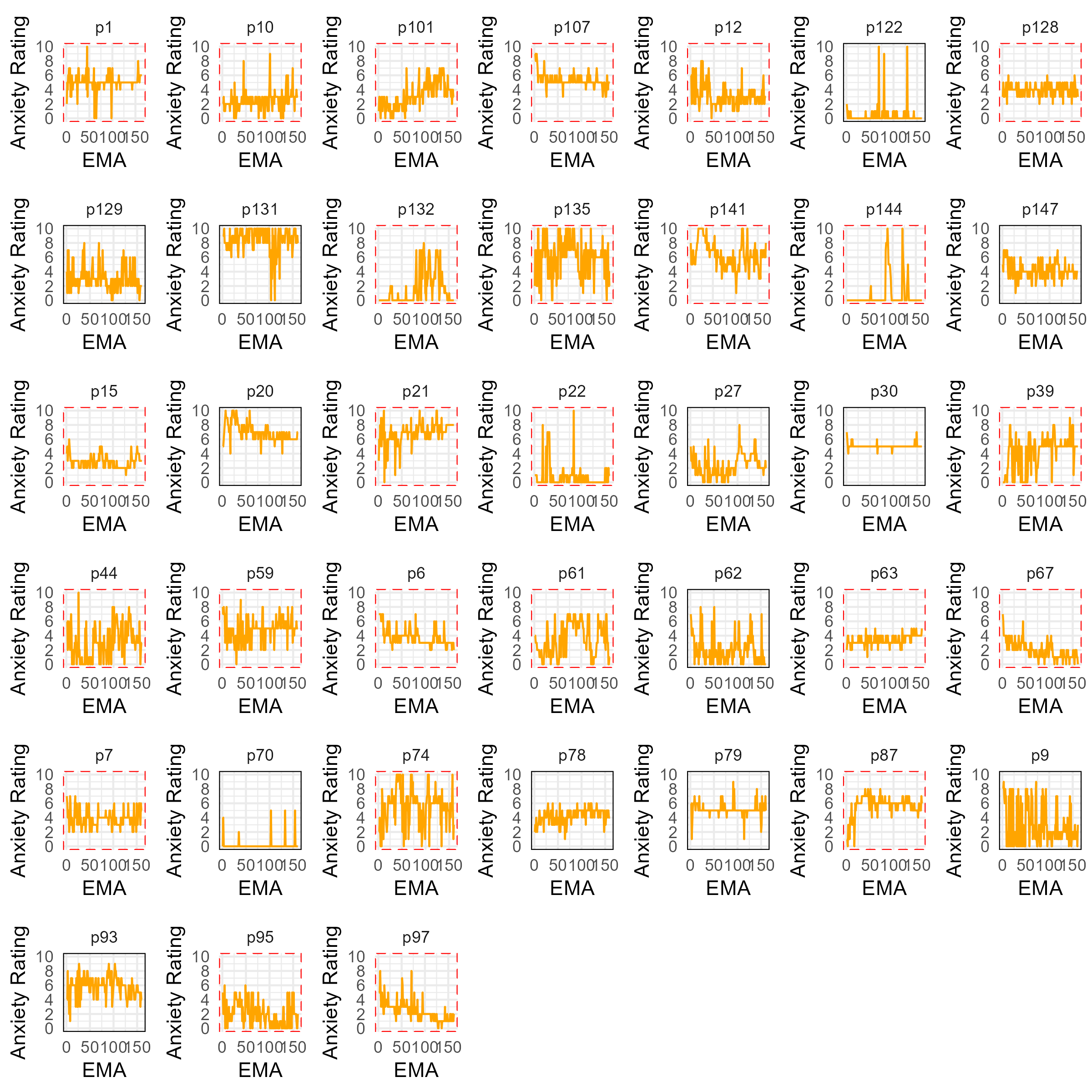

#### Boredom

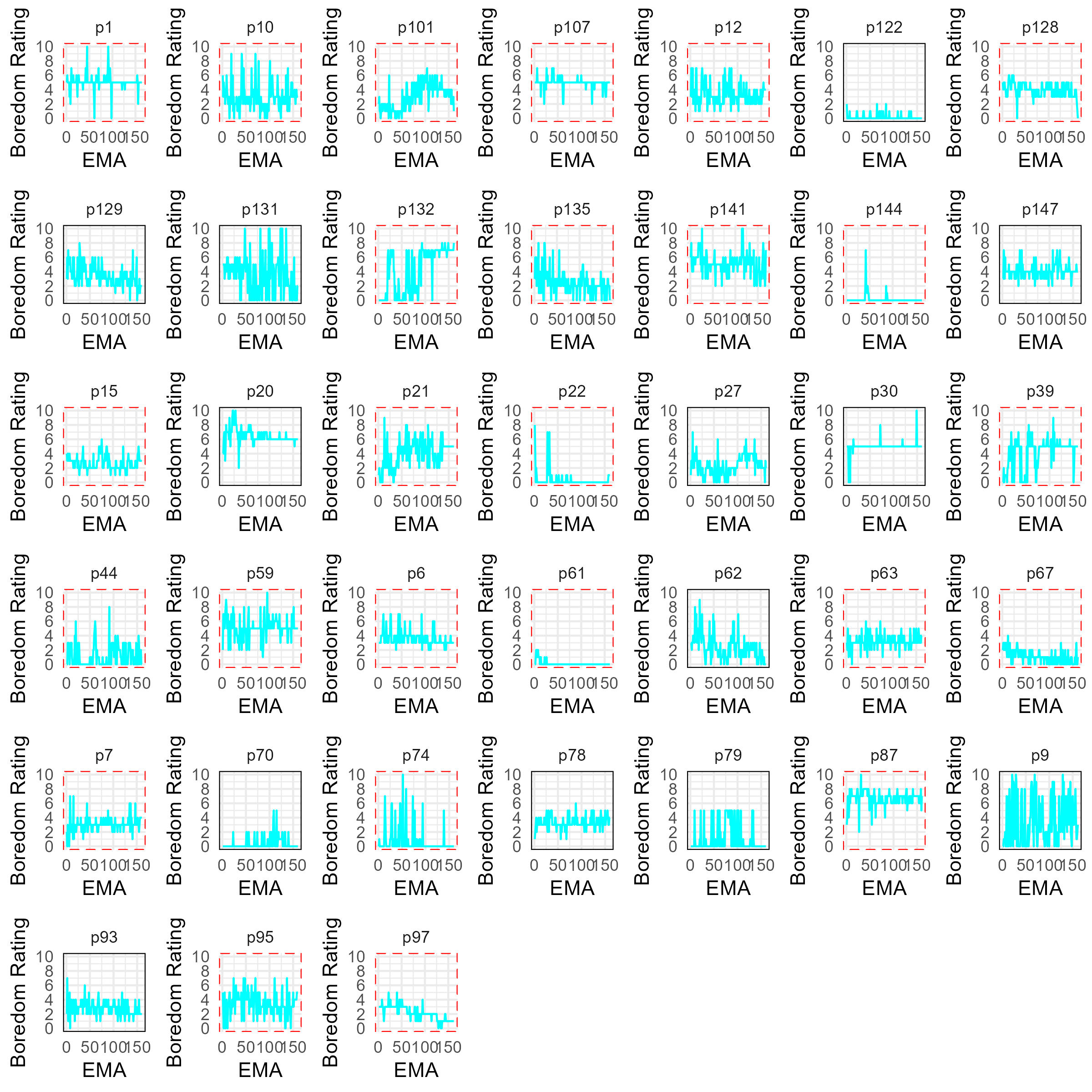

#### Calmness

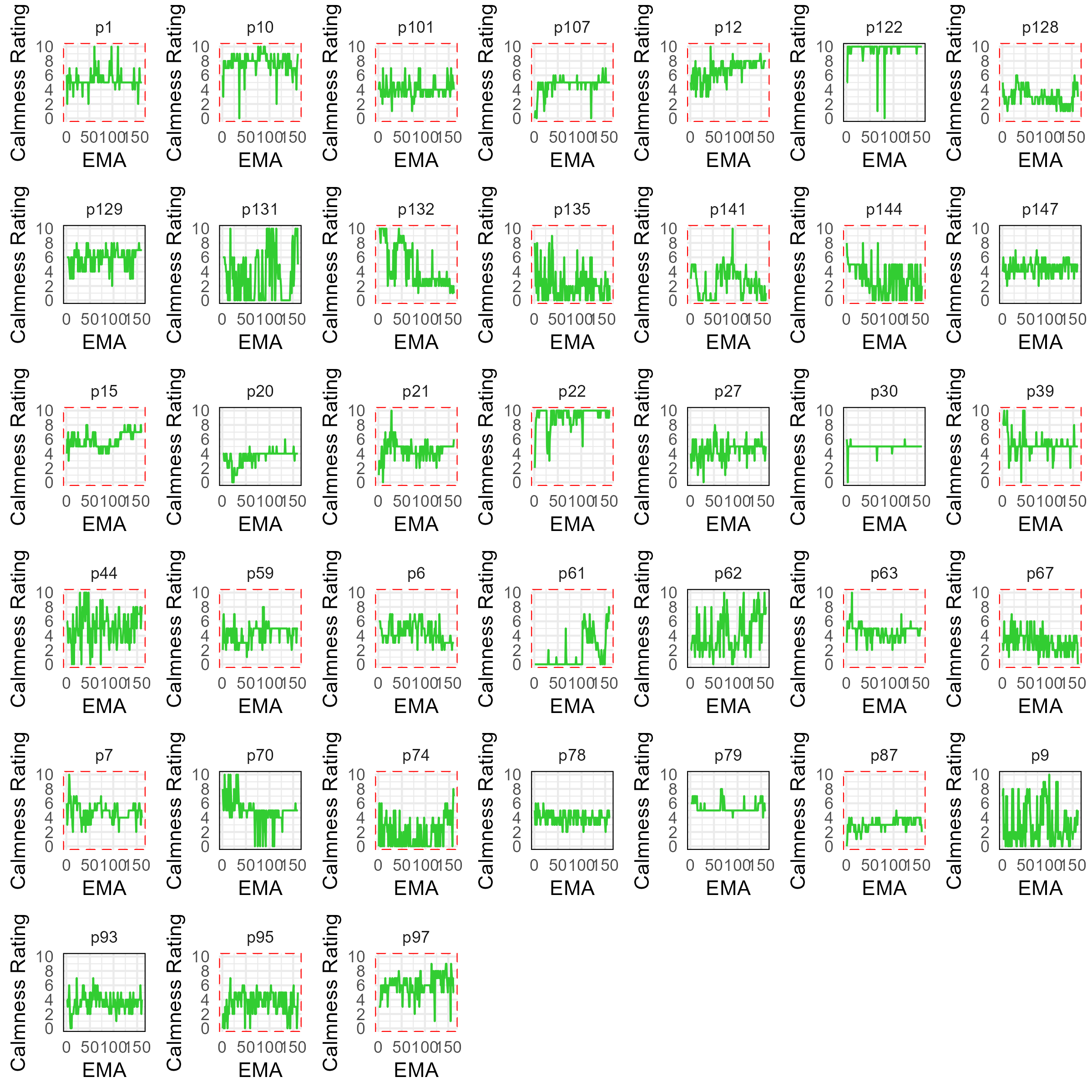

#### Self-efficacy

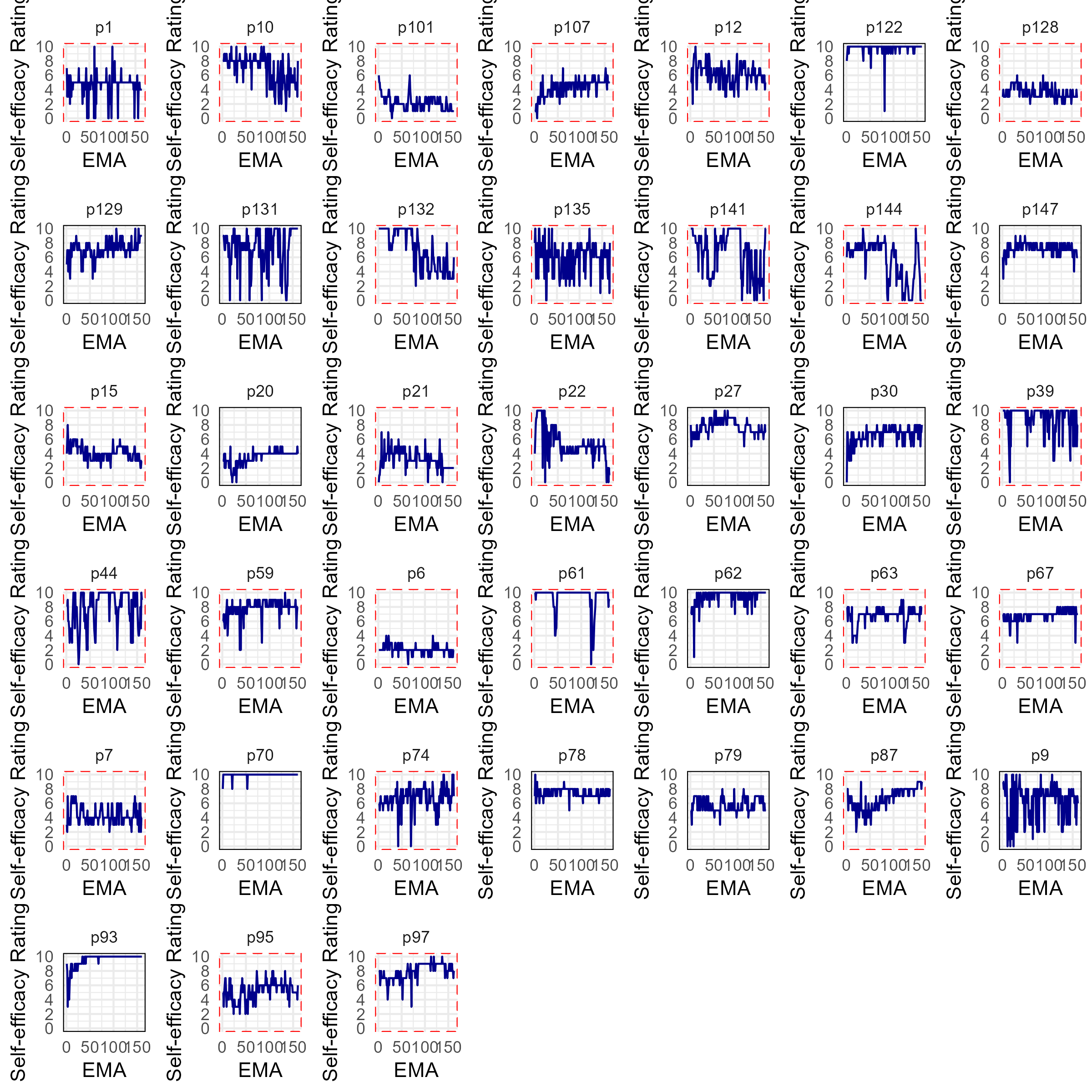

#### Contended

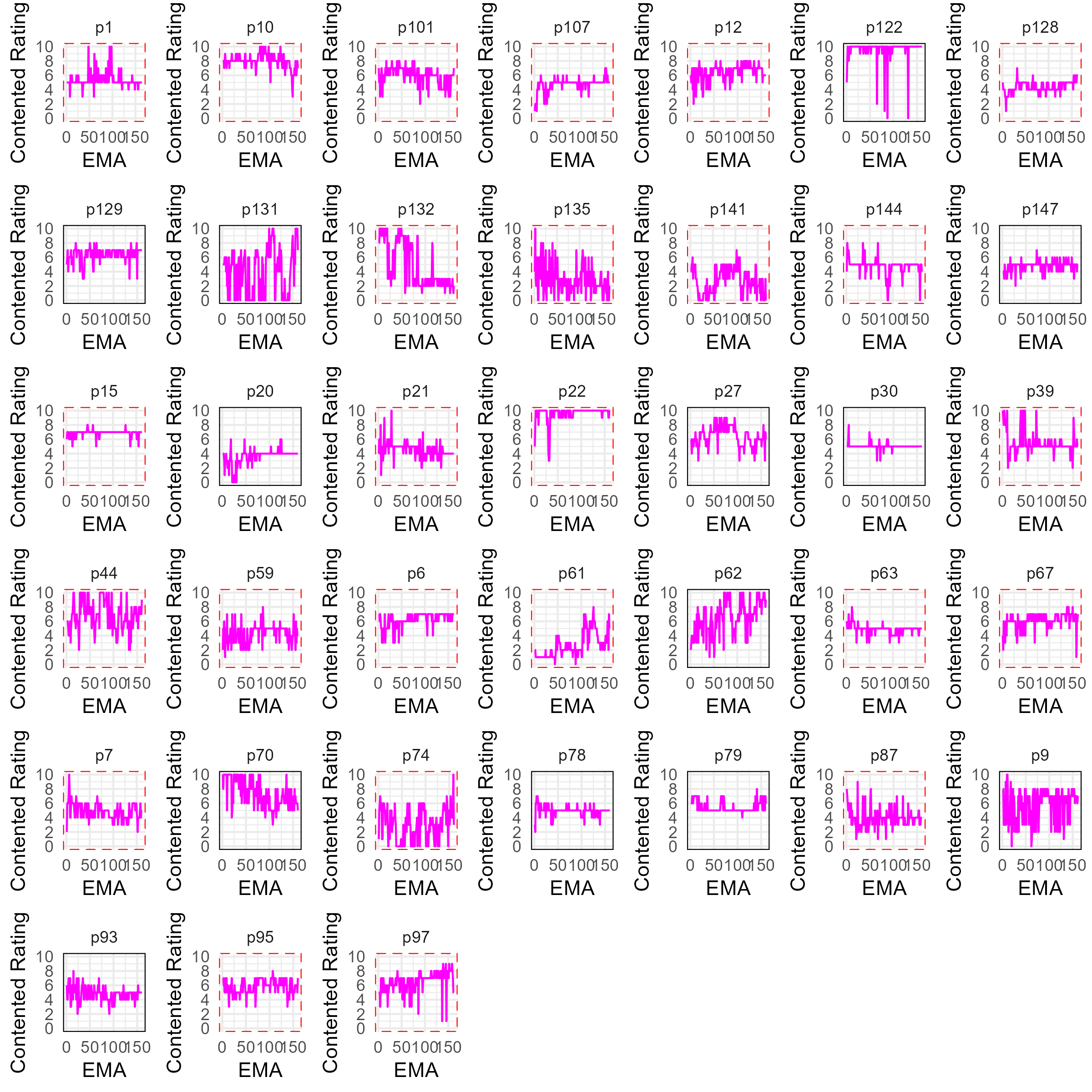

#### Cravings

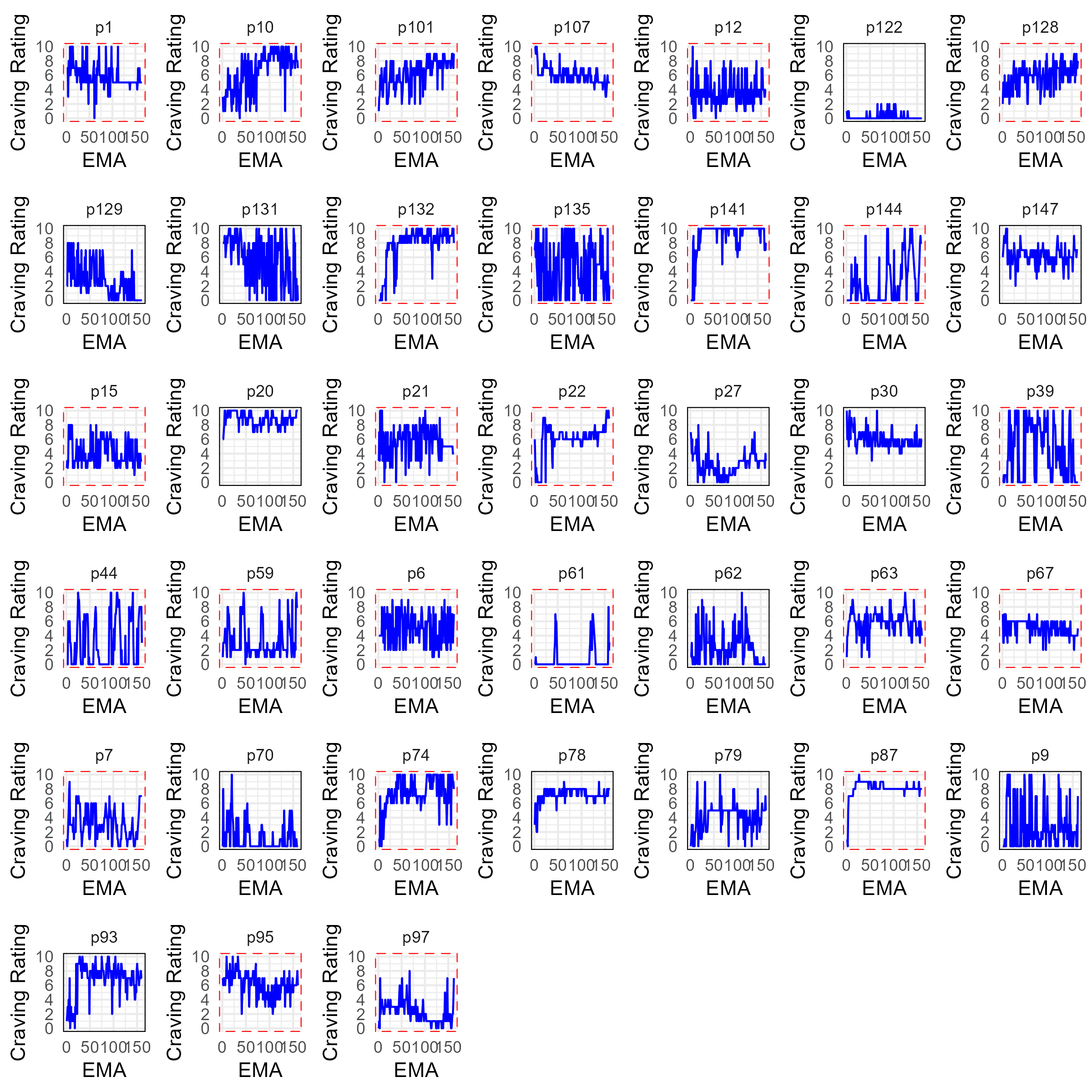

#### Enthusiasm

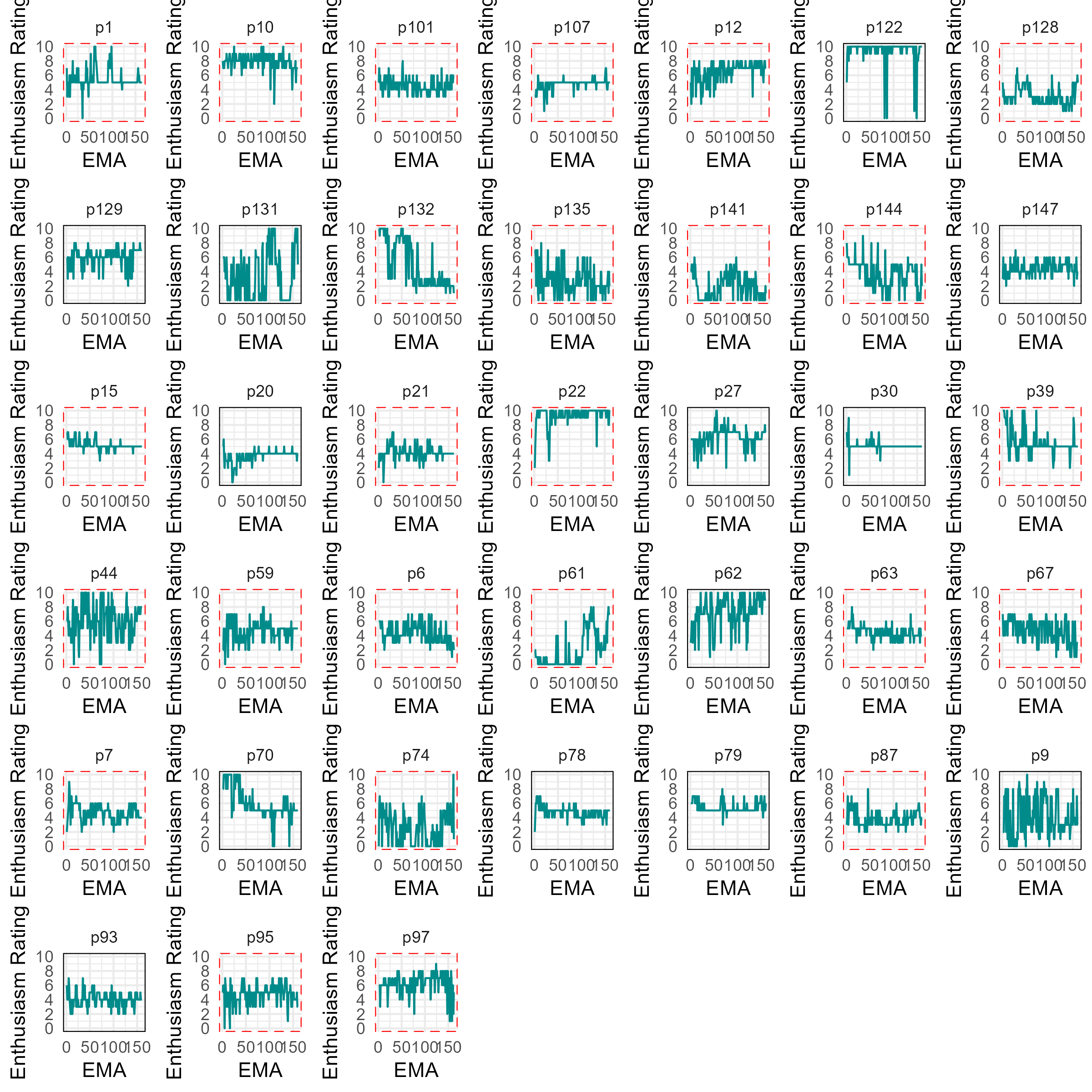

#### Excitement

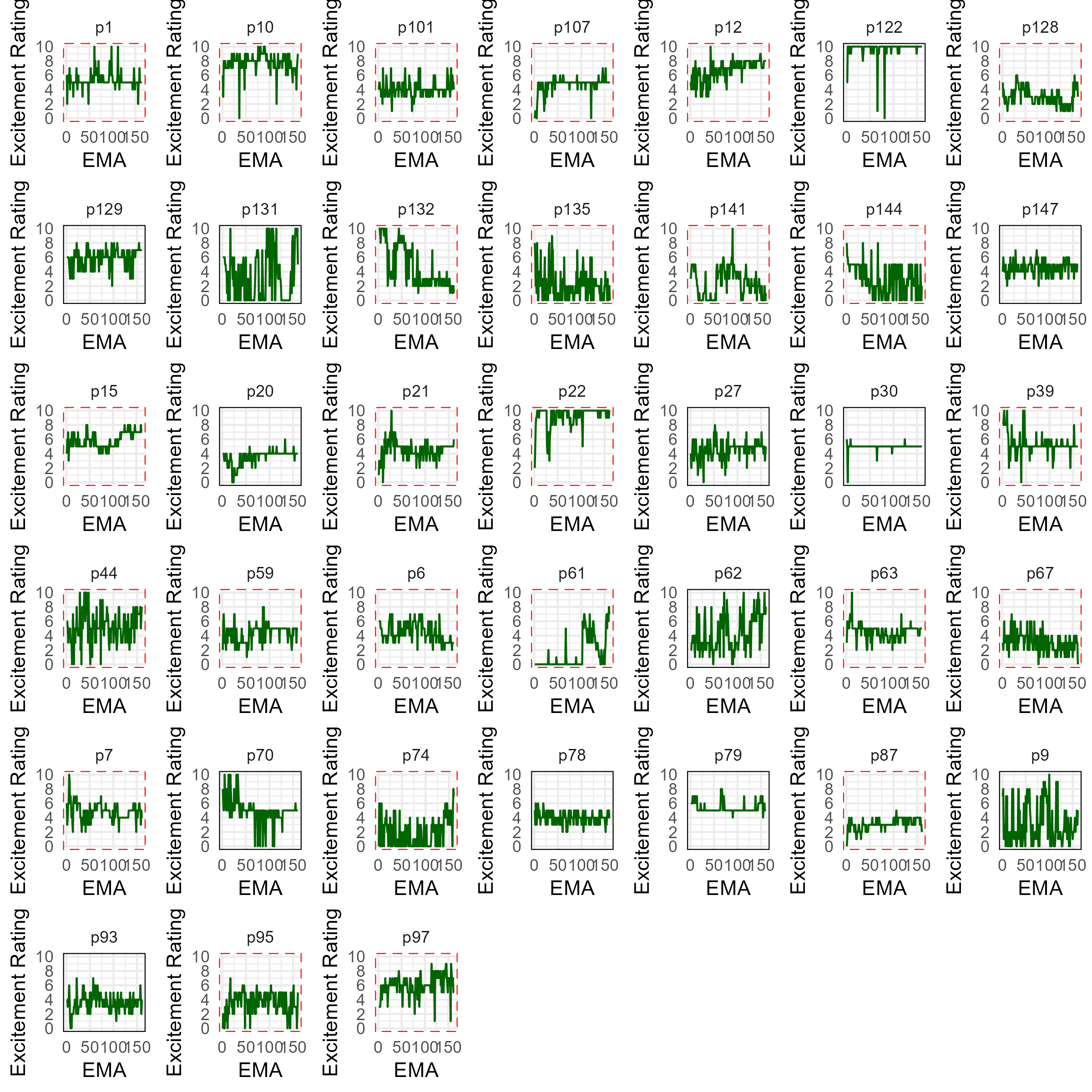

#### Happiness

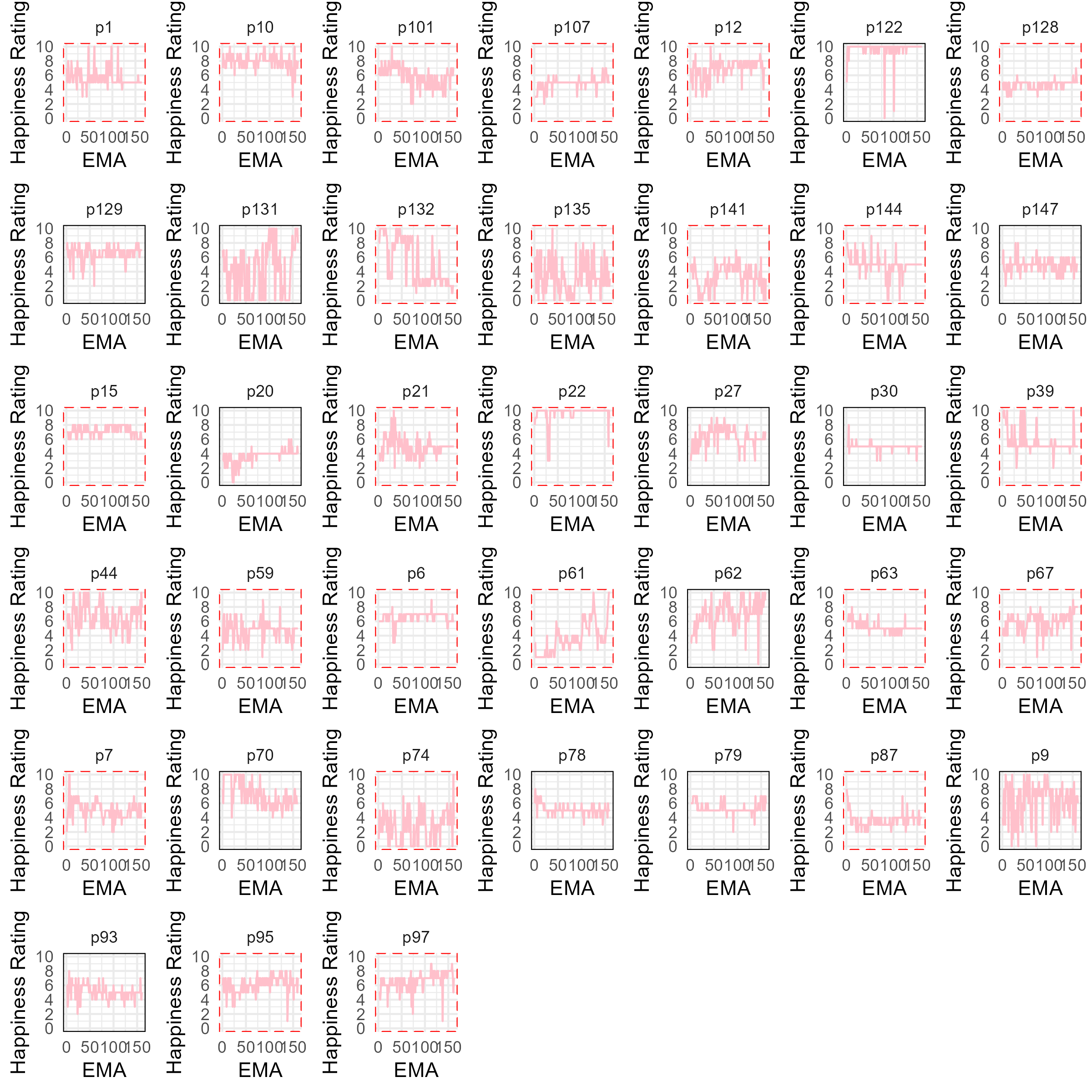

#### Irritability

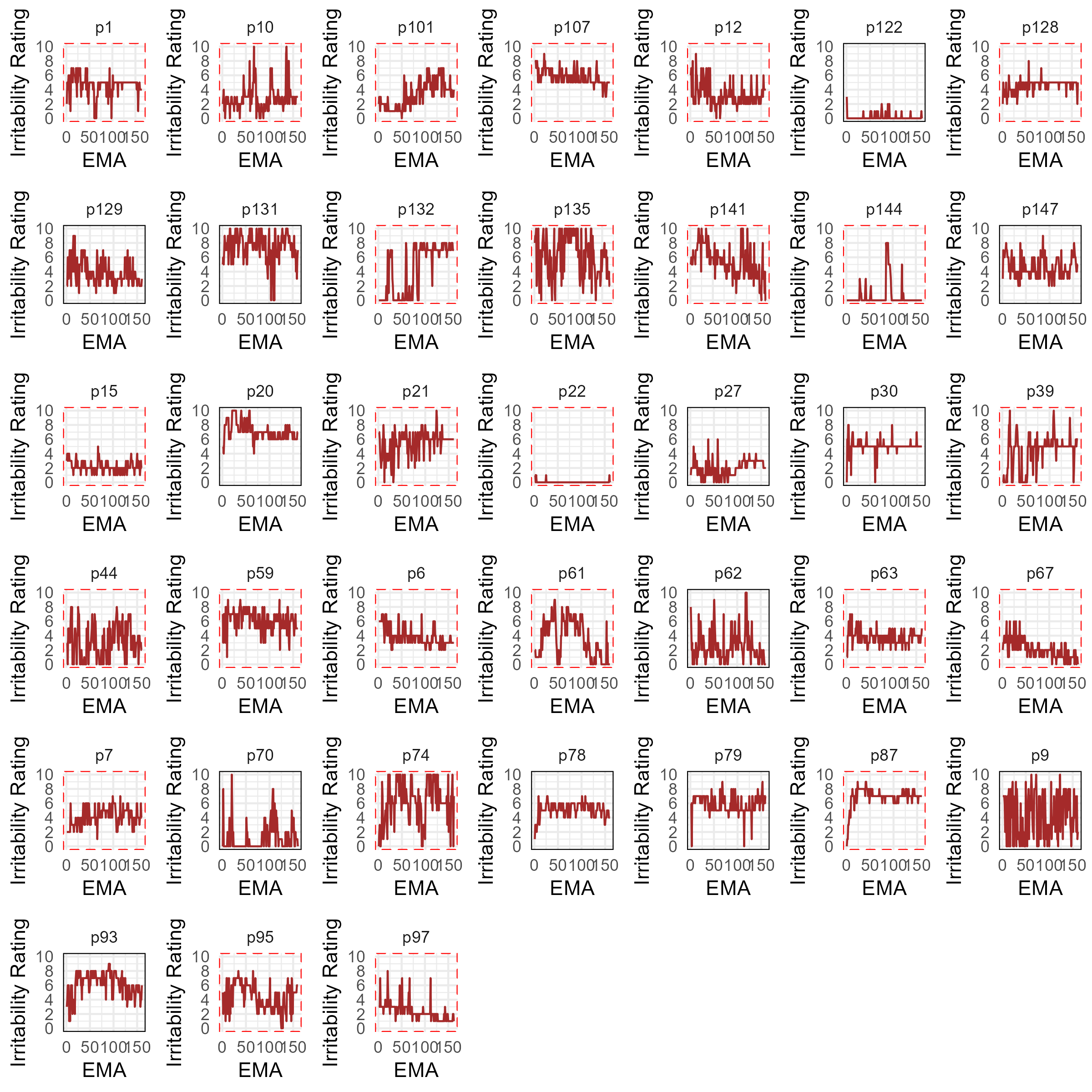

#### Momentary motivation

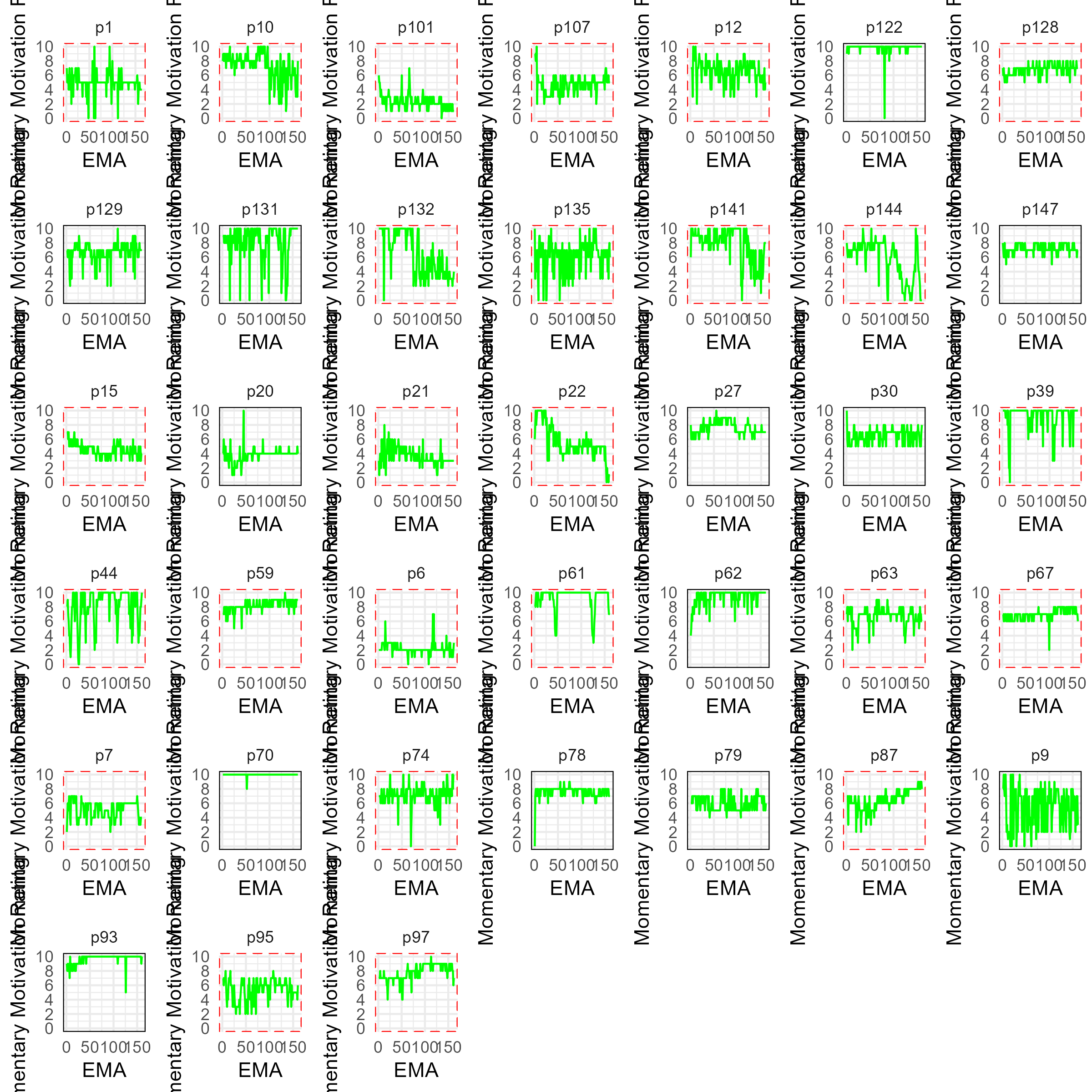

#### Pain

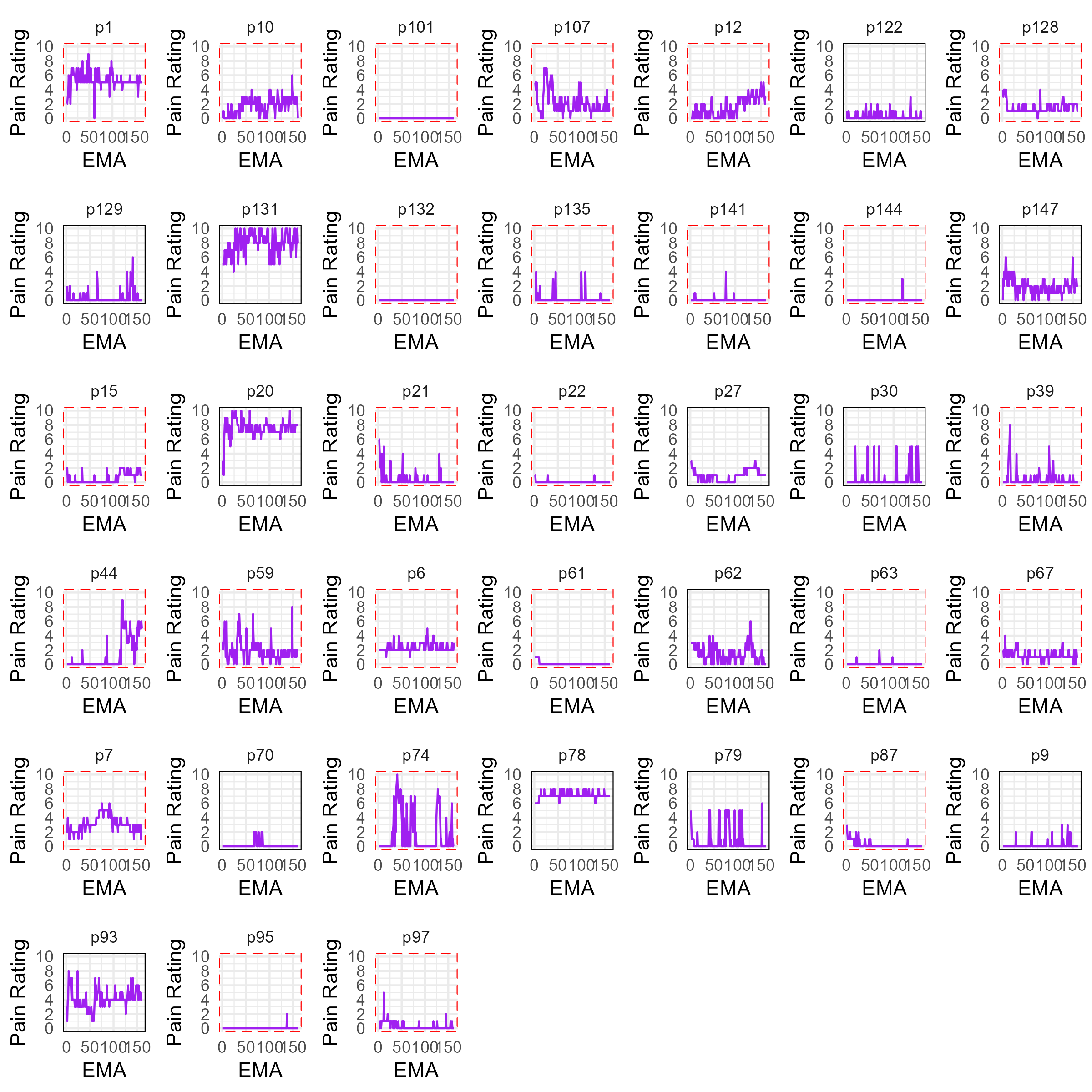

#### Sadness

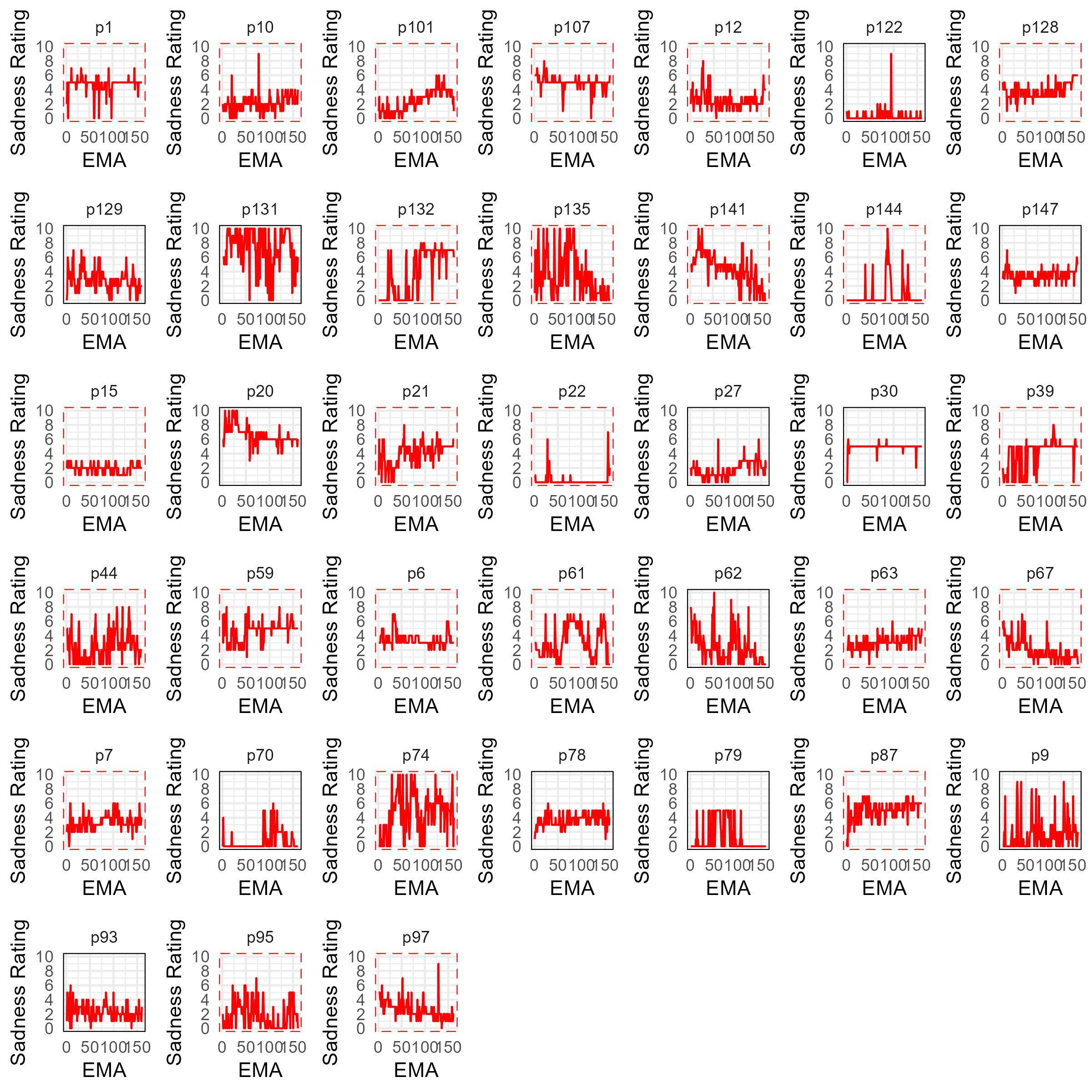

#### Stress

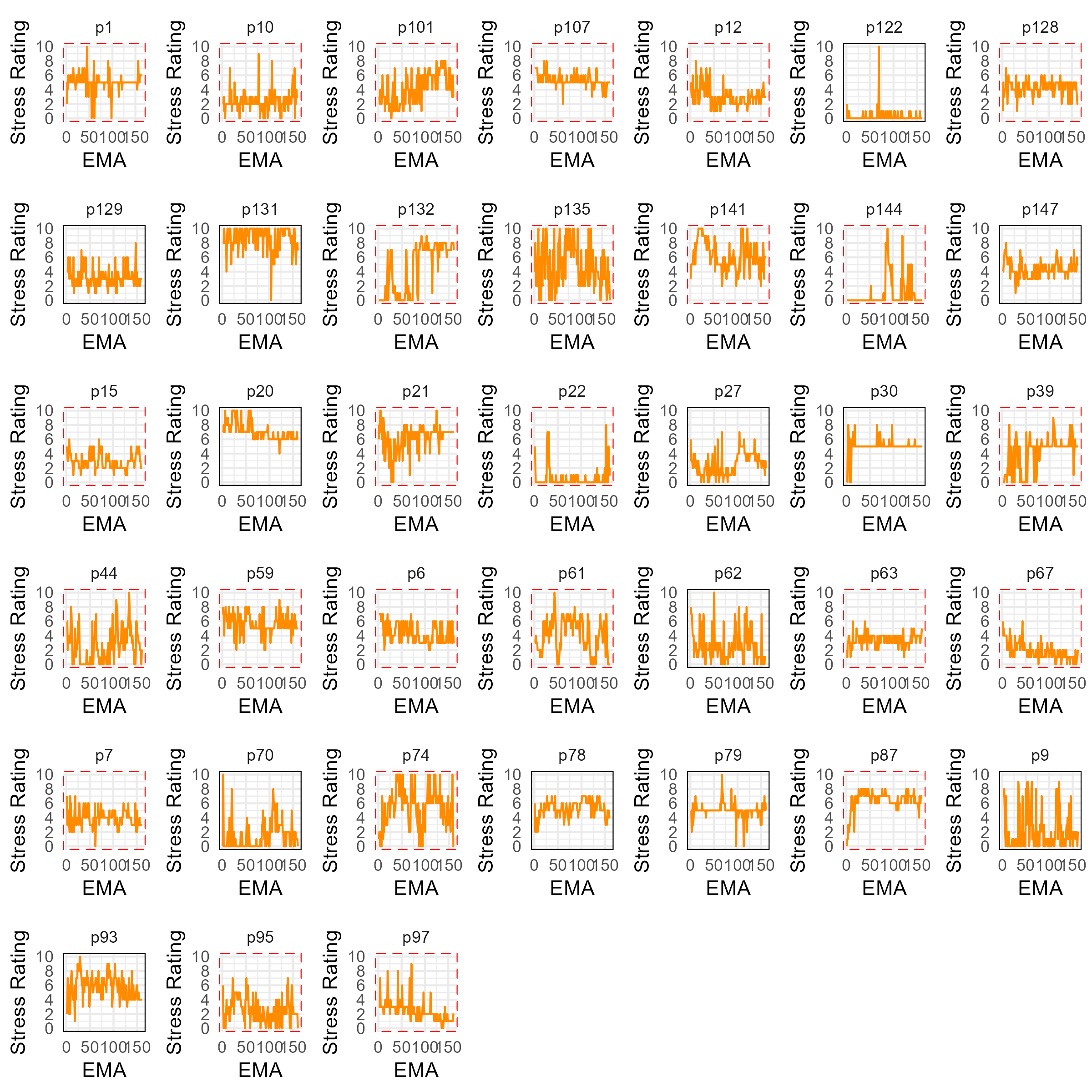

### Context variables

#### Activities

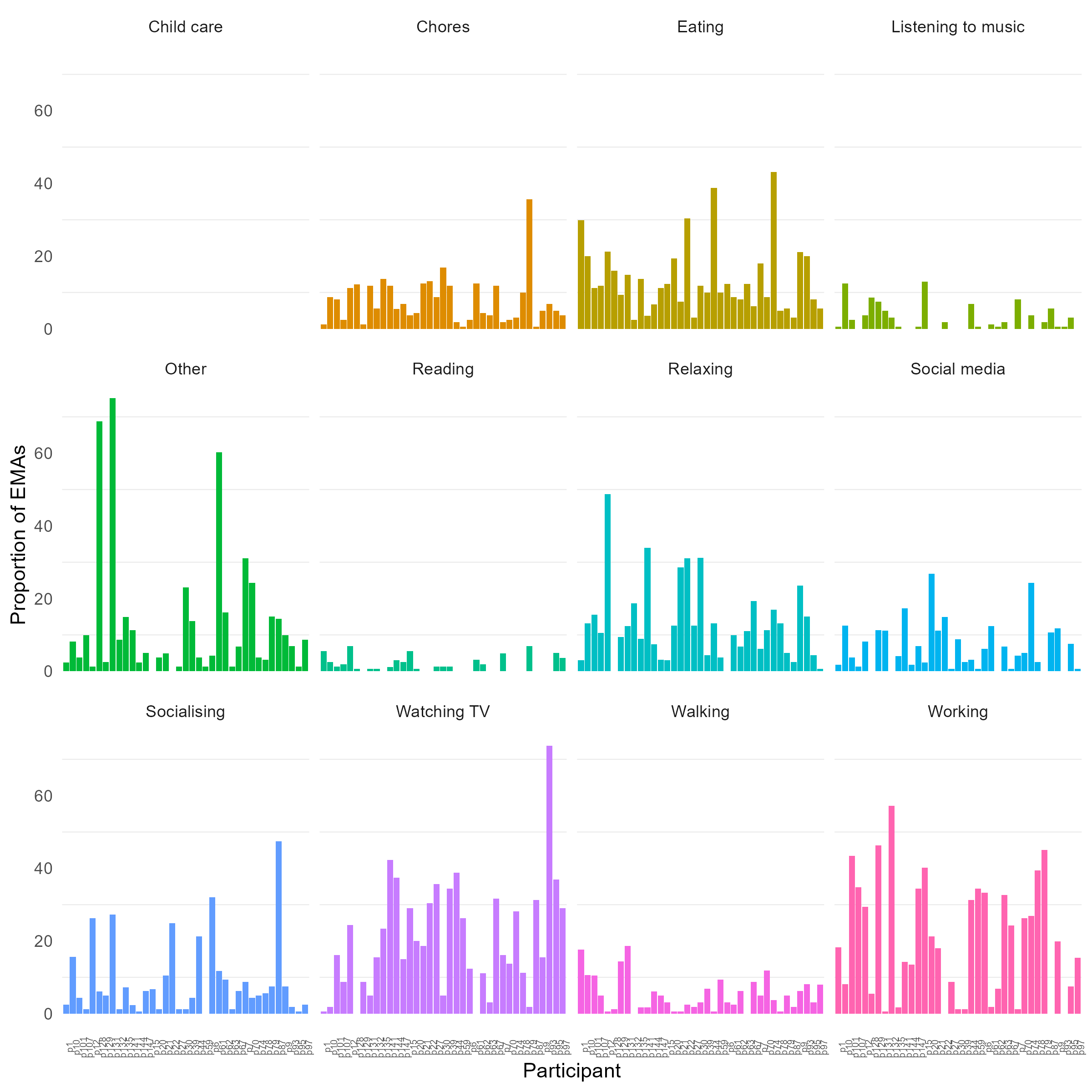

#### Physical context

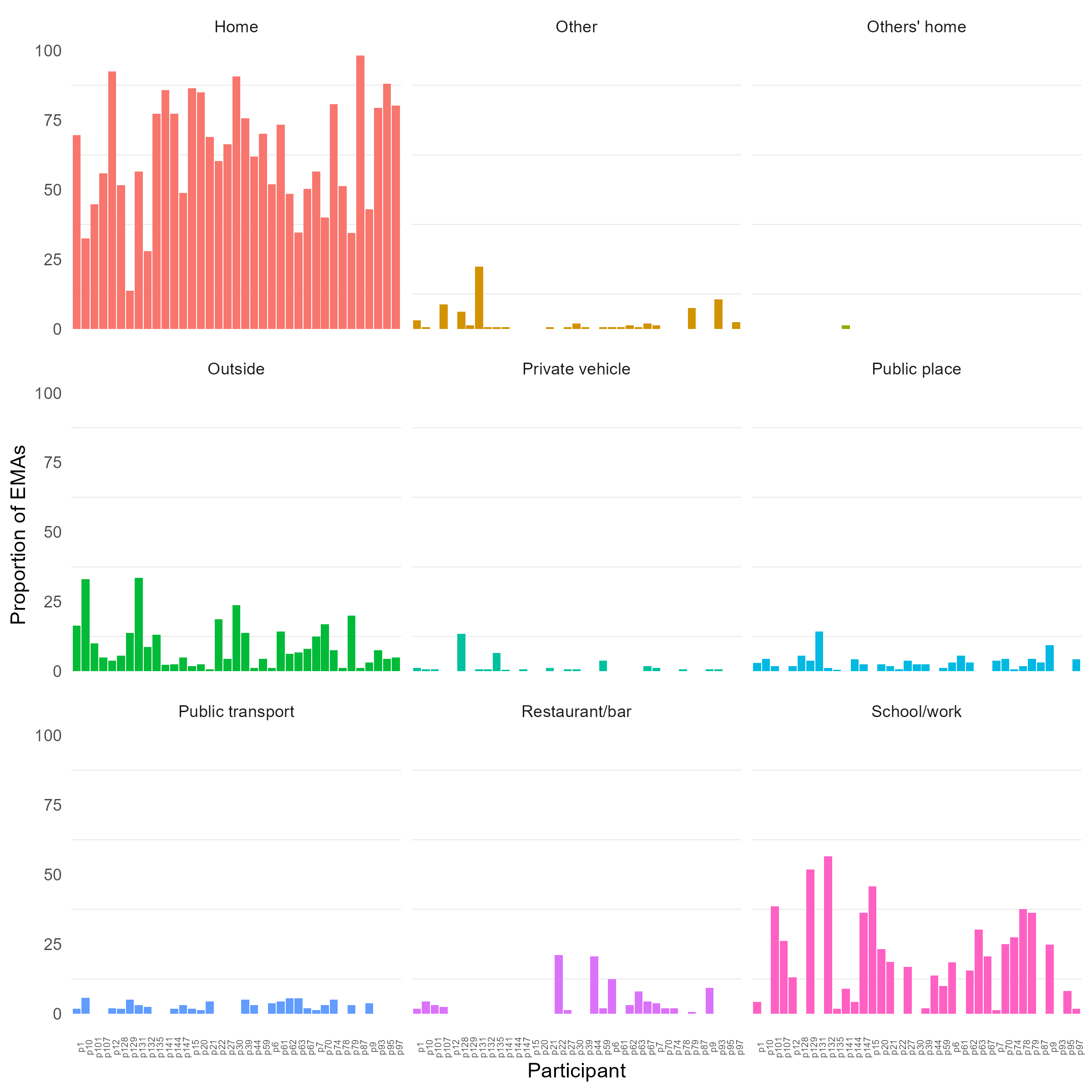

#### Social context

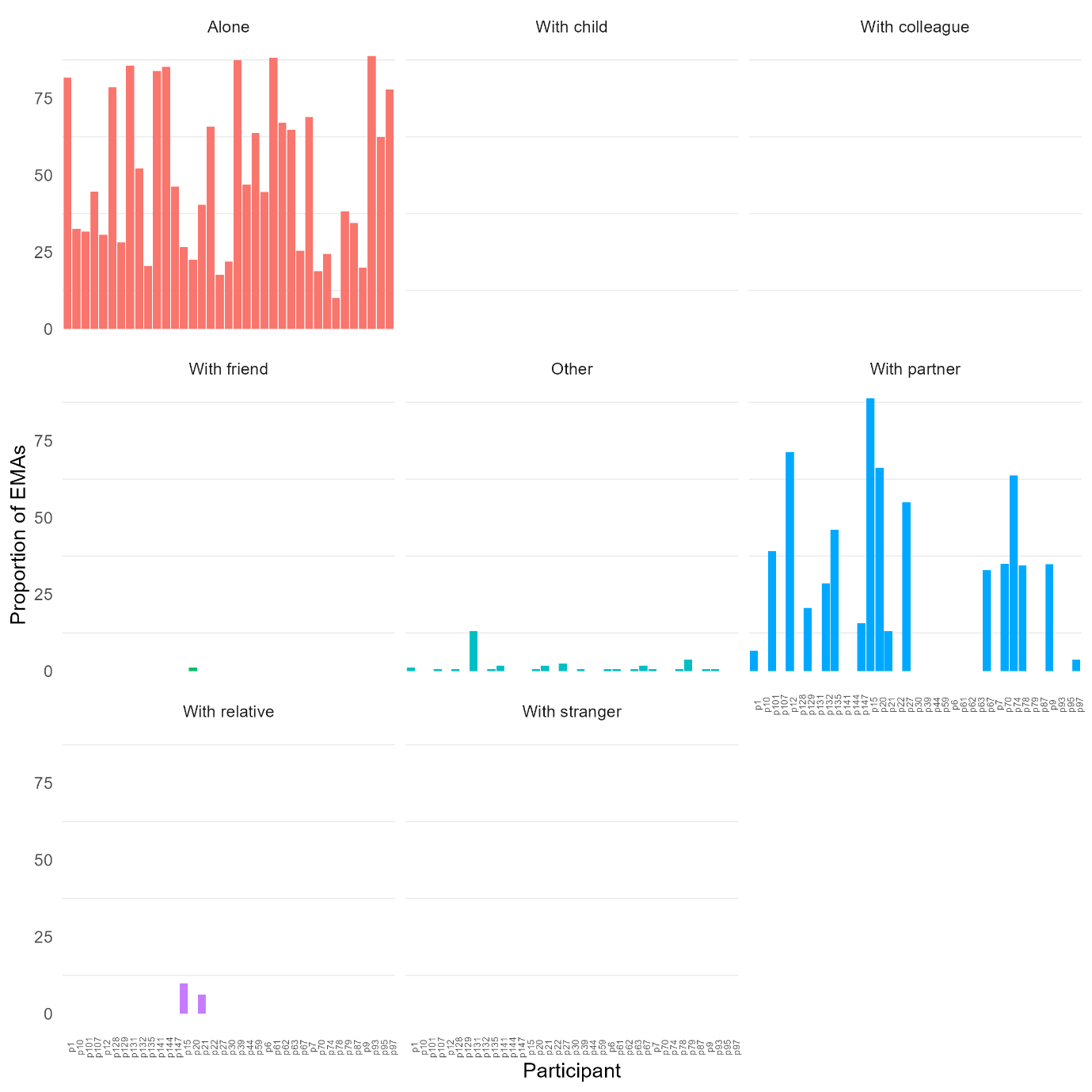
